# Comparison of Population Characteristics in Real-World Clinical Oncology Databases in the US: Flatiron Health, SEER, and NPCR

**DOI:** 10.1101/2020.03.16.20037143

**Authors:** Xinran Ma, Lura Long, Sharon Moon, Blythe J.S. Adamson, Shrujal S. Baxi

**Affiliations:** Flatiron Health, Inc., New York, NY

**Keywords:** cancer, real-world data, cancer registries

## Abstract

**Background and Objective:** The Surveillance, Epidemiology, and End Results Program (SEER) program and the National Program of Cancer Registries (NPCR), are authoritative sources for population cancer surveillance and research in the US. An increasing number of recent oncology studies are based on the electronic health record (EHR)-derived de-identified databases created and maintained by Flatiron Health. This report describes the differences in the originating sources and data development processes, and compares baseline demographic characteristics in the cancer-specific databases from Flatiron Health, SEER, and NPCR, to facilitate interpretation of research findings based on these sources.

**Methods:** Patients with documented care from January 1, 2011 through May 31, 2019 in a series of EHR-derived Flatiron Health de-identified databases covering multiple tumor types were included. SEER incidence data (obtained from the SEER 18 database) and NPCR incidence data (obtained from the US Cancer Statistics public use database) for malignant cases diagnosed from January 1, 2011 to December 31, 2016 were included. Comparisons of demographic variables were performed across all disease-specific databases, for all patients and for the subset diagnosed with advanced-stage disease.

**Results:** As of May 2019, a total of 201,570 patients with 19 different cancer types were included in Flatiron Health datasets. In an overall comparison to national cancer registries, patients in the Flatiron Health databases had similar sex, age at initial diagnosis, and geographic distributions but appeared to be diagnosed with later stages of disease compared with patients in other datasets. For variables such as stage and race, Flatiron Health databases had a greater degree of incompleteness. There are variations in these trends by cancer types.

**Conclusions:** These three databases present general similarities in demographic and geographic distribution, but there are overarching differences across the populations they cover. Differences in data sourcing (medical oncology EHRs vs cancer registries), and disparities in sampling approaches and rules of data acquisition may explain some of these divergences. Furthermore, unlike the steady information flow entered into registries, the availability of medical oncology EHR-derived information reflects the extent of involvement of medical oncology clinics at different points in the specialty management of individual diseases, resulting in inter-disease variability. These differences should be considered when interpreting study results obtained with these databases.

## INTRODUCTION

The field of oncology is undergoing rapid evolution as our understanding of the pathophysiology of cancer expands and therapeutic progress accelerates. This environment calls for the development of tools that facilitate the translation of these advances into improvements in patient care. Recent years have seen increasing use of real-world data (RWD) as a clinical research source, from descriptive epidemiology to intervention effectiveness studies. RWD analyses generate real-world evidence (RWE) that could supplement and complement the evidence for new drug approvals (traditionally gathered from prospective studies), for health services research, policy evaluation, or as a pharmacovigilance tool (1,2).

RWD can be obtained from many different sources, including billing and administrative claim activities, product and disease registries, national surveys, and electronic health records (EHRs). The appropriate and optimal source of RWD will differ by the research question.

Traditionally, registries have been a key RWD source for epidemiologic and population-based outcomes studies; in the US, the Surveillance, Epidemiology, and End Results program (SEER) registries and the National Program of Cancer Registries (NPCR), have been commonly used for oncology research. The SEER Program (3), supported by the Surveillance Research Program (SRP) in the National Cancer Institute’s (NCI) Division of Cancer Control and Population Sciences (DCCPS), collects data on patient demographics, primary tumor site, morphology and stage at diagnosis, and first course of treatment; the SEER program provides cancer incidence and survival data through 16 state level population-based cancer registries currently covering approximately 34.6% of the US population. The NPCR (4) is supported by the US Centers for Disease Control and Prevention (CDC) Division of Cancer Prevention and Control, covers 97% of the US cancer population, spans 46 states, the District of Columbia, Puerto Rico, the US Pacific Island Jurisdictions, and the US Virgin Islands.

Following the passage of the Health Information Technology for Economic and Clinical Health Act (HITECH) (5) in January 2009, EHR systems have been rapidly adopted in the US and are now a key source of RWD. In the field of oncology, adoption has been even swifter than in other fields, and by 2015, approximately 90% of oncology practices had already adopted EHRs (6, 7). This, accompanied by an exponential growth of data storage and mining technology, has allowed for access to detailed clinical data at an unprecedented scale. In oncology, patient-level data can be derived from EHRs to generate granular information about baseline population characteristics as well as longitudinal views of sequential treatments, interventions, and associated outcomes. Flatiron Health is an oncology-focused health technology company that generates RWD from two EHR-derived primary sources: (i) OncoEMR^Ò^, a proprietary oncology-specific EHR used by community oncologists throughout the US, and (ii) EHR data integrations with academic research centers that enable bidirectional transmission of RWD.

The underlying data collection procedures and data model architecture make Flatiron Health a fundamentally different data source from SEER or NPCR (Table 1). SEER and NPCR have been well established as research resources since their inception in 1973 and 1992, respectively. As the most recent of the three sources, data derived from Flatiron Health have become a research resource in the last five years (8–18); therefore, it has become increasingly critical to understand their features. SEER and NPCR collect specific incident disease data points in a systematic and ordered fashion, fulfilling a public health reporting mandate justified by the public health burden of cancer as a disease. Flatiron Health data collection mirrors routine oncology EHR documentation practices; upon curation, this approach yields longitudinal clinical data models with considerable depth, including clinical, genetic, and outcome data. This feature, together with its 30-day recency, makes this a suitable source for detailed investigation of contemporary trends in cancer management, including sequential time-to-event endpoints.

**Figure 1.**
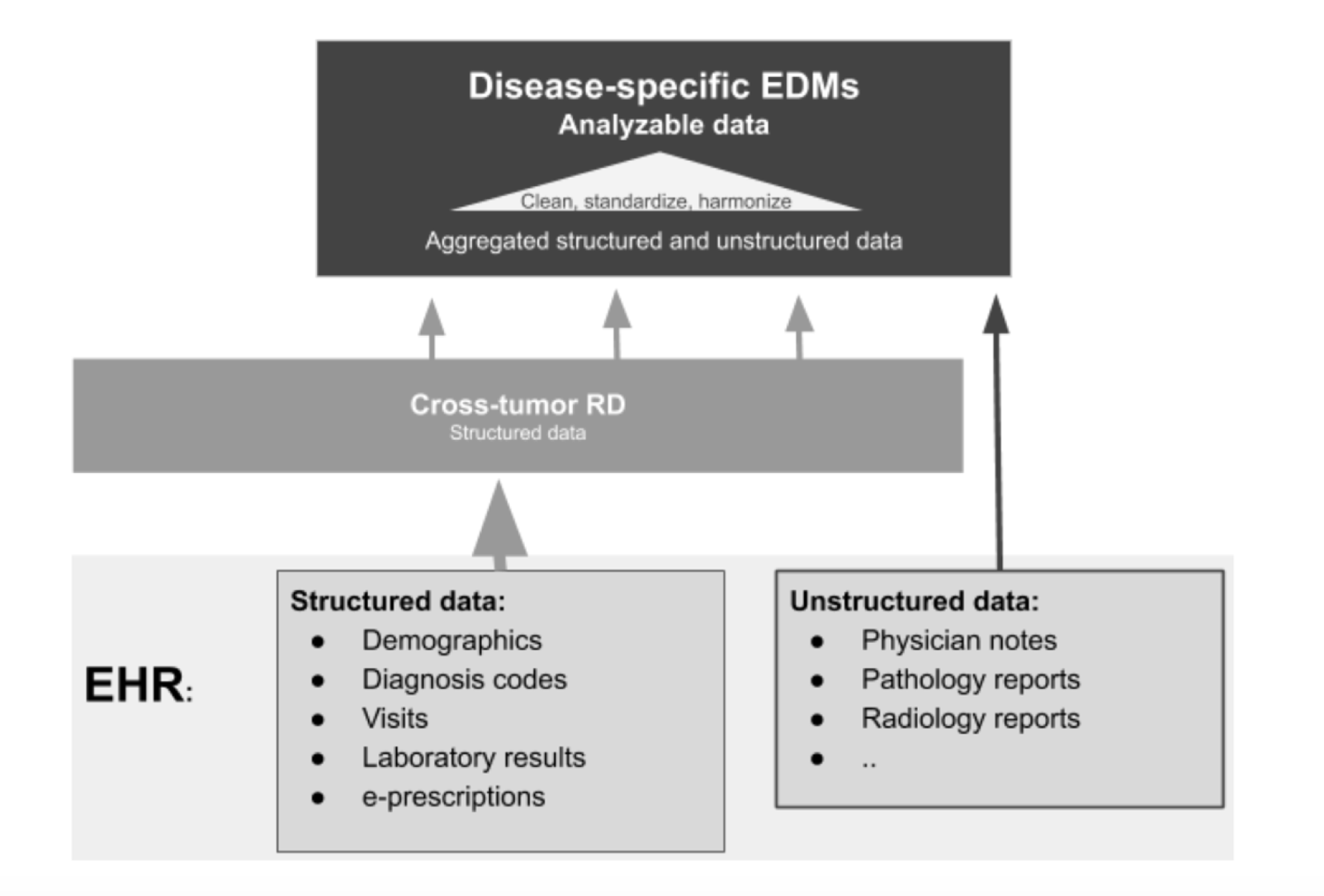
Schema of the structure of the Flatiron Health databases. RD=research database; EDM=enhanced data mart

**Table 1.** General features of SEER, NPCR and Flatiron Health databases.

| Feature | SEER | NPCR | Flatiron Health |
| --- | --- | --- | --- |
| Source | State disease registries (3) receive information about cancer diagnoses and treatment from public health reporting entities (including healthcare organizations, facilities and providers).<br>Geographic distribution of participating states is based on maintaining a high-quality population-based cancer reporting system, and pre-specified and adjusted to reach specific regional and racial representation.<br>Mortality information is obtained from the CDC's National Center for Health Statistics' National Vital Statistics System (20), which collects mortality data along with cause-specific death classification information. | 46 state and D.C. disease registries, receive information about cancer diagnoses from healthcare providers from any specialty.]<br>All-cause mortality is assessed from the CDC's National Center for Health Statistics' National Vital Statistics System (20). | EHRs from community and academic oncology clinics participating in the Flatiron network. Databases receive routinely collected information (i.e., without intentional or pre-specified collection) from non-oncology care settings or sites outside of participating clinics via scanned in documentation.<br>Mortality is assessed via a composite variable that integrates multiple data sources (EHR information, a commercial death data source, and the Social Security Death Index) and is benchmarked against the National Death Index (NDI). |
| Collection approach | Data collection is based in SEER and NAACCR data standards, and is intentional in order to meet pre- | Data collection is intentional in order to meet pre-specified registry program standards for | Originating information is collected as routinely documented or entered by participating practitioners. |
|  | specified registry program standards for completeness and quality. | completeness. |  |
| Curation | Unstructured and structured data are abstracted by tumor registrars, who enter text manually into a data collection template.<br>In order to facilitate pooling of data, all SEER and NPCR cancer registries use uniform data items and codes as documented by the North American Association of Central Cancer Registries (3,34). | Unstructured and structured data are abstracted by tumor registrars, who enter text manually into a data collection template. Some pathology reports are received electronically.<br>In order to facilitate pooling of data, all NPCR and SEER cancer registries use uniform data items and codes as documented by the North American Association of Central Cancer Registries (3,34). | Structured data are harmonized and mapped to common units and terminology and combined with unstructured data processed by technology-enabled manual abstraction. Abstractors (clinical oncology nurses and tumor registrars) are trained to identify and extract relevant information by following optimized policies and procedures, and oversight is provided by medical oncologists. |
| Recency | Releases data with a 2-year delay | Releases data with a 2-year delay | Data are available with a 1-month delay |
| **Key data model elements** |  |  |  |
| **Demographics<sup>a</sup>** | Age, sex, race, ethnicity, insurance type, geographic location | Age, sex, race, ethnicity, geographic location | Age, gender,<sup>b</sup> race, insurance type, geographic location |
| **Tumor details** | Date of initial diagnosis, primary tumor site, limited biomarker information (34,35),<sup>c</sup> morphology/histology, behavior, laterality and stage at diagnosis | Date of initial diagnosis, primary tumor site, limited biomarker information,<sup>c</sup> a morphology/histology and stage at diagnosis | Date of initial diagnosis, primary tumor site, morphology/histology and stage at diagnosis, standard biomarker information,<sup>c</sup> date of advanced/metastatic diagnosis, sites of metastatic disease<sup>d</sup> |
| **Treatment** | First course of treatment at initial diagnosis:<br>• Diagnostic/procedure<br>• Surgery<br>• Radiation<br>• Systemic therapy (chemotherapy, hormone therapy, biological response modifier [immunotherapy], or other) | First course of treatment at initial diagnosis:<br>• Surgery | Sequence of treatments received after advanced/metastatic disease:<sup>e</sup><br>• Surgery, limited to the documentation in the oncology EHR, in certain diseases.<br>• Radiation, limited to the documentation in the oncology EHR, in certain diseases.<br>• Systemic therapy: type, dosing, dates of individual drug episodes (e.g. ordered dates, administered dates) including abstracted oral therapies |
| **Variables** | Cause specific mortality | Mortality | Mortality (19)<br>Real world progression (rwP) (36). |

<sup>a</sup>In the May 2023 update of this paper, “ethnicity” was removed as a data model element from Flatiron Health for clarity, as race values that were documented as ethnicity (“Hispanic or Latino”) were recoded as “Unknown,” and ethnicity was not separately reported in the results. This change is also reflected in Tables 2-20 and Table A2.
<sup>b</sup>Gender values likely reflect a patient’s sex assigned at birth. However, it is unknown whether these values represent sex assigned at birth or gender identity or whether these values are self- or clinician-reported. These values vary by practices at individual clinical sites and the limitations of the EHR software used.
<sup>c</sup>The biomarker portfolio offered by each database is different, with SEER providing information about a limited set of specific biomarkers that might predict outcomes or response to specific therapies, the US Cancer Statistics database providing ER, PR and HER2 status, and Flatiron Health offering a more extensive portfolio of biomarkers for which testing is considered standard of care.
<sup>d</sup>Flatiron Health databases provide information on specific sites of metastatic disease for a selected set of tumor types.
<sup>e</sup>Flatiron Health has assembled early-stage disease datasets (for which information about early-stage treatment is collected) for certain selected tumor types.

Researchers considering Flatiron Health data for their studies may be interested in gaining insights into the nature of the originating source, on the principles of data collection, and on the characteristics of its patient population. Descriptive statistics of the Flatiron Health, SEER, and NPCR data can be informative for the interpretation of findings when these large data sets are used in cancer research. This study aims to describe data sources and collection procedures, and to provide a detailed comparison of the demographic characteristics in the disease-specific databases from Flatiron Health, SEER, and NPCR, identifying similarities and differences within the data elements common across the three.

## METHODS

### Flatiron Health databases

Flatiron Health has developed de-identified disease-specific dynamic databases (termed enhanced data marts) derived from information available in an EHR. These databases combine curated manually-abstracted unstructured data with structured data. The starting point is a single database composed only of structured data elements available within an EHR (termed the Flatiron Health research database) refreshed on a monthly basis. This large general cross-tumor cohort includes all patients with at least one International Classification of Diseases (ICD)-9 or ICD-10 cancer code and at least one unique-date clinic encounter documented in the EHR (reflected by records of vital signs, treatment administration, and/or laboratory tests) on or after January 1, 2011, from both academic and community care sites combined. From there, patient data are sampled each month into each disease-specific database with randomized approaches implemented through software code, to ensure uniform application of the sampling approach and to avoid the potential for bias. Inclusion is based on cancer-specific cohort inclusion and exclusion criteria (i.e., relevant ICD-9 and ICD-10 codes and technology-assisted abstraction of unstructured information, such as metastatic status).

All manual abstraction of unstructured information, including confirmation of diagnosis and stage, is carried out by abstractors (i.e., clinical oncology nurses or tumor registrars). Clinically-relevant details specific to each cancer type are abstracted from any form of clinical documentation available in the EHR including clinic visit notes, radiology reports, pathology reports, etc. Abstractors are trained to identify and extract relevant information by following policies and procedures tested and optimized for reliability and reproducibility through iterative processes, and oversight is provided by medical oncologists. Each month, the datasets grow with new cases as well as incremental abstraction of newly-available clinical documentation from pre-existing patients. Therefore, at any given cutoff time, the size of a dataset is dependent on the initiation date for that disease-specific dataset and the overall population prevalence (which determines the growth rate). Typical data recency is 30 days. Practically speaking, a data cutoff of December 31, 2019 would include all information entered into the EHR through November 30, 2019, and subsequent cutoffs (i.e., January 31, 2020, February 29, 2020, etc) will render increasingly larger sample sizes. In addition, each database undergoes continuous audit procedures to monitor abstractor performance while proprietary technology links each curated data variable to its source documentation within the EHR, enabling subsequent review, when necessary. At the individual patient level, this approach provides a recent and robust longitudinal view into the clinical course, capturing new clinical information as it is documented within the EHR. Flatiron Health data are available for research via Institutional review board (IRB) approval of a master study protocol with waiver of informed consent (IRB # RWE-001, “The Flatiron Health Real-World Evidence Parent Protocol”, Tracking # FLI1-18-044 by the Copernicus Group IRB), obtained prior to study conduct, which covers the data from all sites represented.

As of May 31, 2019, there were 19 disease-specific databases available at Flatiron Health: advanced urothelial cancer, metastatic breast cancer, early breast cancer, chronic lymphocytic leukemia (CLL), metastatic colorectal cancer, diffuse large B-cell lymphoma (DLBCL), follicular lymphoma (FL), advanced gastric/esophageal carcinoma, advanced hepatocellular carcinoma (HCC), advanced head and neck cancer, advanced melanoma, malignant pleural mesothelioma, multiple myeloma (MM), advanced non-small cell lung cancer (NSCLC), ovarian carcinoma, metastatic pancreatic carcinoma, metastatic prostate cancer, advanced renal-cell carcinoma (RCC), and small-cell lung cancer (SCLC) (Appendix I). At least two unique-date clinic encounters documented in the EHR in the Flatiron Health database (reflected by records of vital signs, treatment administration, and/or laboratory tests) on or after January 1, 2011, are required for patient data to be entered into a given dataset, with the exception of a few diseases with different start dates (Appendix I).

At the time of this analysis, the Flatiron Health EHR-derived database included de-identified data from over 280 cancer practices representing more than 2.2 million patients and about 800 distinct sites of care from all 50 states and Puerto Rico. The distribution of patients across community and academic practices largely reflects patterns of care in the US, where most patients are treated in community clinics, but can vary for each disease. Mortality information is captured via a composite variable that uses multiple data sources (structured and unstructured EHR content, commercial sources, Social Security Death Index) and is benchmarked against the National Death Index data as a gold standard (19).

### SEER and NPCR databases

The SEER Program supports most aspects of cancer surveillance research, providing analytical tools, and methodological expertise in collecting, analyzing, interpreting and disseminating population-based statistics. SEER population-based data include cancer incidence and survival data by age, sex, race, year of diagnosis, and geographic areas (including SEER registry and county). SEER releases new research data each spring based on the previous November’s data submission. Data are available across various registries and versions of the SEER Program from 1975 through 2016. Mortality information in SEER is obtained from the CDC’s National Center for Health Statistics’ National Vital Statistics System (20) and includes mortality data along with cause-specific death classification information.

The NPCR cancer registries routinely capture data elements including the type, extent, and location of the cancer, the type of initial treatment, and outcomes of newly diagnosed cancers. Medical facilities such as hospitals, physician offices, and pathology laboratories send information about cancer cases to their respective central cancer registry, and each central cancer registry submits electronically de-identified demographic and clinical information to the NPCR on a yearly basis (4). Mortality information in the NPCR is obtained from the CDC’s National Center for Health Statistics’ National Vital Statistics System (20). As of May 31, 2019, the most recent information available from NPCR included new incident malignancies diagnosed through December 31, 2016. NPCR data is made available through the US Cancer Statistics dataset, which combines NPCR data and data from 4 SEER-funded states (Connecticut, Hawaii, Iowa, and New Mexico). This data provides information on 100% of the US population. In this paper, “NPCR data” was obtained through the US Cancer Statistics public use research dataset and was restricted to the 46 NPCR funded states and D.C.

### Comparative analysis

#### Variables

For each cancer type, demographic and clinical characteristics including race, age, region, year and stage at diagnosis were compared between the Flatiron Health and the SEER and NPCR databases. To overcome coding discrepancies across databases, cancer types were matched using ICD-9, ICD-10, and histology codes (e.g. ICD-0-3). All comparisons were unadjusted. Variables not available across two or more data sources were not included in the comparison (e.g., smoking status, treatment detail, real world progression [rwP] information).

In the Flatiron Health databases, cancer staging information was collected as entered into the EHR by the treating physician or otherwise as assessed by Flatiron Health abstractors; during the study time period, the applicable staging criteria for solid tumors were those of the American Joint Commission of Cancer (AJCC) 7th edition and 8th edition manuals (21, 22), Rai staging for CLL and the International Staging System (ISS) for MM. For SEER and NPCR, diagnosis and staging information was abstracted from various sources including medical records and pathology reports. For both programs, staging information followed the Collaborative Stage coding systems (23). See additional information on variable definitions in Appendix I.

#### Patient eligibility and time frames

Diagnostic codes and eligibility criteria used to select the patients eligible for each database are listed in Appendix I. For SEER and NPCR, only malignant cases diagnosed on or after January 1st, 2011 were included for all the analyses (benign, uncertain behavior, carcinoma in situ, secondary malignancy cancers were excluded). For the Flatiron Health databases, patients diagnosed between January 1, 2011 and May 31, 2019, were included (except those missing diagnosis year and/or birth year).

In order to compare the particular data segments common across all three databases (the most recent SEER and NPCR data releases reach through 2016 as initial diagnosis year), descriptive analyses were performed not only for all patients available for analysis across the entire time frame of January 2011 - May 2019 in the Flatiron Health databases, but also in the subset available for analysis from January 2011 - December 2016.

As sensitivity analyses to address potential biases related to temporal drifts, we performed separate comparisons for the patient subgroups who had stage IV disease at diagnosis in each cancer type, for whom survival times would be expected to be shorter and the date of diagnosis would be expected to be closer to the database entry point. The potential biases to address were twofold: (i) as noted above, SEER and NPCR only collect specific incident disease data points, whereas Flatiron Health databases include both incident and prevalent cases. Therefore, Flatiron Health databases may receive patients at the time of diagnosis but also patients with initial diagnosis dates in the past; these cases may have long intervening periods between the initial diagnosis date and the date of entry into the Flatiron Health database, introducing a potential bias for patient characteristics associated with longer survival times (when compared with strictly incident cases in cancer registries); (ii) in addition, temporal trends where certain patient characteristics (i.e., sex, age) may be associated with cancer diagnoses during discrete time periods and can affect distributions depending on diagnosis year.

#### Analyses

Case-level data for patients in SEER were extracted from the SEER 18 November 2018 data submission dataset to the SEER Program by using the Case Listing Session feature in SEER*Stat software (Version 8.3.6, Information Management Services, Inc., Silver Spring, MD) and processed by using R 3.6.1. For patients in NPCR, case listing is not publicly available in the US Cancer Statistics public use SEER*Stat dataset, and case-level data cannot be accessed or downloaded. Frequencies by demographic and clinical characteristics for all malignant cases were calculated in SEER*Stat software using the November 2018 data submission.

The analysis of patients in the Flatiron Health databases was refreshed in April 2023 to incorporate an update to the birth year variable, reflecting best practices in patient de-identification (see Appendix II for more detail). This refresh resulted in updates to the distributions in calculated age at initial diagnosis in Tables 2 through 20 and A3.A through A3.S.

**Table 2.**
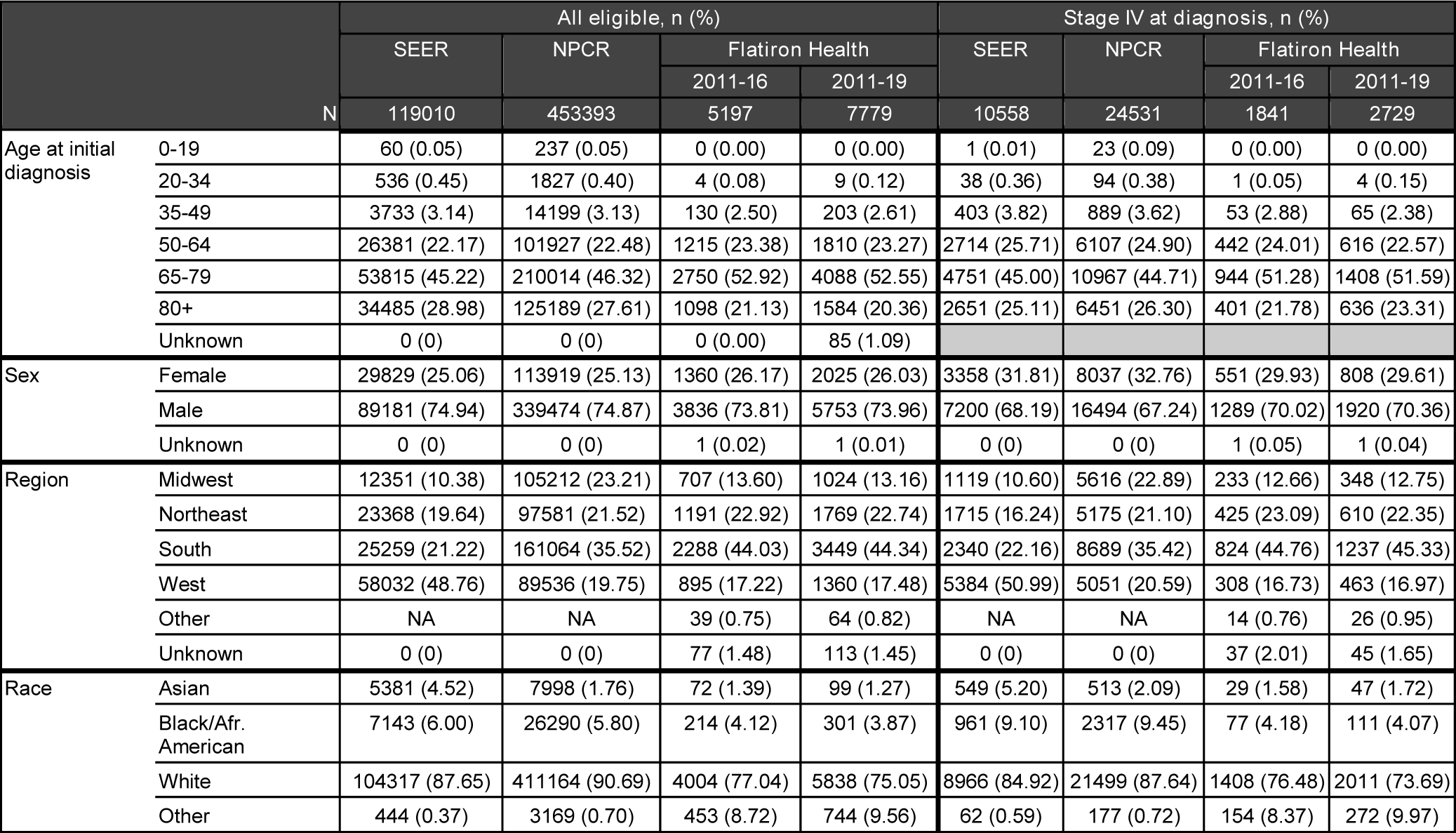
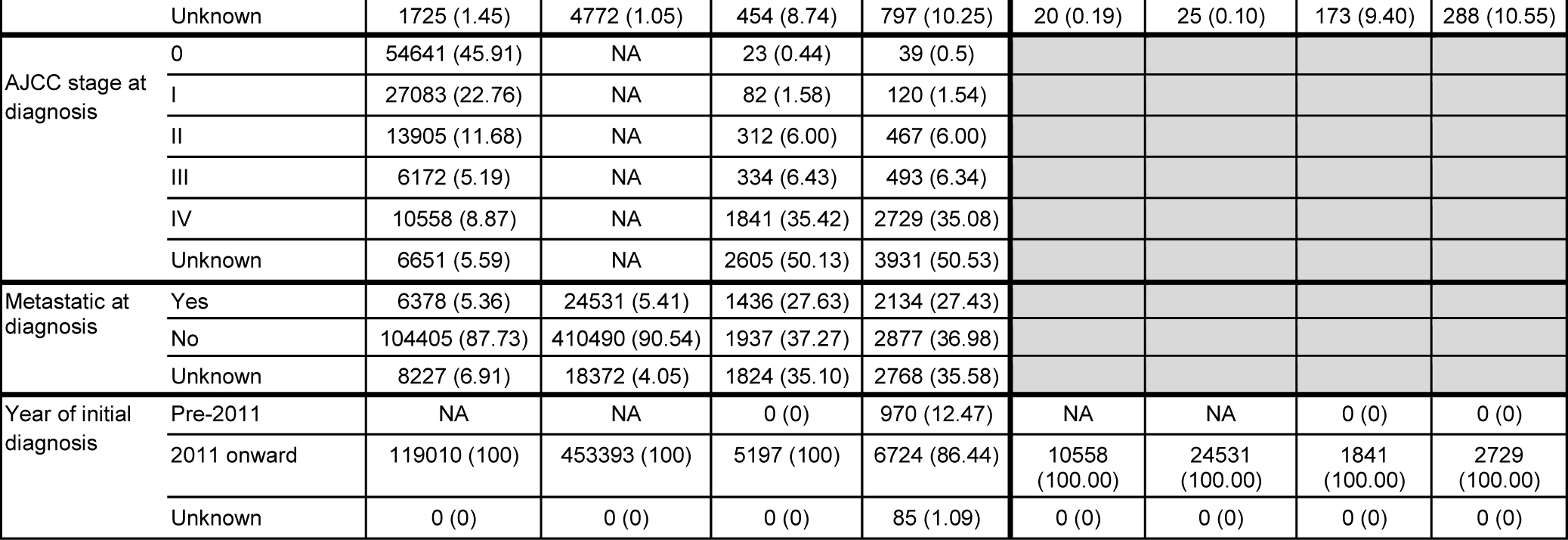
Characteristics for patients with bladder cancer. **Tables 2-20**: Shaded areas are groups with no reported results. NA = not available from the source data, or numbers under the reporting suppression value to preserve patient confidentiality

**Table 3.**
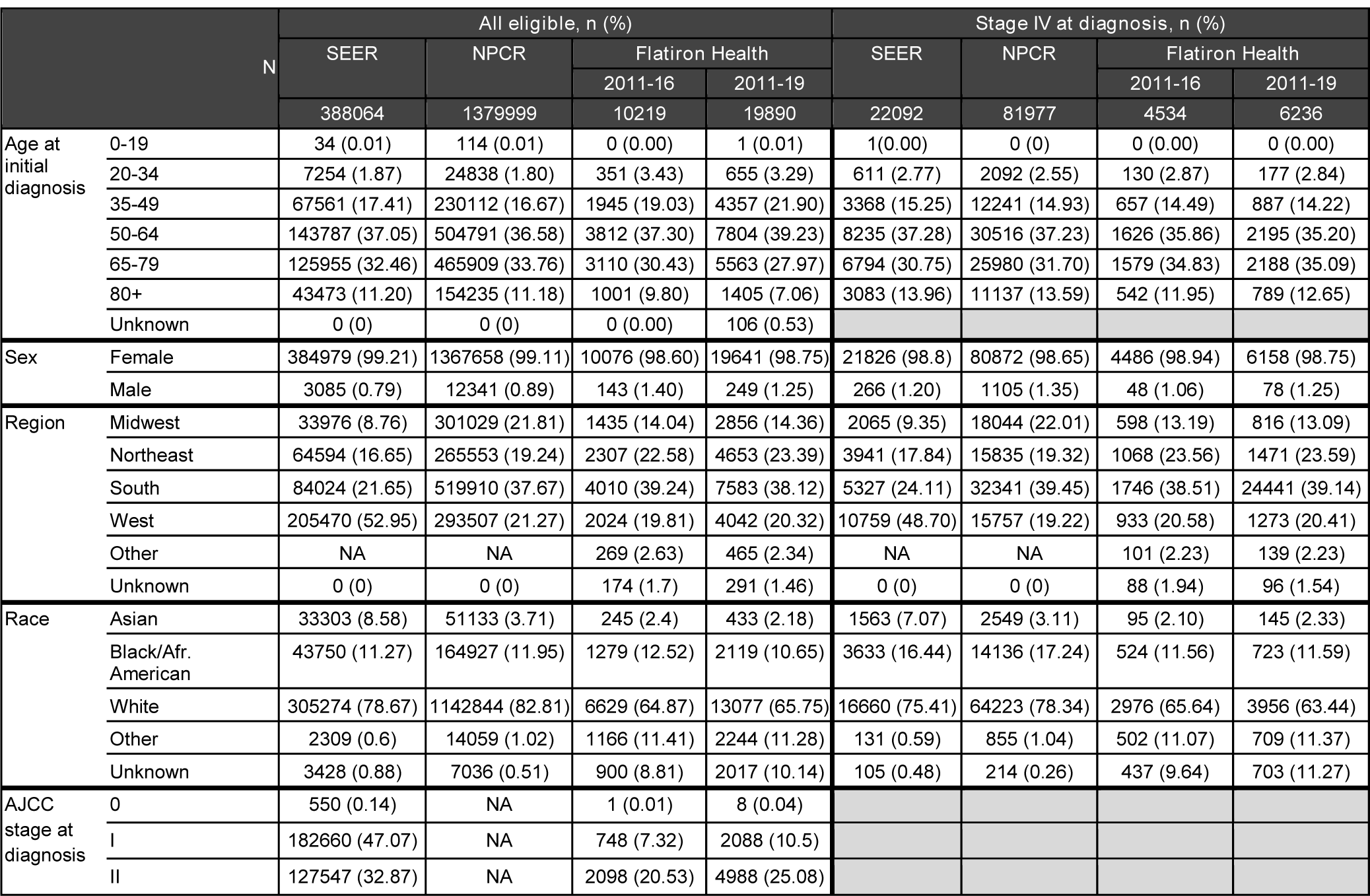
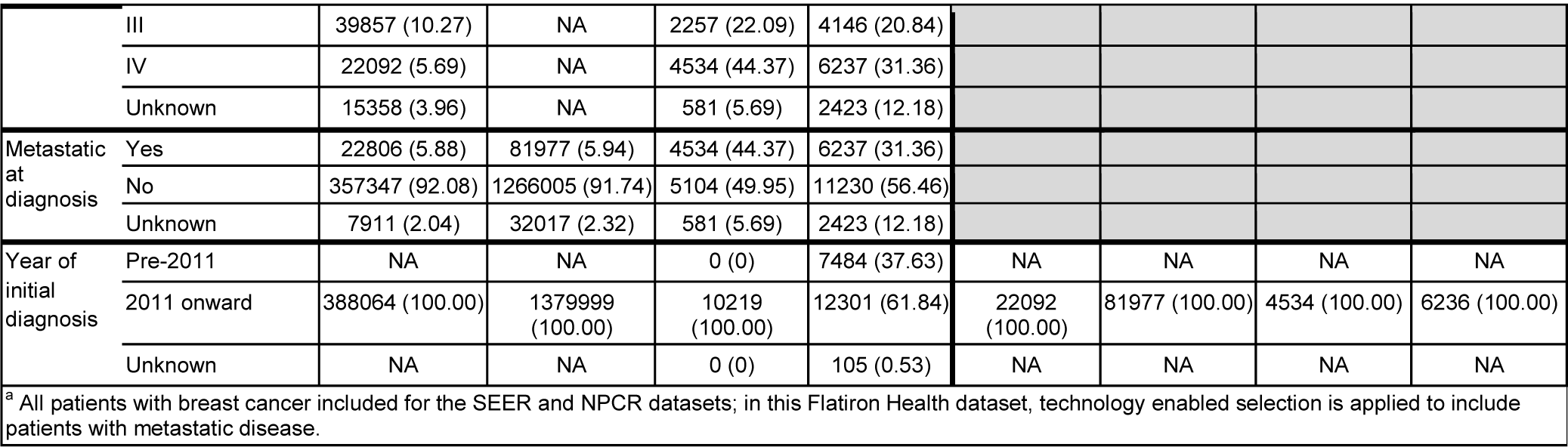
Characteristics for patients with metastatic breast cancer in the Flatiron Health database, compared with patients with breast cancer (any stage) in SEER and NPCR^a^

**Table 4.**
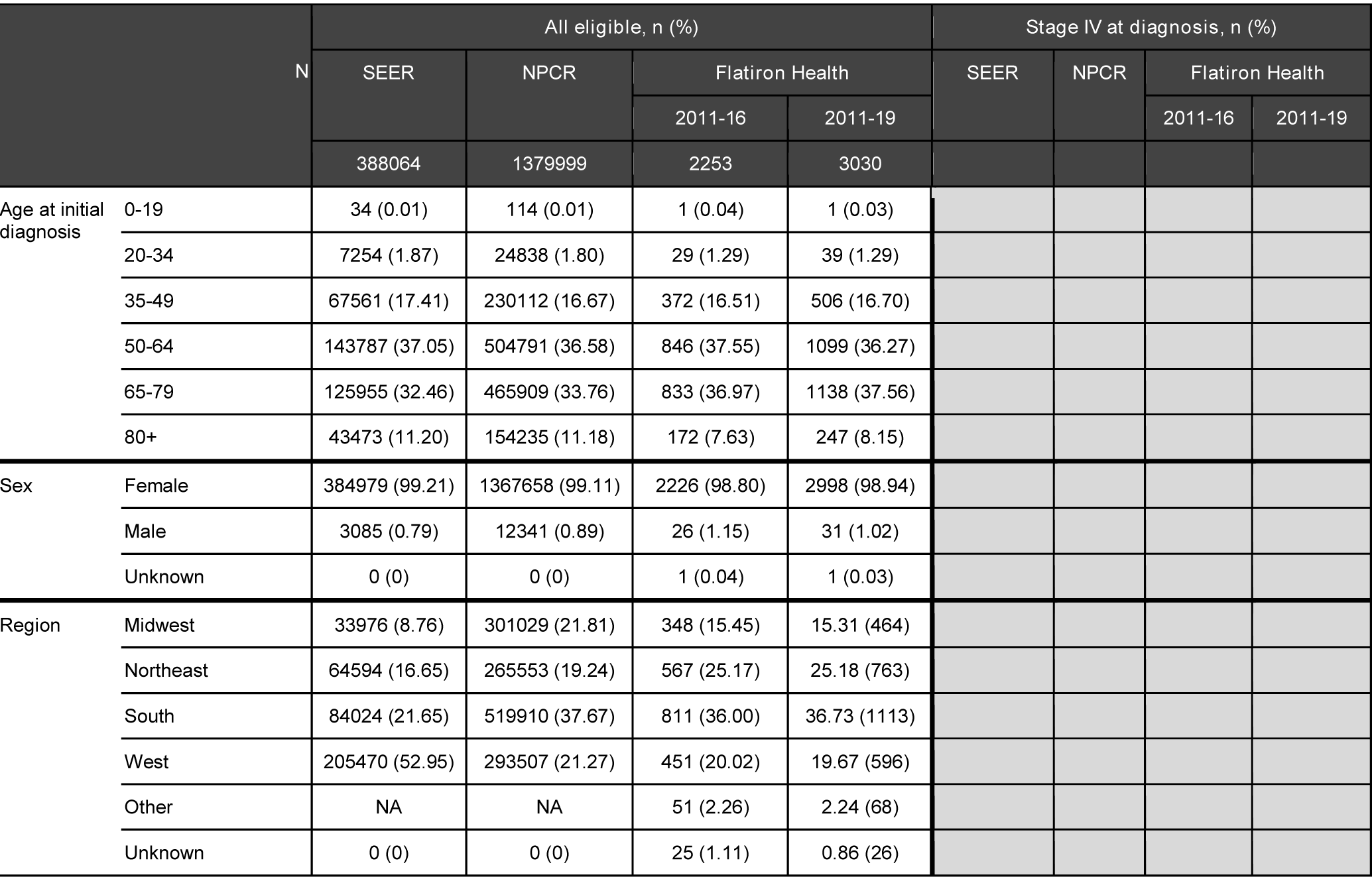
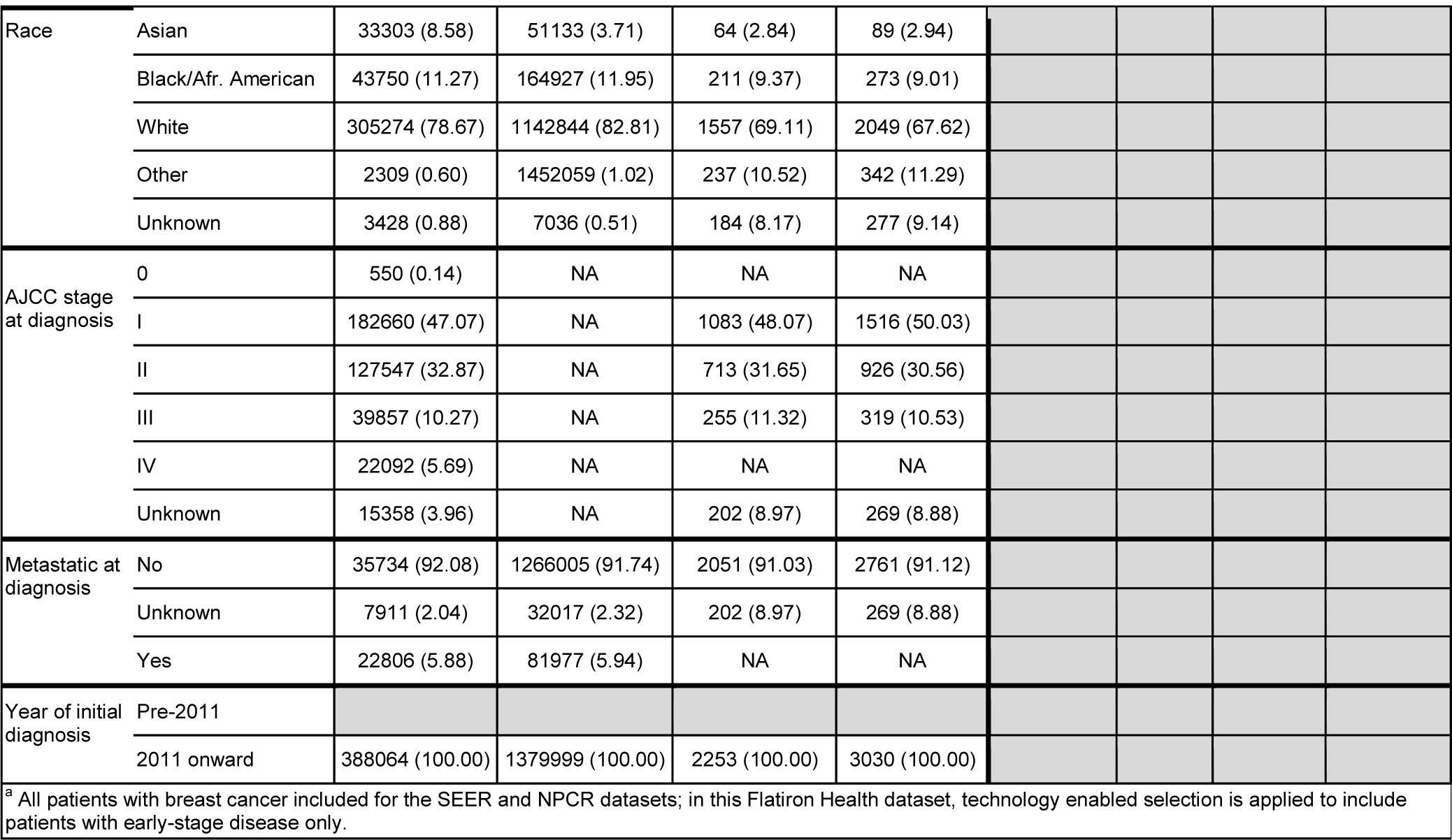
Characteristics for patients with early breast cancer in the Flatiron Health database, compared with patients with breast cancer (any stage) in SEER and NPCR^a^

**Table 5.**
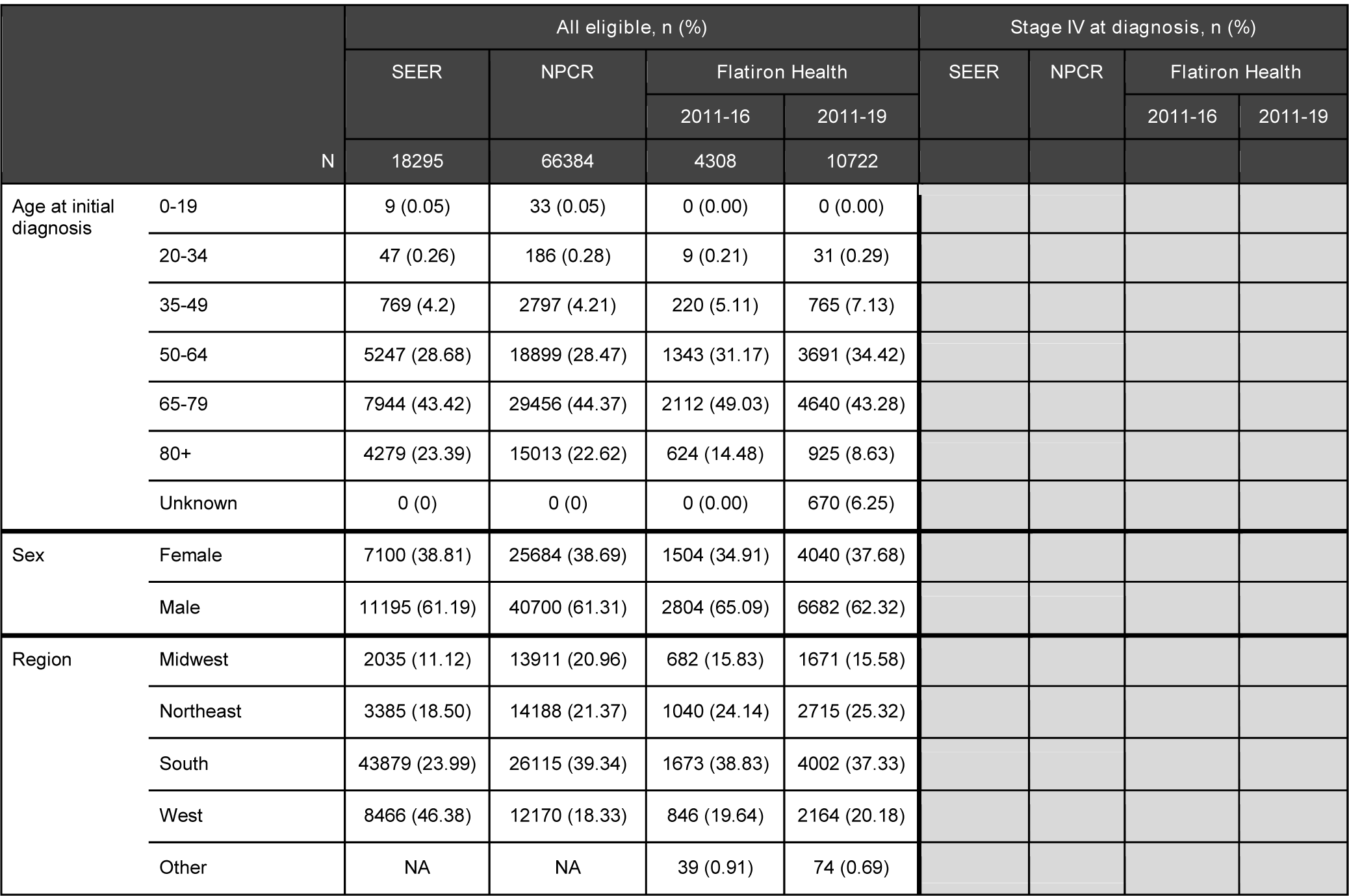
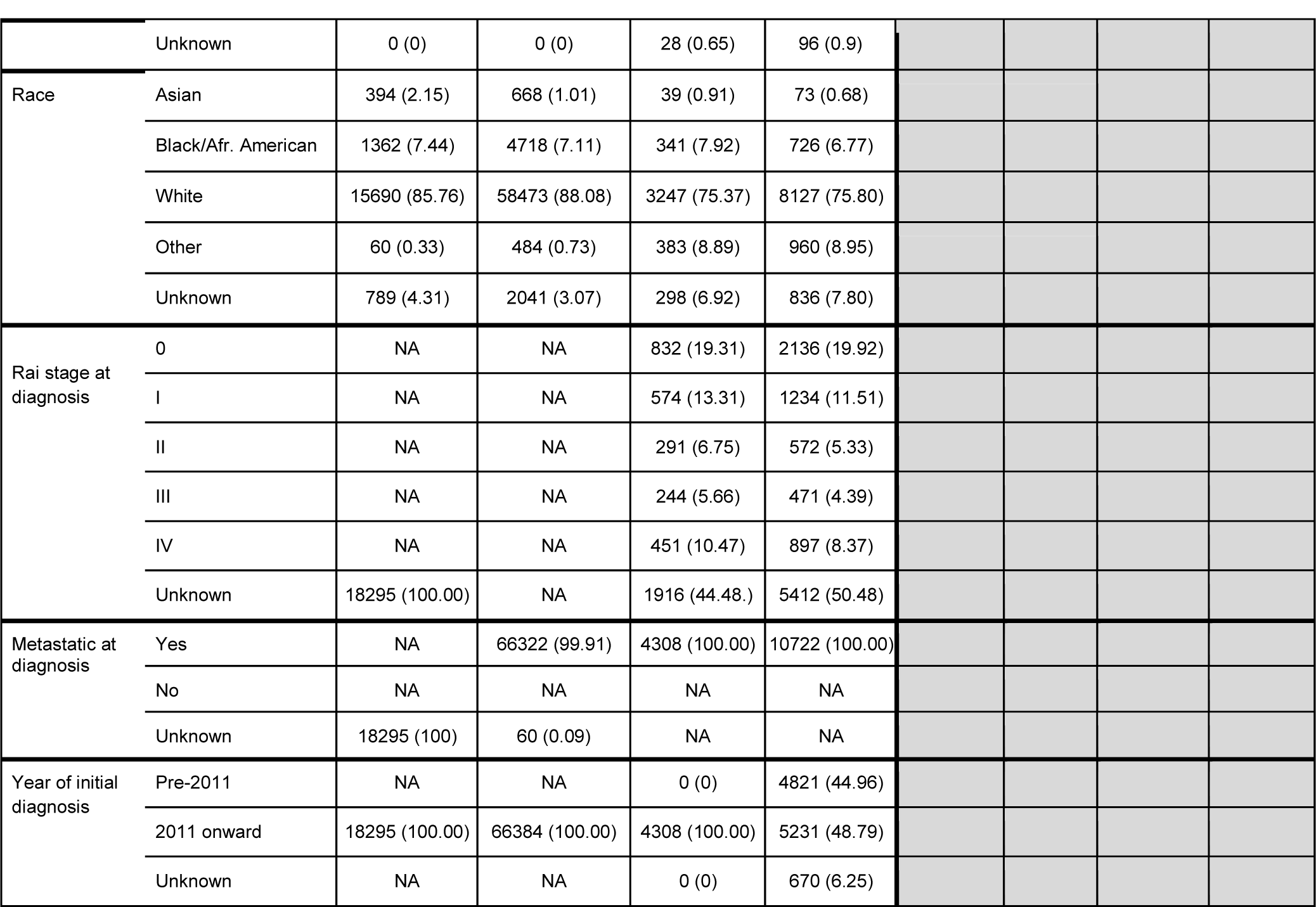
Characteristics for patients with chronic lymphocytic leukemia

**Table 6.**
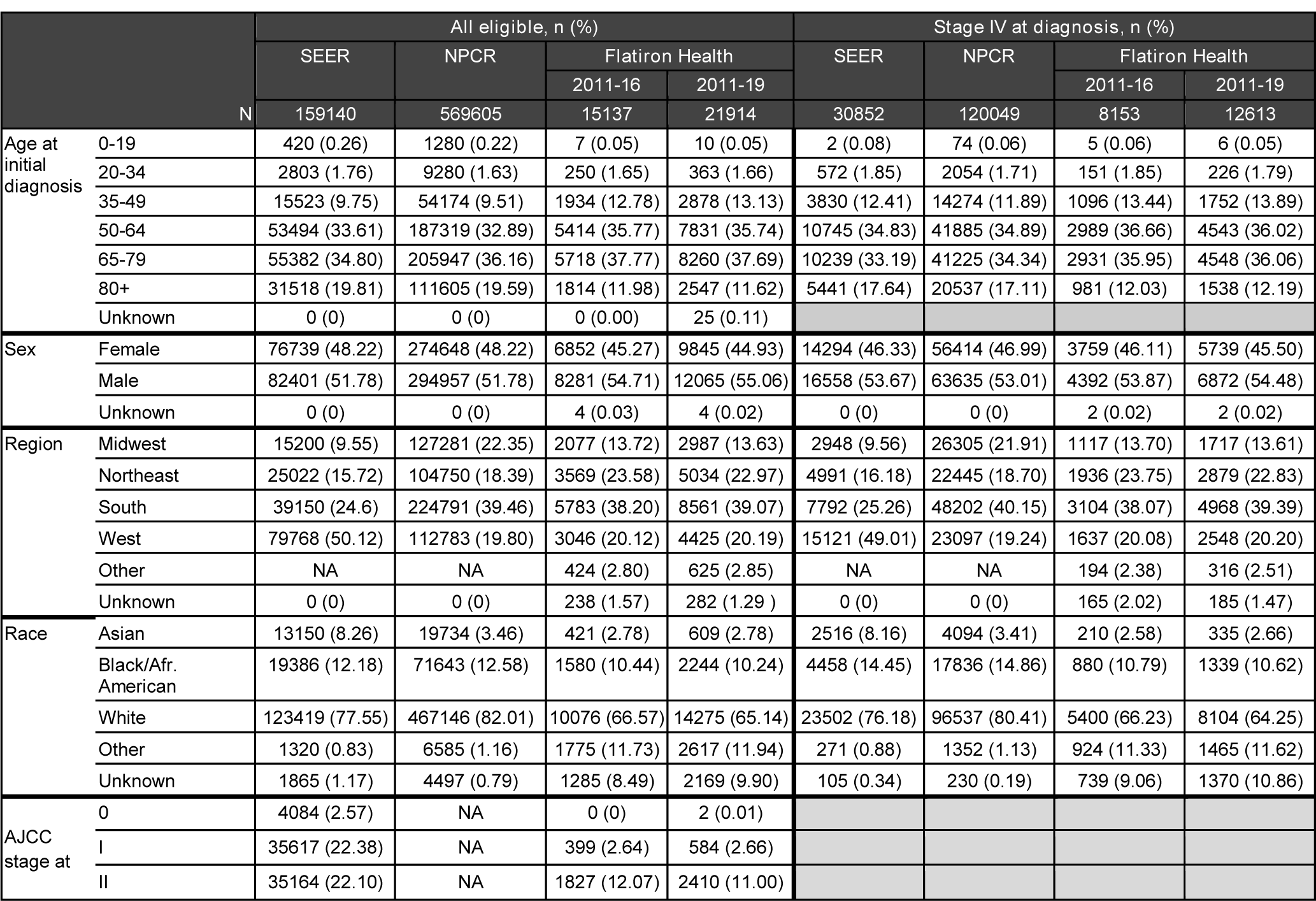
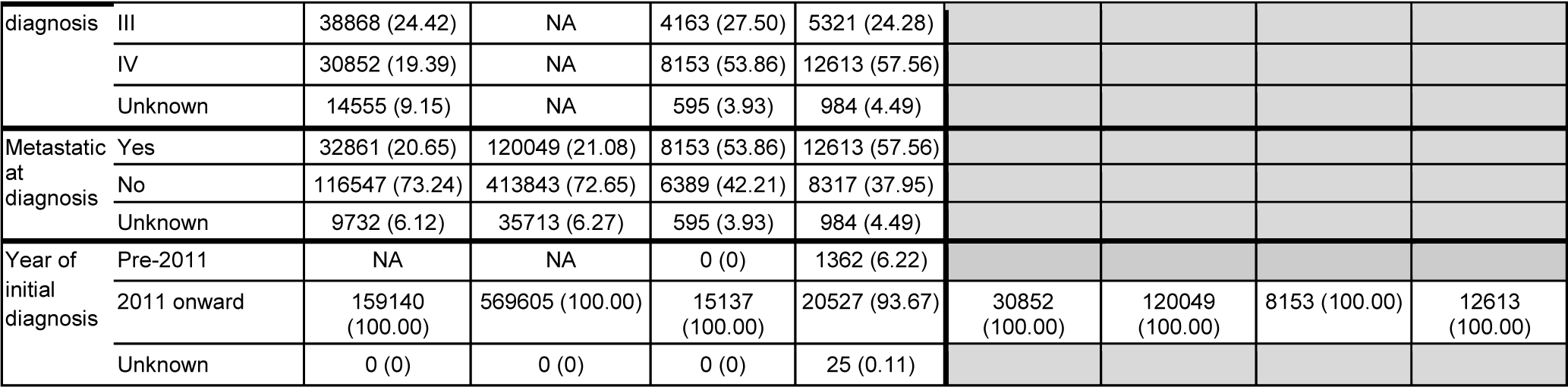
Characteristics for patients with colorectal cancer

**Table 7.**
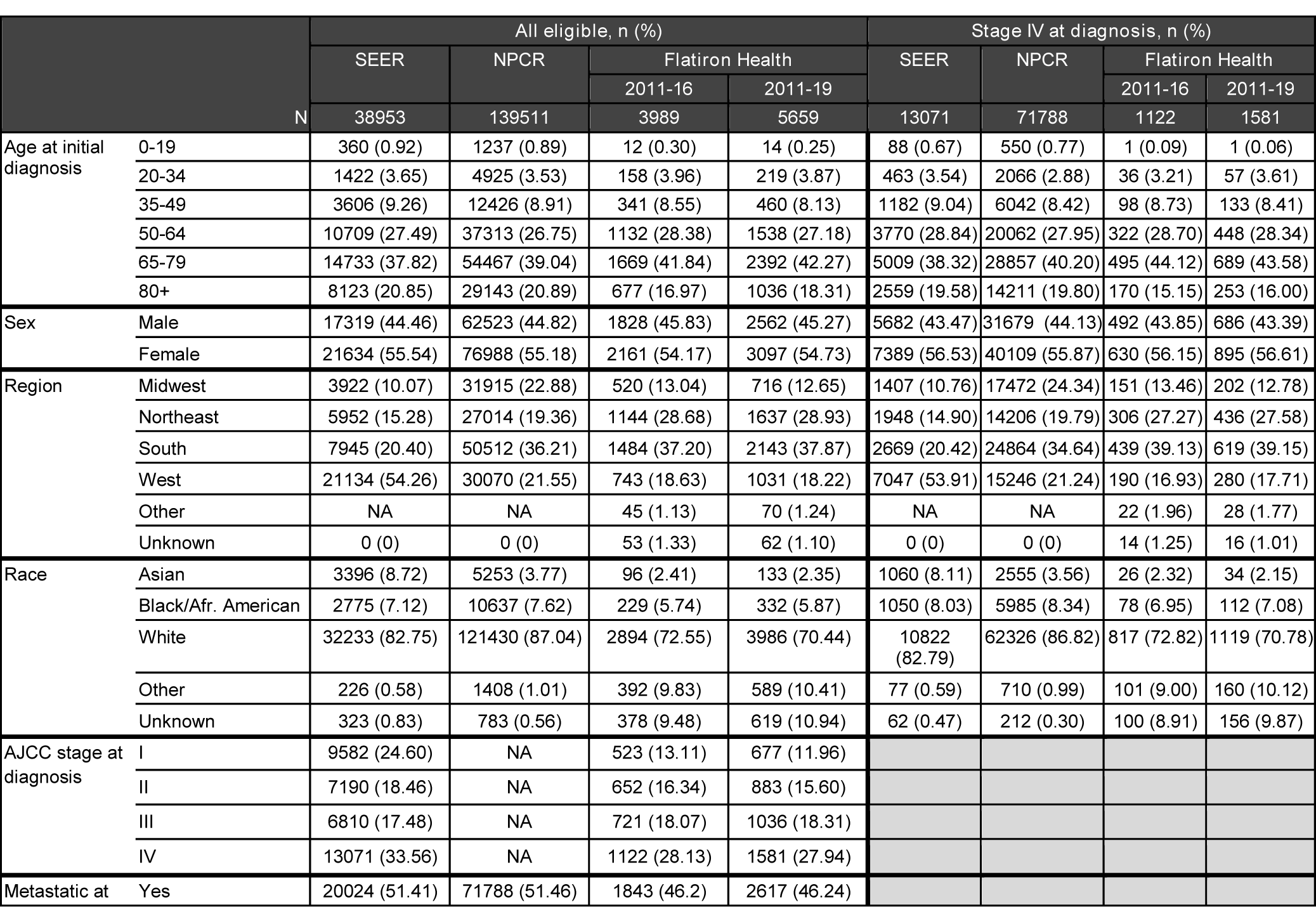
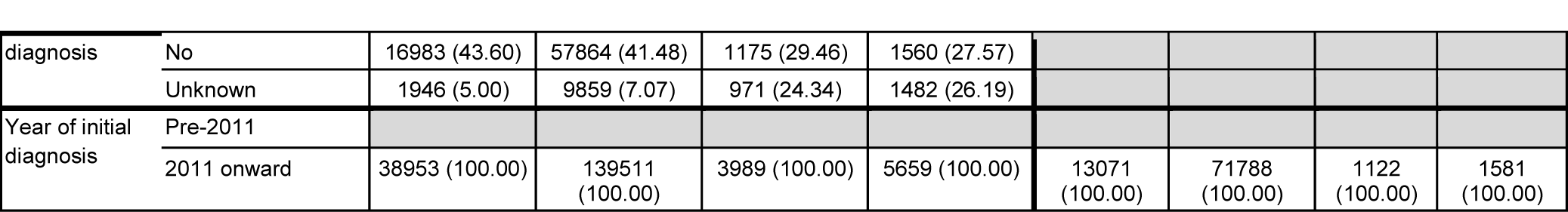
Characteristics for patients with diffuse large B-cell lymphoma

**Table 8.**
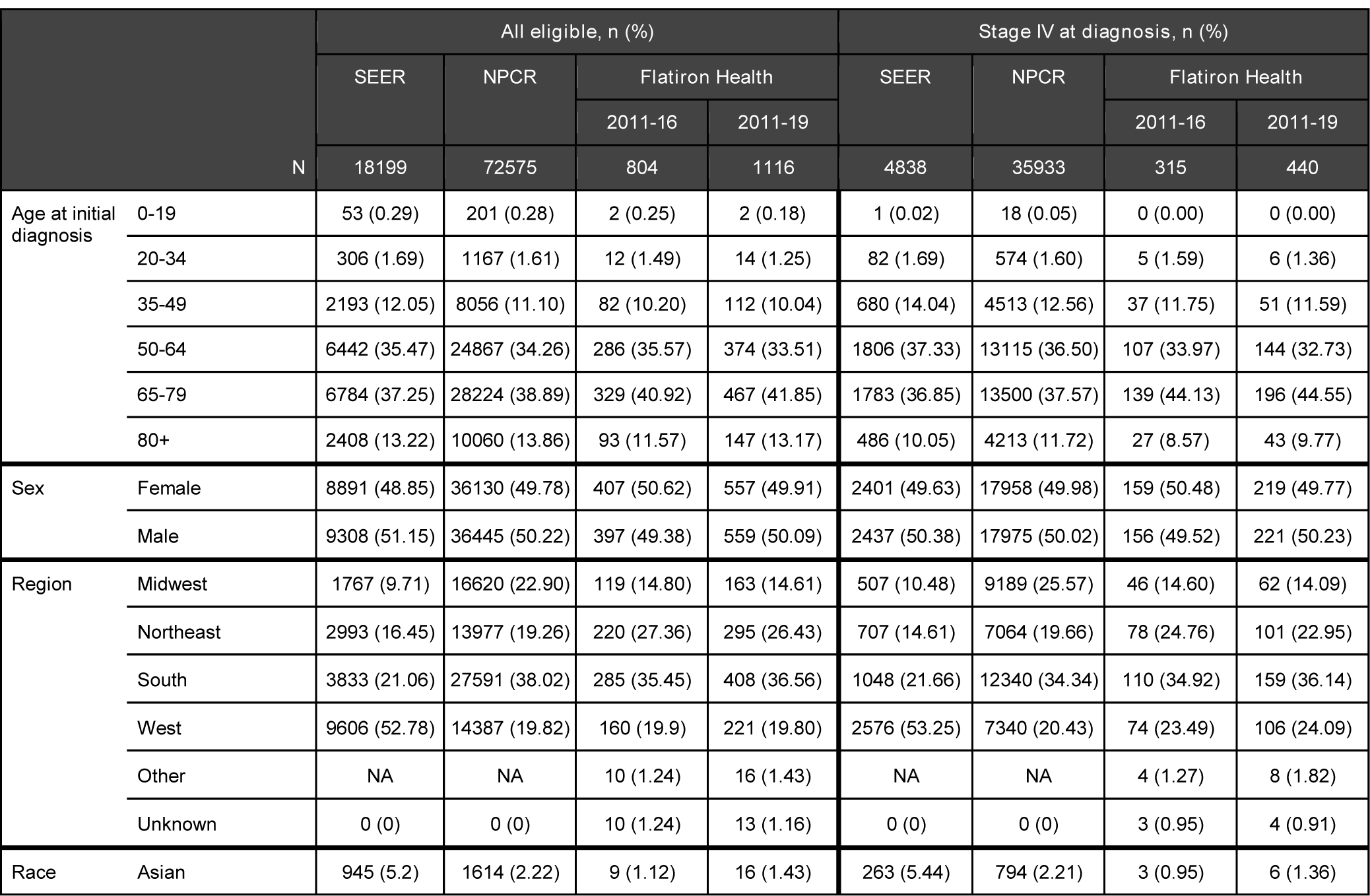
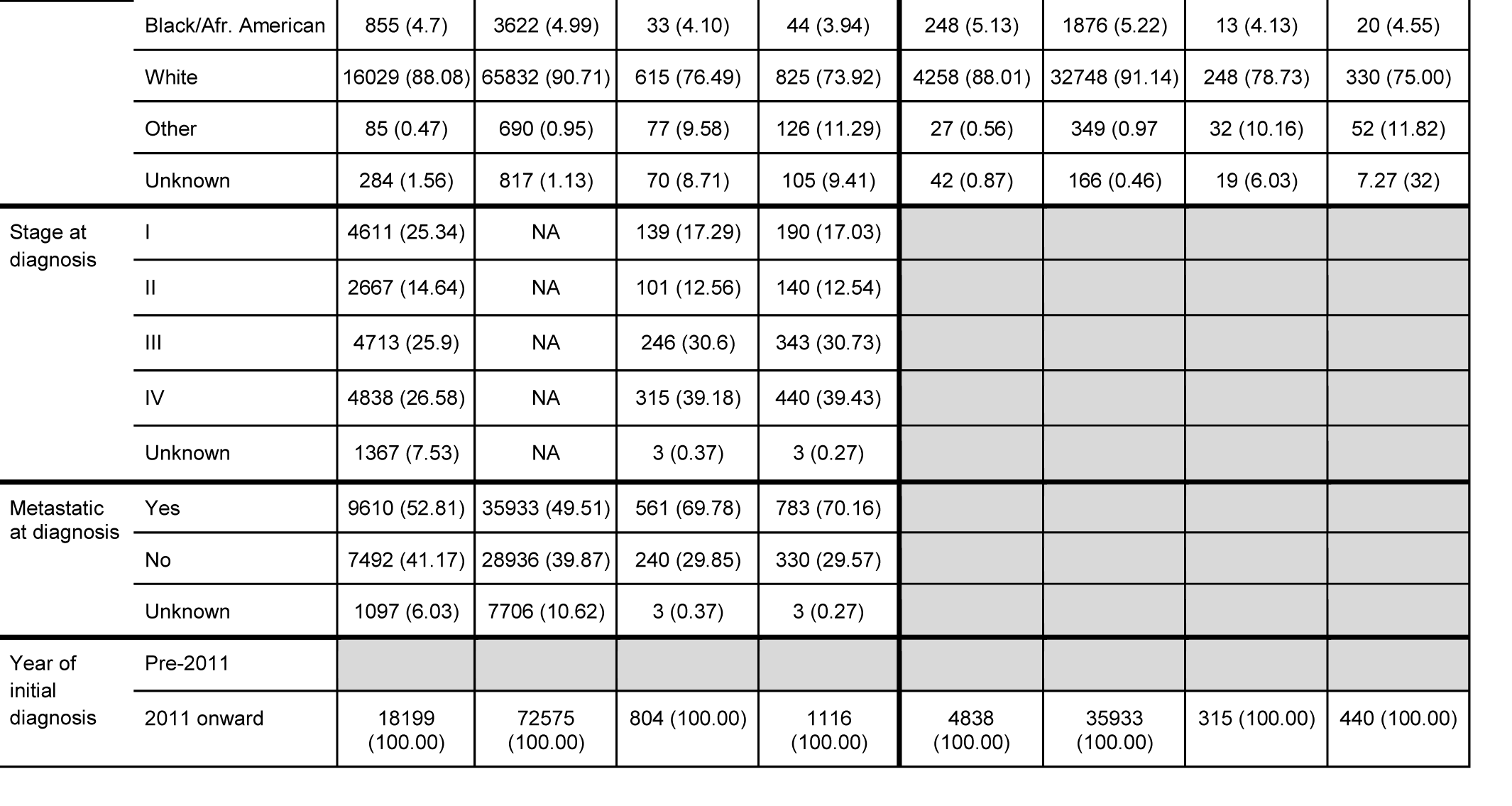
Characteristics for patients with follicular lymphoma

**Table 9.**
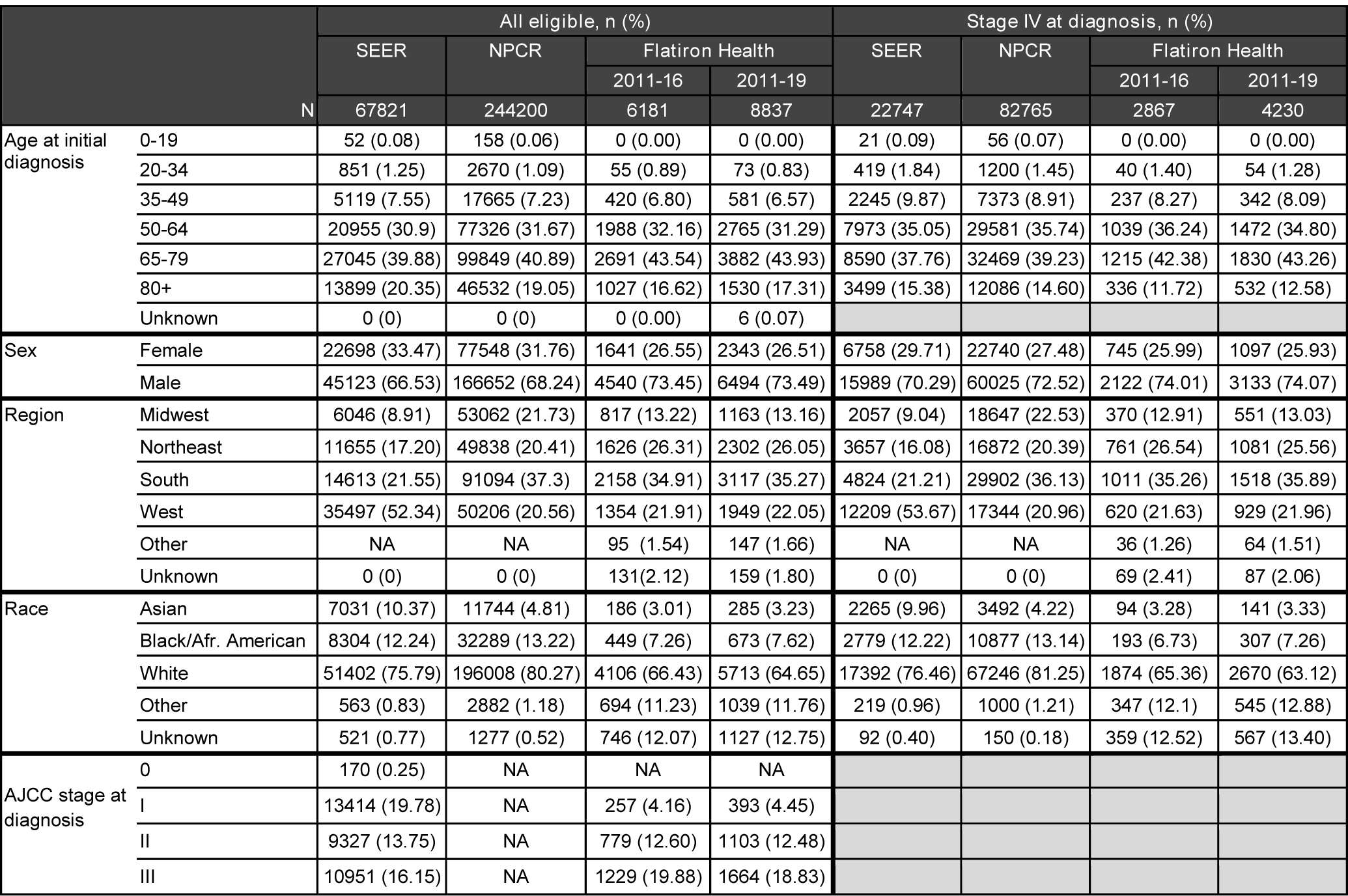
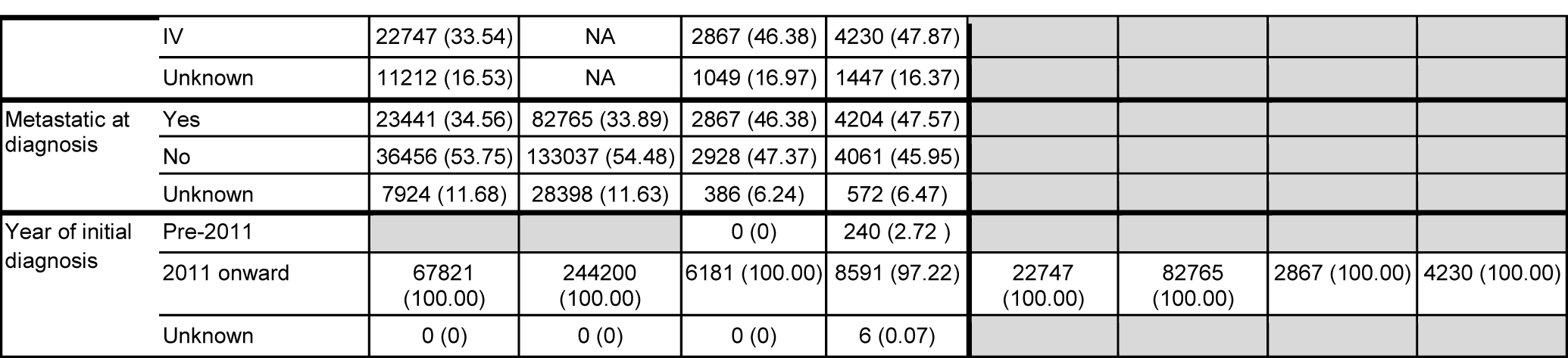
Characteristics for patients with esophageal/gastric carcinoma

**Table 10.** Characteristics for patients with hepatocellular carcinoma

| N |  | All eligible, n (%) |  |  |  | Stage IV at diagnosis, n (%) |  |  |  |
| --- | --- | --- | --- | --- | --- | --- | --- | --- | --- |
|  |  | SEER | NPCR | Flatiron Health |  | SEER | NPCR | Flatiron Health |  |
|  |  |  |  | 2011-16 | 2011-19 |  |  | 2011-16 | 2011-19 |
|  |  | 41443 | 131285 | 2744 | 4003 | 6908 | 19028 | 347 | 566 |
| Age at initial diagnosis | 0-19 | 69 (0.17) | 258 (0.20) | 0 (0.00) | 0 (0.00) | 24 (0.35) | 68 (0.36) | 0 (0.00) | 0 (0.00) |
|  | 20-34 | 244 (0.59) | 819 (0.62) | 15 (0.55) | 18 (0.45) | 76 (1.10) | 173 (0.91) | 2 (0.58) | 2 (0.35) |
|  | 35-49 | 1875 (4.52) | 5883 (4.48) | 93 (3.39) | 121 (3.02) | 339 (4.91) | 929 (4.88) | 8 (2.31) | 15 (2.65) |
|  | 50-64 | 20494 (49.45) | 66437 (50.61) | 1212 (44.17) | 1671 (41.74) | 3495 (50.59) | 9588 (50.39) | 156 (44.96) | 233 (41.17) |
|  | 65-79 | 14625 (35.29) | 45378 (34.56) | 1117 (40.71) | 1752 (43.77) | 2358 (34.13) | 6477 (34.04) | 148 (42.65) | 253 (44.70) |
|  | 80+ | 4136 (9.98) | 12510 (9.53) | 307 (11.19) | 441 (11.02) | 616 (8.92) | 1793 (9.42) | 33 (9.51) | 63 (11.13) |
| Sex | Female | 9617 (23.21) | 29896 (22.77) | 660 (24.05) | 941 (23.51) | 1303 (18.86) | 3674 (19.31) | 66 (9.02) | 102 (18.02) |
|  | Male | 31826 (76.79) | 101389 (77.23) | 2083 (75.91) | 3061 (76.47) | 5605 (81.14) | 15354 (80.69) | 281 (80.98) | 464 (81.98) |
|  | Unknown | 0 (0) | 0 (0) | 1 (0.04) | 1 (0.02) |  |  |  |  |
| Region | Midwest | 2965 (7.15) | 22598 (17.21) | 197 (7.18) | 317 (7.92) | 522 (7.56) | 3361 (17.66) | 7.49 (26) | 51 (9.01) |
|  | Northeast | 5224 (12.61) | 23610 (17.98) | 860 (31.34) | 1238 (30.93) | 912 (13.20) | 3419 (17.97) | 24.78 (86) | 135 (23.85) |
|  | South | 8332 (20.10) | 52854 (40.26) | 967 (35.24) | 1443 (36.05) | 1629 (23.58) | 8057 (42.34) | 155 (44.67) | 254 (44.88) |
|  | West | 24922 (60.14) | 32223 (24.54) | 634 (23.10) | 888 (22.18) | 3845 (55.66) | 4191 (22.03) | 71 (20.46) | 111 (19.61) |
|  | Other | NA | NA | 39 (1.42) | 61 (1.52) | NA | NA | 4 (1.15) | 8 (1.41) |
|  | Unknown | 0 (0) | 0 (0) | 47 (1.71) | 56 (1.40) | 0 (0) | 0 (0) | 5 (1.44) | 7 (1.24) |
| Race | Asian | 6098 (14.71) | 9879 (7.52) | 135 (4.92) | 190 (4.75) | 934 (13.52) | 1348 (7.08) | 20 (5.76) | 25 (4.42) |
|  | Black/Afr. American | 5781 (13.95) | 21529 (16.40) | 297 (10.82) | 438 (10.94) | 1141 (16.52) | 3562 (18.72) | 37 (10.66) | 61 (10.78) |
|  | White | 28722 (69.30) | 96995 (73.88) | 1597 (58.20) | 2318 (57.91) | 4703 (68.08) | 13753 (72.28) | 208 (59.94) | 328 (57.95) |
|  | Other | 582 (1.40) | 2486 (1.89) | 379 (13.81) | 563 (14.06) | 105 (1.52) | 335 (1.76) | 41 (11.82) | 77 (13.60) |
|  | Unknown | 260 (0.63) | 396 (0.30) | 336 (12.24) | 494 (12.34) | 25 (0.36) | 30 (0.16) | 41 (11.82) | 75 (13.25) |
| AJCC stage at diagnosis | I | 14388 (34.72) | NA | 179 (6.52) | 235 (5.87) |  |  |  |  |
|  | II | 7252 (17.50) | NA | 122 (4.45) | 183 (4.57) |  |  |  |  |
|  | III | 6754 (16.30) | NA | 189 (6.89) | 274 (6.84) |  |  |  |  |
|  | IV | 6908 (16.67) | NA | 347 (12.65) | 566 (14.14) |  |  |  |  |
|  | Unknown | 6141 (14.82) | NA | 1907 (69.50) | 2745 (68.57) |  |  |  |  |
| Metastatic at diagnosis | Yes | 5716 (13.79) | 19028 (14.49) | 307 (11.19) | 496 (12.39) |  |  |  |  |
|  | No | 31841 (76.83) | 99927 (76.11) | 908 (33.09) | 1310 (32.73) |  |  |  |  |
|  | Unknown | 3886 (9.38) | 12330 (9.39) | 1529 (55.72) | 2197 (54.88) |  |  |  |  |
| Year of initial diagnosis | Pre-2011 |  |  |  |  |  |  |  |  |
|  | 2011 onward | 41443 (100.00) | 131285 (100.00) | 2744 (100.00) | 4003 (100.00) | 6908 (100.00) | 19028 (100.00) | 347 (100.00) | 566 (100.00) |

**Table 11.**
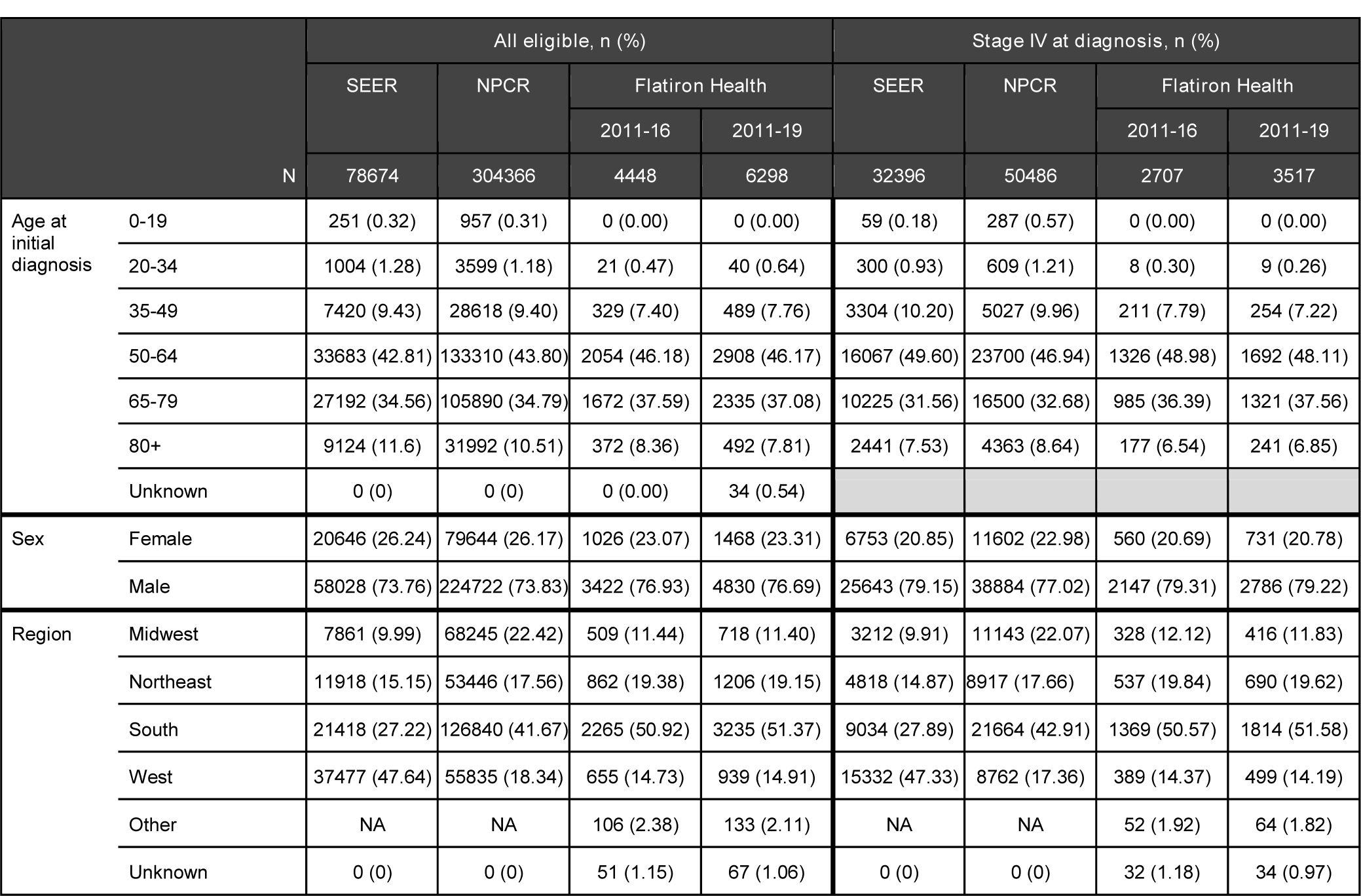
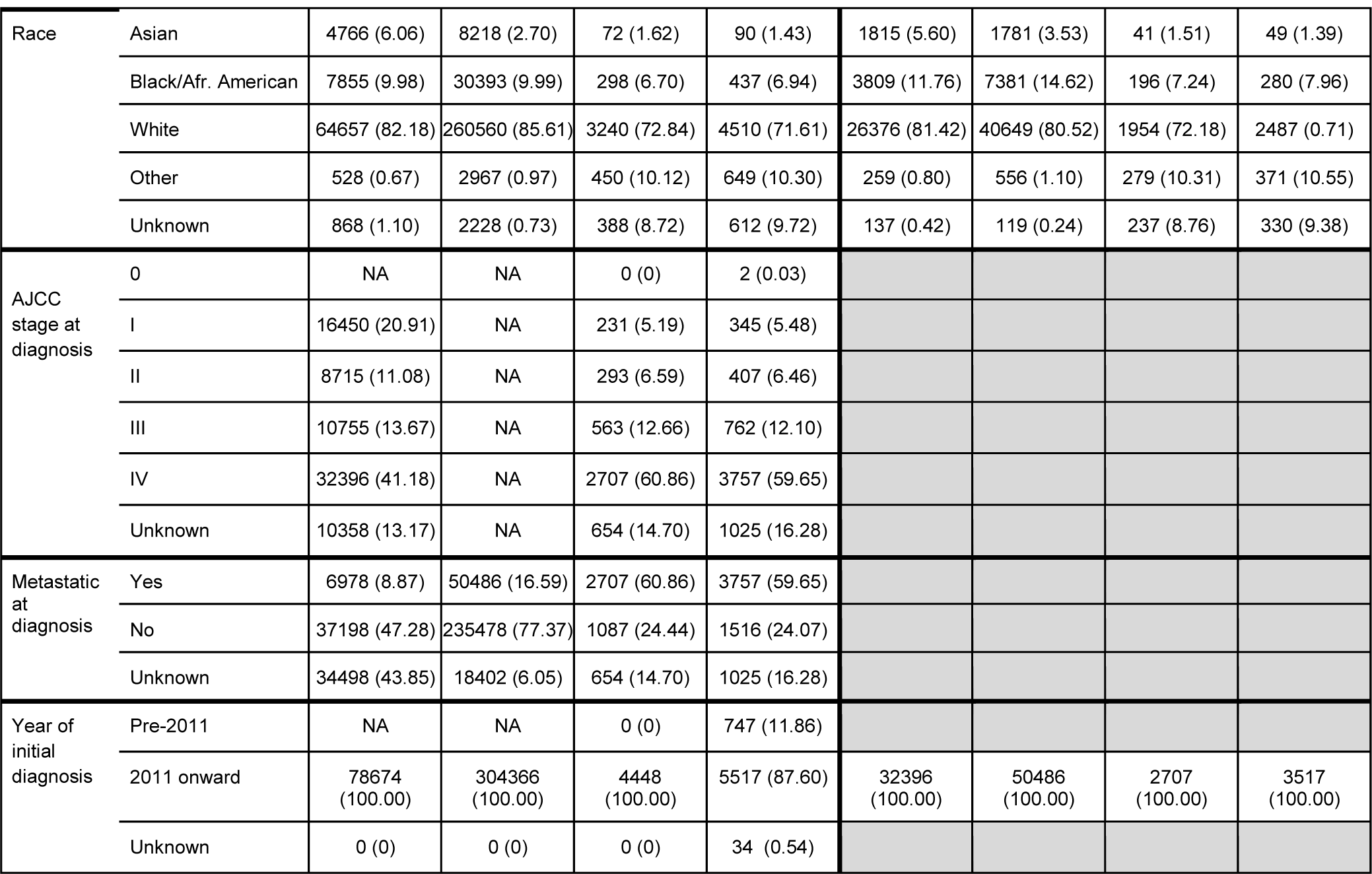
Characteristics for patients with head and neck cancer

**Table 12.**
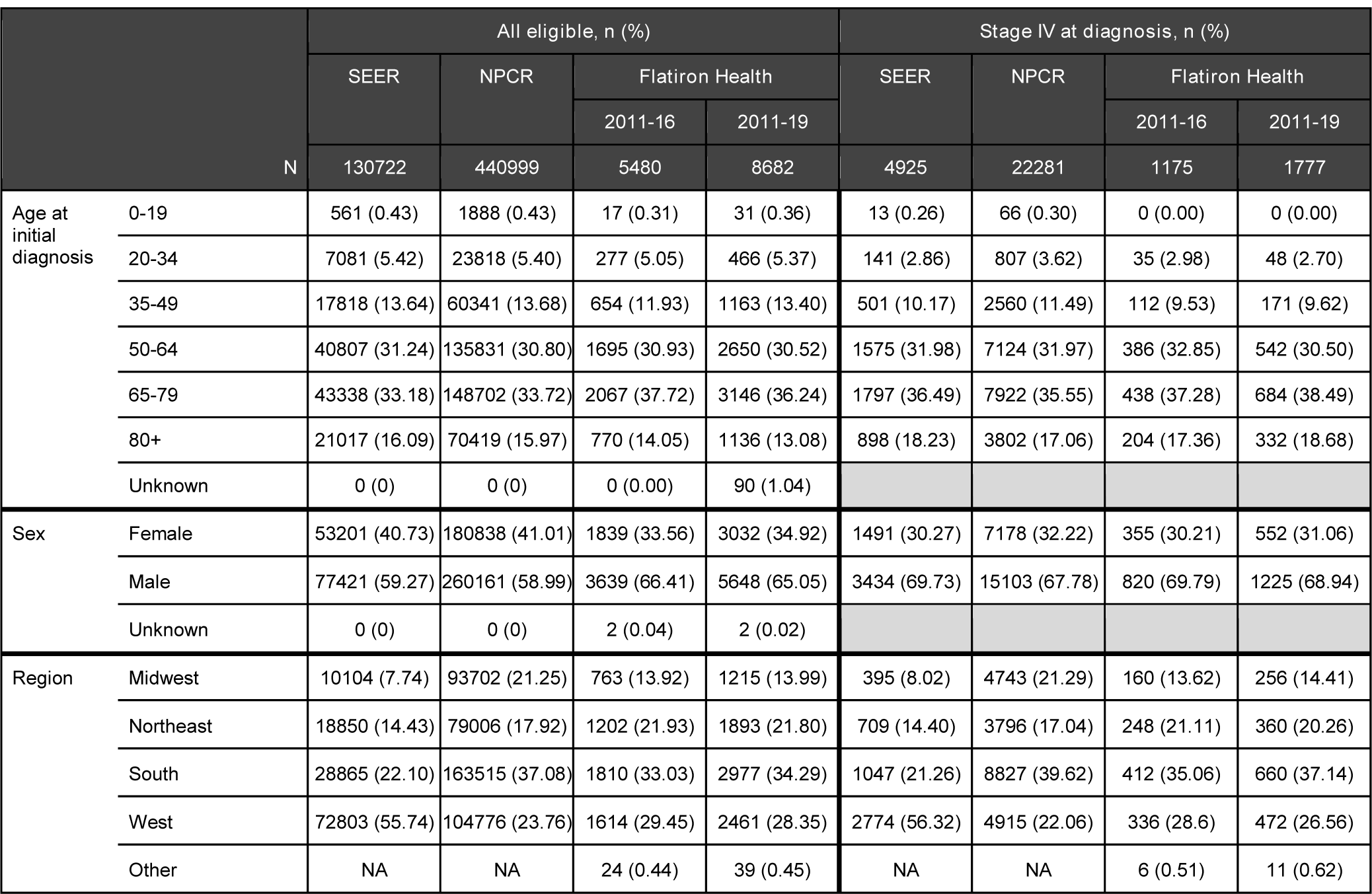
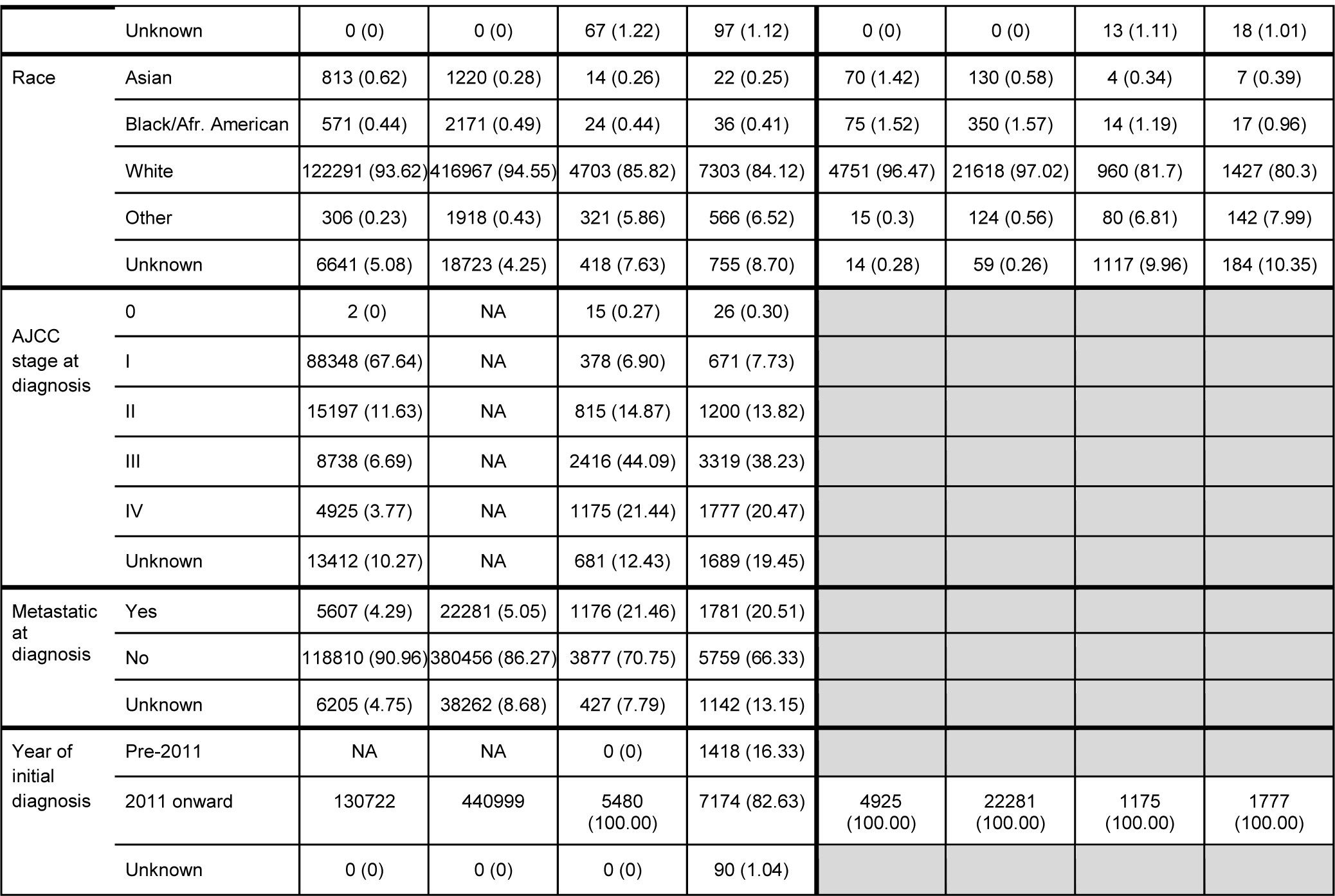
Characteristics for patients with melanoma

**Table 13.**
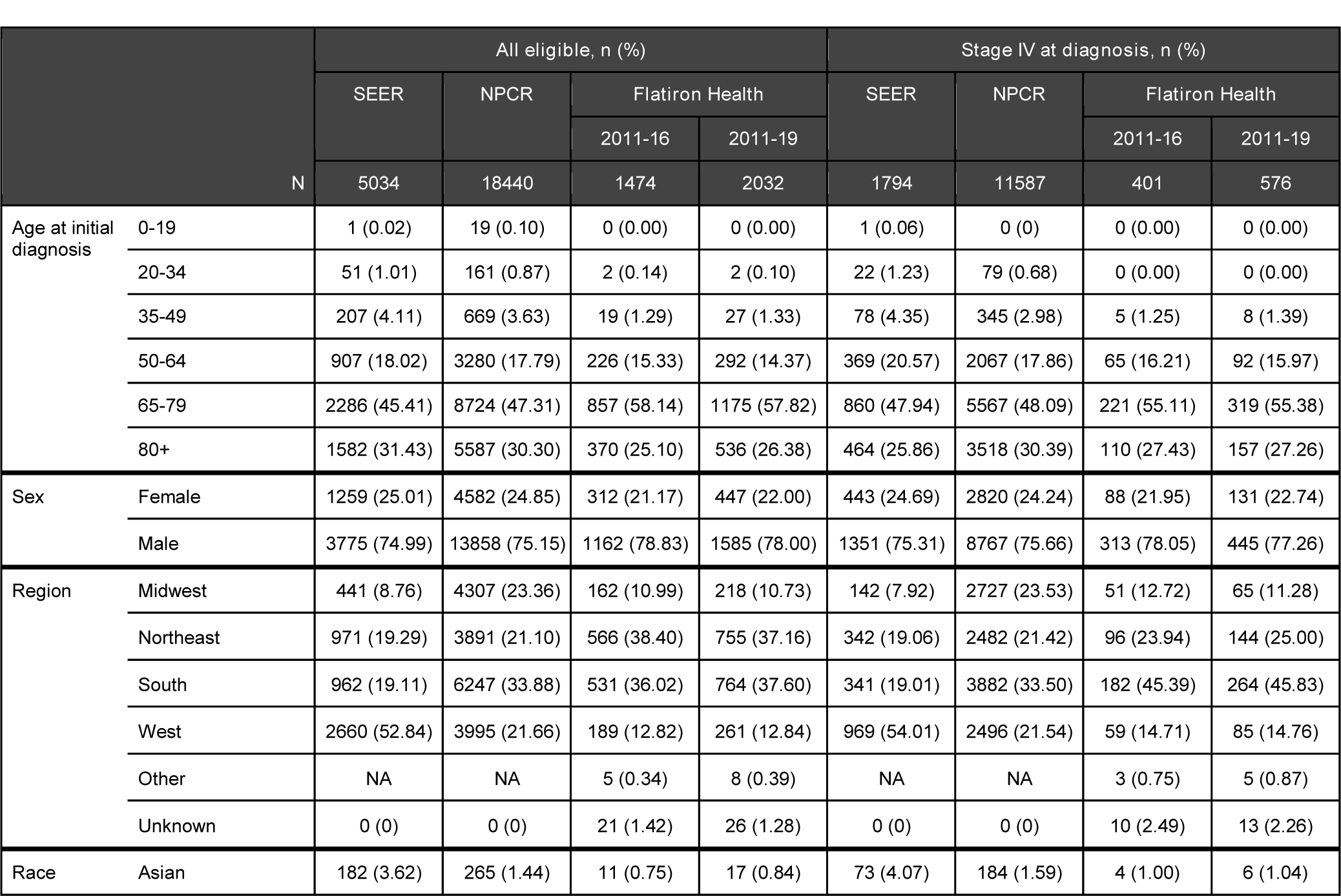
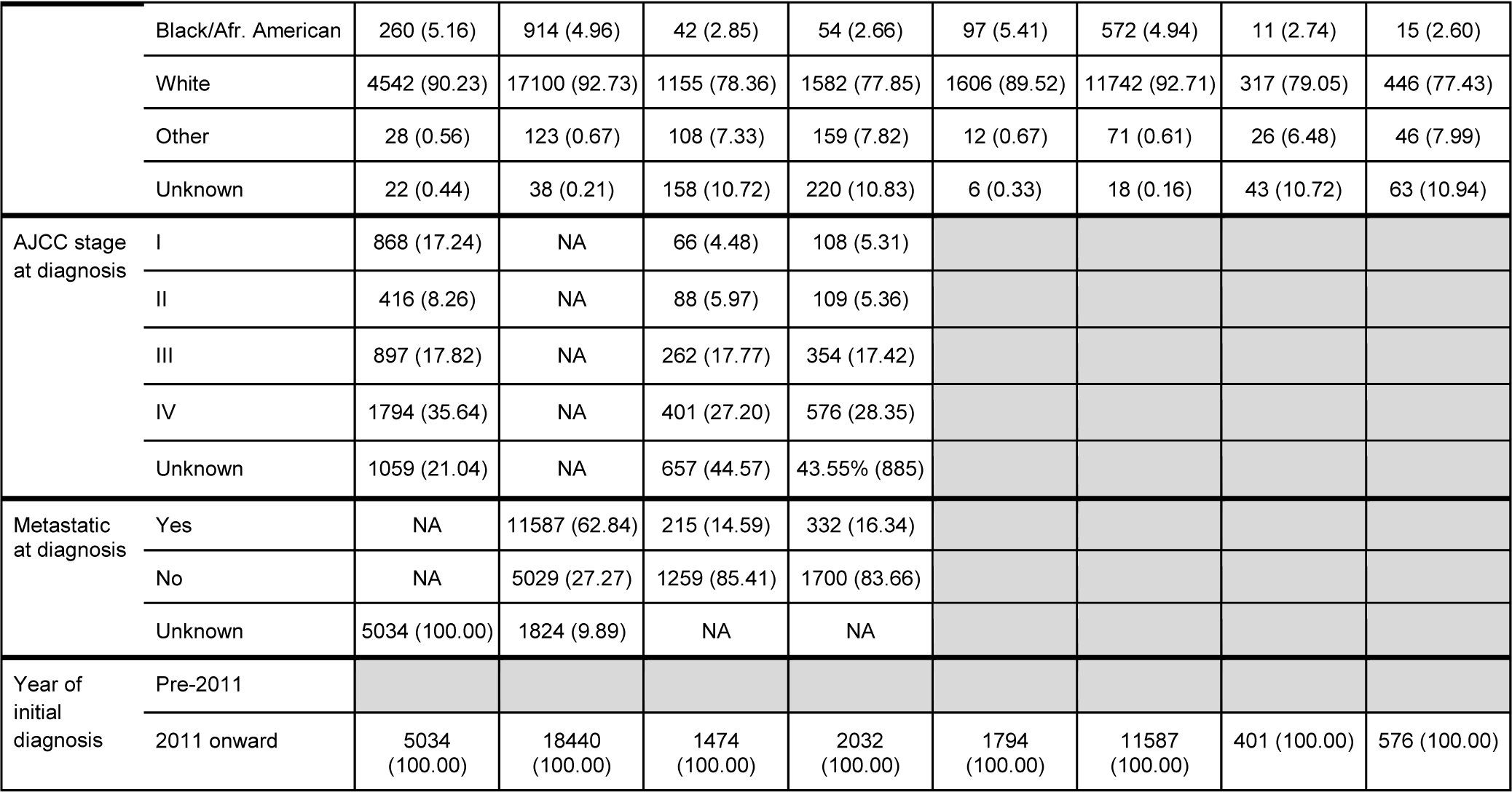
Characteristics for patients with malignant pleural mesothelioma

**Table 14.** Characteristics for patients with multiple myeloma

|  |  | All eligible, n (%) |  |  |  | Stage IV at diagnosis, n (%) |  |  |  |
| --- | --- | --- | --- | --- | --- | --- | --- | --- | --- |
|  |  | SEER | NPCR | Flatiron Health |  | SEER | NPCR | Flatiron Health |  |
|  |  |  |  | 2011-16 | 2011-19 |  |  | 2011-16 | 2011-19 |
| N |  | 38519 | 139422 | 6927 | 9696 |  |  |  |  |
| Age at initial diagnosis | 0-19 | 10 (0.03) | 30 (0.02) | 0 (0.00) | 0 (0.00) |  |  |  |  |
|  | 20-34 | 193 (0.50) | 642 (0.46) | 27 (0.39) | 34 (0.35) |  |  |  |  |
|  | 35-49 | 2511 (6.52) | 8954 (6.42) | 381 (5.50) | 522 (5.38) |  |  |  |  |
|  | 50-64 | 11557 (30.00) | 41842 (30.01) | 2130 (30.75) | 2871 (29.61) |  |  |  |  |
|  | 65-79 | 16660 (43.25) | 61575 (44.16) | 3180 (45.91) | 4552 (46.95) |  |  |  |  |
|  | 80+ | 7588 (19.70) | 26379 (18.92) | 1209 (17.45) | 1717 (17.71) |  |  |  |  |
| Sex | Female | 16870 (43.80) | 61494 (44.11) | 3254 (46.98) | 4466 (46.06) |  |  |  |  |
|  | Male | 21649 (56.20) | 77928 (55.89) | 3673 (53.02) | 5230 (53.94) |  |  |  |  |
| Region | West | 3852 (10.00) | 28852 (20.69) | 879 (12.69) | 1222 (12.60) |  |  |  |  |
|  | Northeast | 6420 (16.67) | 26944 (19.33) | 1936 (27.95) | 2681 (27.65) |  |  |  |  |
|  | South | 9677 (25.12) | 57654 (41.35) | 2578 (37.22) | 3678 (37.93) |  |  |  |  |
|  | Midwest | 18570 (48.21) | 25972 (18.63) | 1315 (18.98) | 1840 (18.98) |  |  |  |  |
|  | Other | NA | NA | 108 (1.56) | 1.59 (154) |  |  |  |  |
|  | Unknown | 0 (0) | 0 (0) | 111 (1.60) | 121 (1.251) |  |  |  |  |
| Race | Asian | 2182 (5.66) | 3358 (2.41) | 123 (1.78) | 170 (1.75) |  |  |  |  |
|  | Black/Afr. American | 7759 (20.14) | 29076 (20.85) | 1115 (16.10) | 1564 (16.13) |  |  |  |  |
|  | White | 27912 (72.46) | 104409 (74.89) | 4332 (62.54) | 5891 (60.76) |  |  |  |  |
|  | Other | 245 (0.64) | 1434 (1.03) | 743 (10.73) | 1076 (11.10) |  |  |  |  |
|  | Unknown | 421 (1.09) | 1145 (0.82) | 614 (8.86) | 995 (10.26) |  |  |  |  |
| ISS stage at diagnosis | I | NA | NA | 1115 (16.10) | 1654 (17.06) |  |  |  |  |
|  | II | NA | NA | 1065 (15.37) | 1598 (16.48) |  |  |  |  |
|  | III | NA | NA | 1044 (15.07) | 1582 (16.32) |  |  |  |  |
|  | Unknown | 38519 (100.00)) | NA | 3703 (53.46) | 4862 (50.14) |  |  |  |  |
| Metastatic at diagnosis | Yes | 36762 (95.44) | 132842 (95.28) | 6927 (100.00) | 9696 (100.00) |  |  |  |  |
|  | No | 1700 (4.41) | 6399 (4.59) | NA | NA |  |  |  |  |
|  | Unknown | 57 (0.15) | 181 (0.13) | NA | NA |  |  |  |  |
| Year of initial diagnosis | Pre-2011 |  |  |  |  |  |  |  |  |
|  | 2011 onward | 38519 (100.00) | 139422 (100.00) | 6927 (100.00) | 9696 (100.00) |  |  |  |  |

**Table 15.**
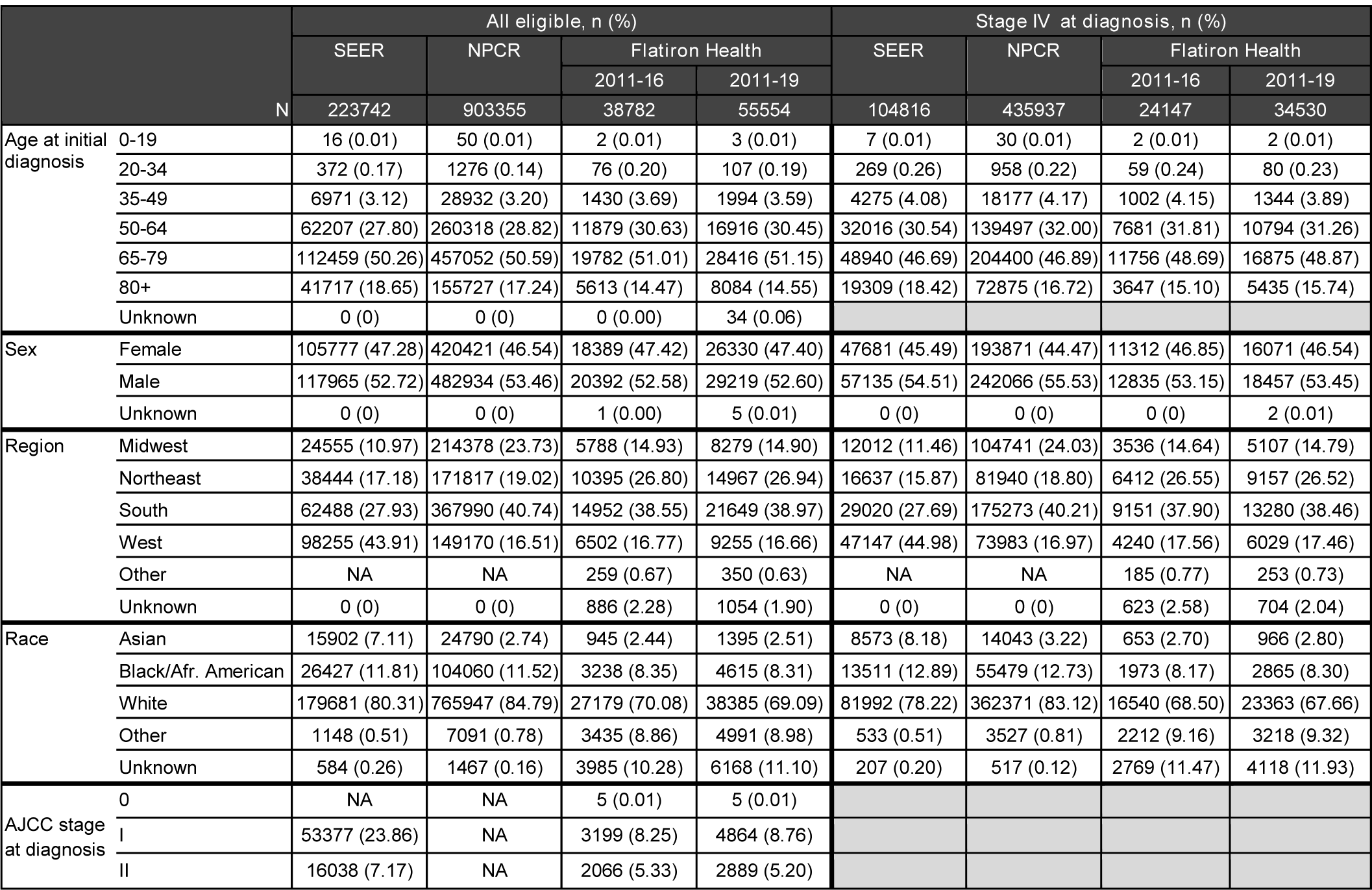
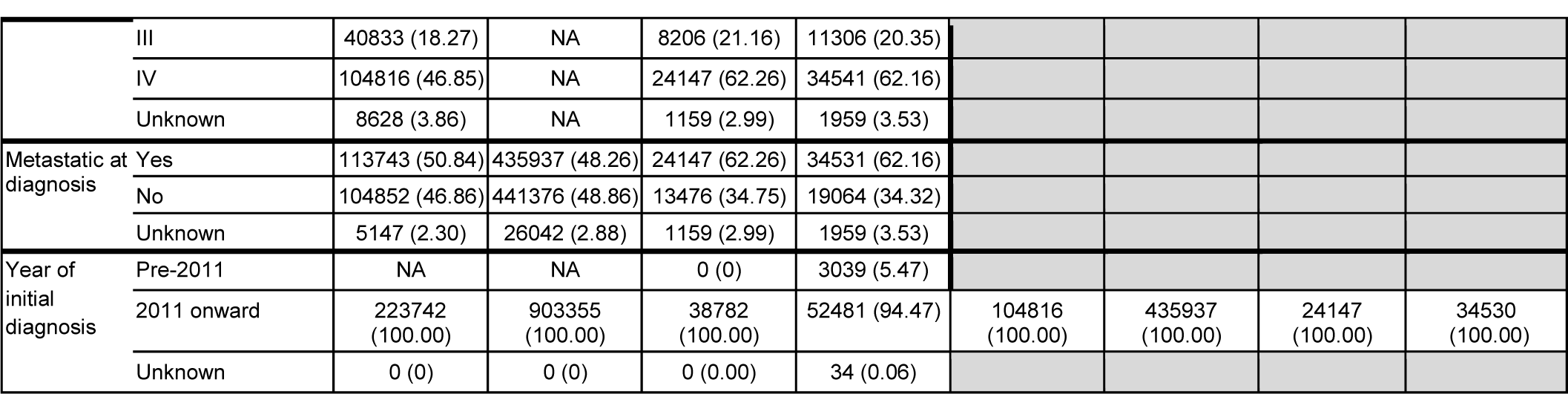
Characteristics for patients with non-small cell lung cancer (NSCLC)

**Table 16.**
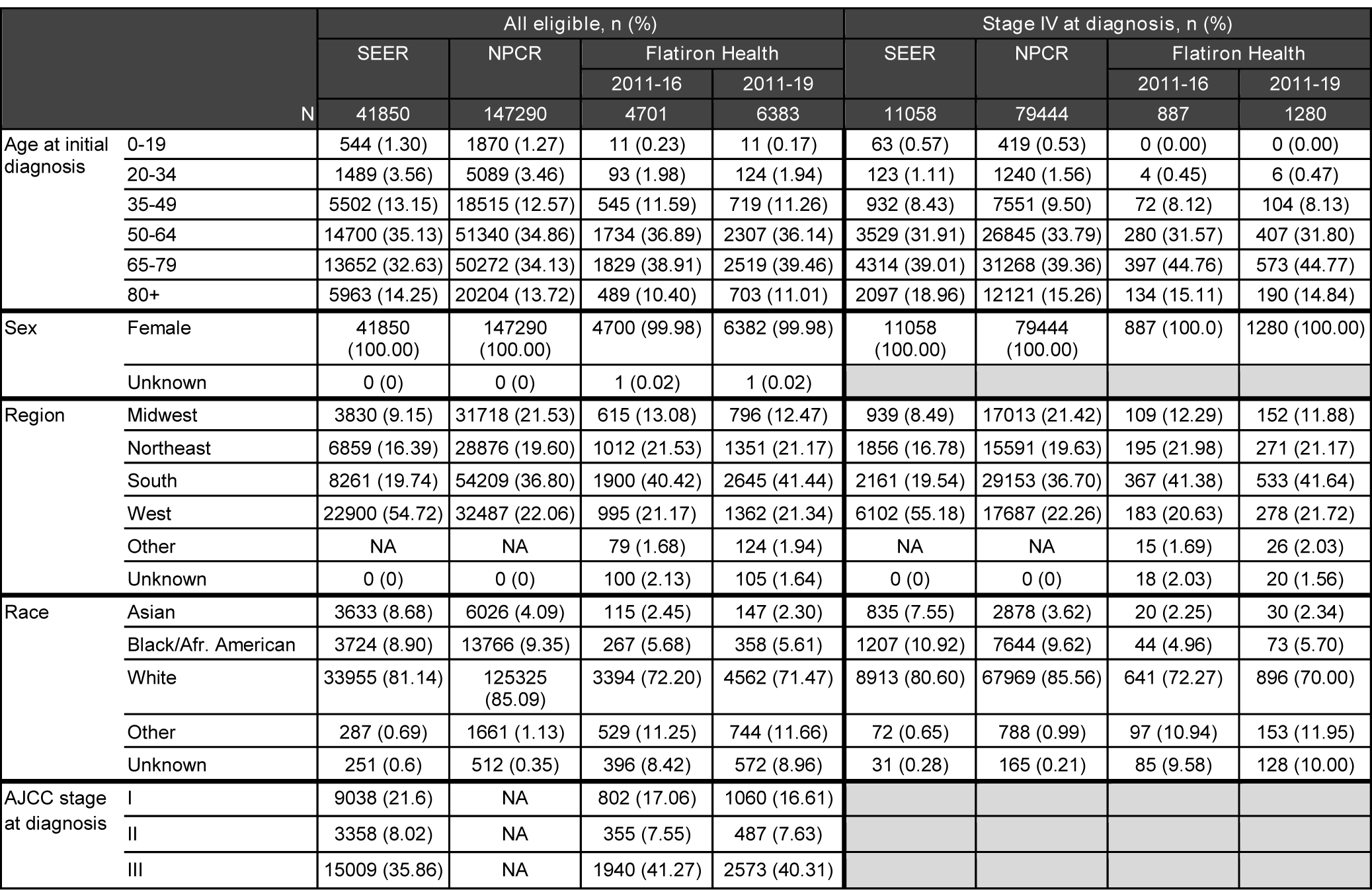
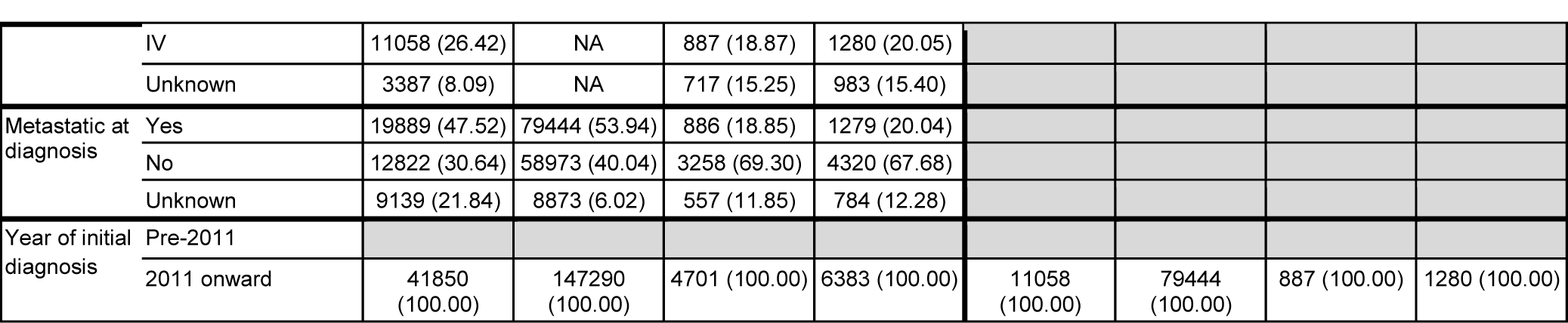
Characteristics for patients with ovarian carcinoma

**Table 17.**
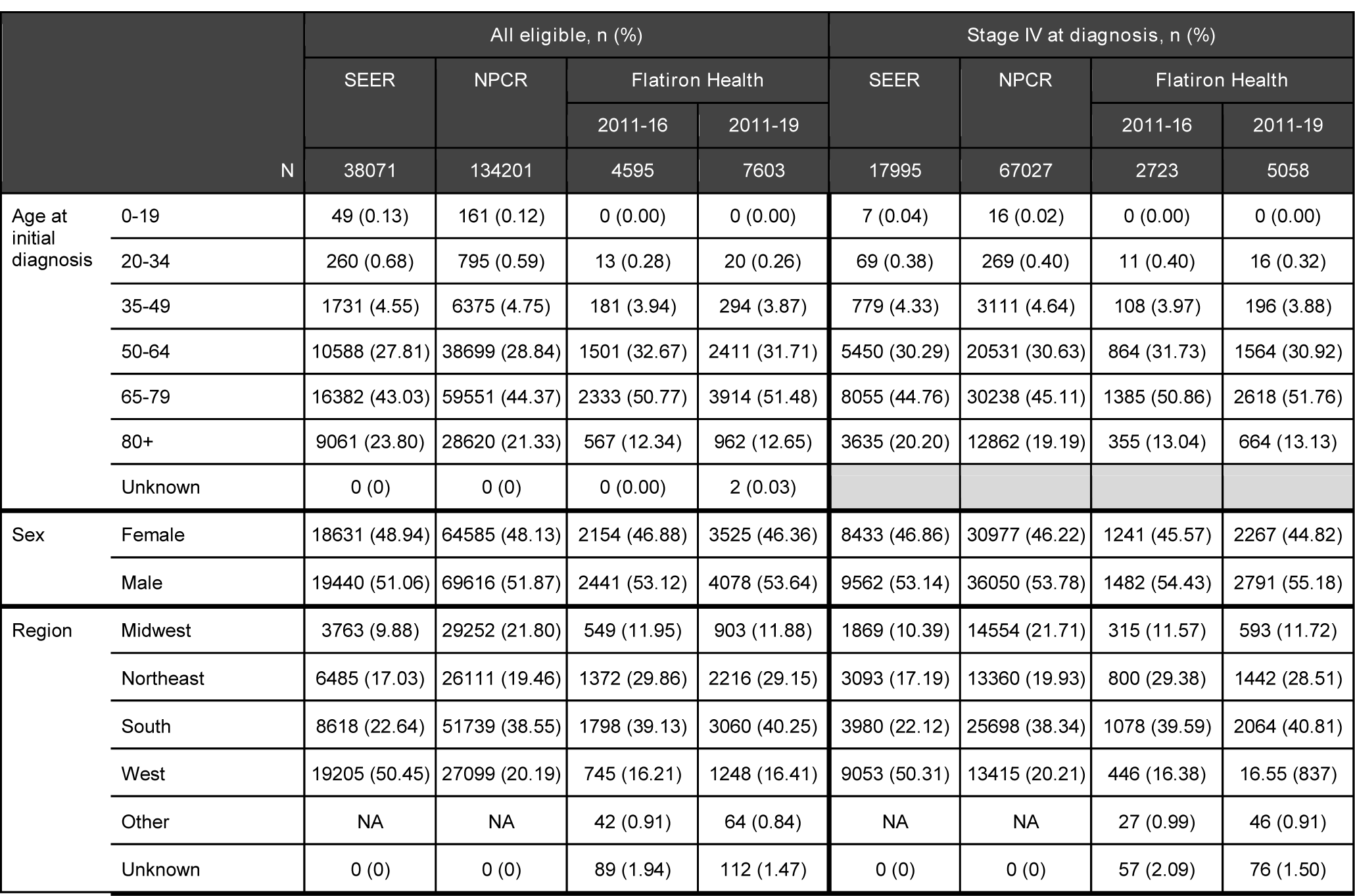
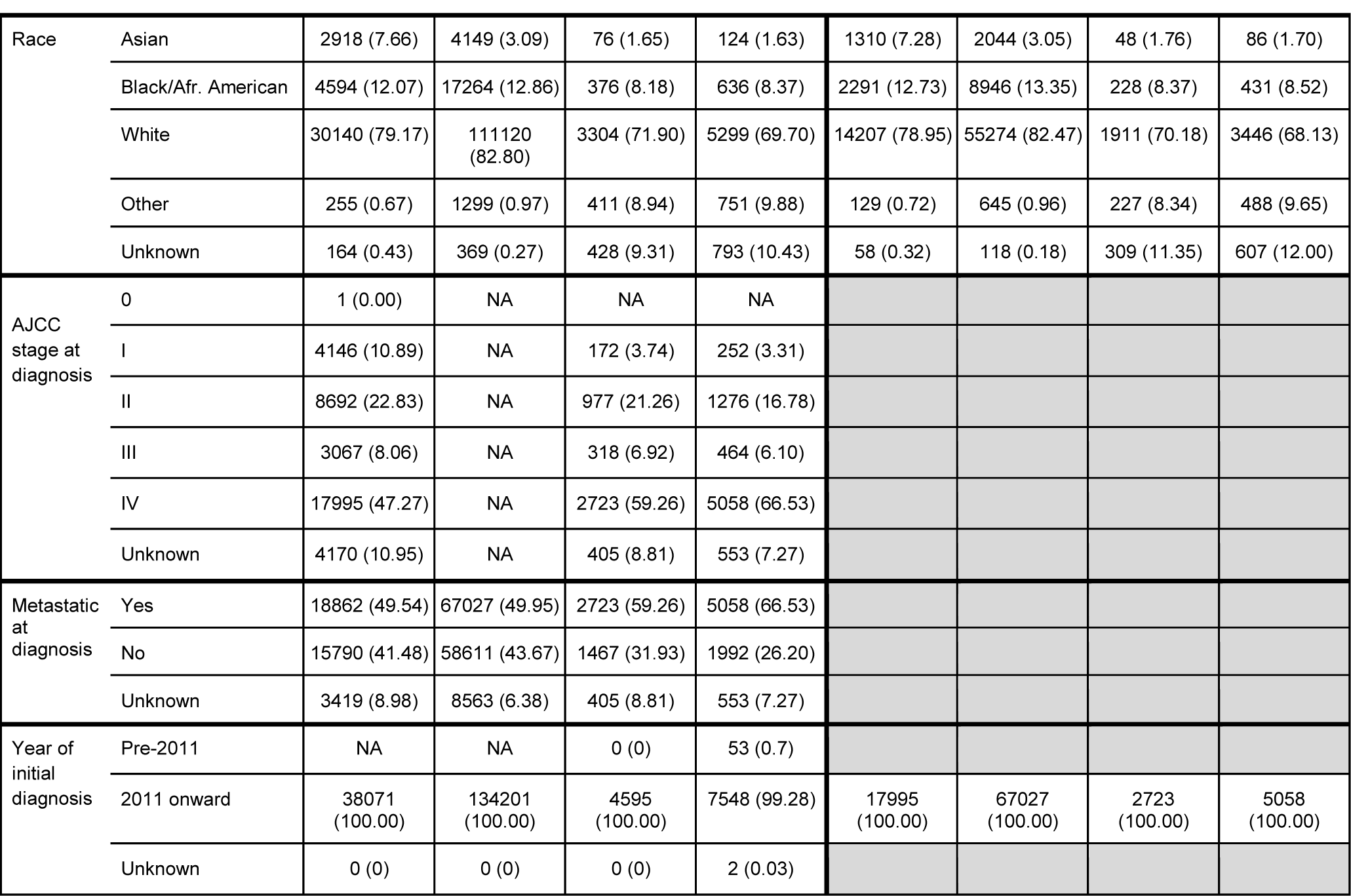
Characteristics for patients with pancreatic carcinoma

| N |  | All eligible, n (%) |  |  |  | Stage IV at diagnosis, n (%) |  |  |  |
| --- | --- | --- | --- | --- | --- | --- | --- | --- | --- |
|  |  | SEER | NPCR | Flatiron Health |  | SEER | NPCR | Flatiron Health |  |
|  |  |  |  | 2011-16 | 2011-19 |  |  | 2011-16 | 2011-19 |
|  |  | 38071 | 134201 | 4595 | 7603 | 17995 | 67027 | 2723 | 5058 |
| Age at initial diagnosis | 0-19 | 49 (0.13) | 161 (0.12) | 0 (0.00) | 0 (0.00) | 7 (0.04) | 16 (0.02) | 0 (0.00) | 0 (0.00) |
|  | 20-34 | 260 (0.68) | 795 (0.59) | 13 (0.28) | 20 (0.26) | 69 (0.38) | 269 (0.40) | 11 (0.40) | 16 (0.32) |
|  | 35-49 | 1731 (4.55) | 6375 (4.75) | 181 (3.94) | 294 (3.87) | 779 (4.33) | 3111 (4.64) | 108 (3.97) | 196 (3.88) |
|  | 50-64 | 10588 (27.81) | 38699 (28.84) | 1501 (32.67) | 2411 (31.71) | 5450 (30.29) | 20531 (30.63) | 864 (31.73) | 1564 (30.92) |
|  | 65-79 | 16382 (43.03) | 59551 (44.37) | 2333 (50.77) | 3914 (51.48) | 8055 (44.76) | 30238 (45.11) | 1385 (50.86) | 2618 (51.76) |
|  | 80+ | 9061 (23.80) | 28620 (21.33) | 567 (12.34) | 962 (12.65) | 3635 (20.20) | 12862 (19.19) | 355 (13.04) | 664 (13.13) |
|  | Unknown | 0 (0) | 0 (0) | 0 (0.00) | 2 (0.03) |  |  |  |  |
| Sex | Female | 18631 (48.94) | 64585 (48.13) | 2154 (46.88) | 3525 (46.36) | 8433 (46.86) | 30977 (46.22) | 1241 (45.57) | 2267 (44.82) |
|  | Male | 19440 (51.06) | 69616 (51.87) | 2441 (53.12) | 4078 (53.64) | 9562 (53.14) | 36050 (53.78) | 1482 (54.43) | 2791 (55.18) |
| Region | Midwest | 3763 (9.88) | 29252 (21.80) | 549 (11.95) | 903 (11.88) | 1869 (10.39) | 14554 (21.71) | 315 (11.57) | 593 (11.72) |
|  | Northeast | 6485 (17.03) | 26111 (19.46) | 1372 (29.86) | 2216 (29.15) | 3093 (17.19) | 13360 (19.93) | 800 (29.38) | 1442 (28.51) |
|  | South | 8618 (22.64) | 51739 (38.55) | 1798 (39.13) | 3060 (40.25) | 3980 (22.12) | 25698 (38.34) | 1078 (39.59) | 2064 (40.81) |
|  | West | 19205 (50.45) | 27099 (20.19) | 745 (16.21) | 1248 (16.41) | 9053 (50.31) | 13415 (20.21) | 446 (16.38) | 16.55 (837) |
|  | Other | NA | NA | 42 (0.91) | 64 (0.84) | NA | NA | 27 (0.99) | 46 (0.91) |
|  | Unknown | 0 (0) | 0 (0) | 89 (1.94) | 112 (1.47) | 0 (0) | 0 (0) | 57 (2.09) | 76 (1.50) |
| Race | Asian | 2918 (7.66) | 4149 (3.09) | 76 (1.65) | 124 (1.63) | 1310 (7.28) | 2044 (3.05) | 48 (1.76) | 86 (1.70) |
|  | Black/Afr. American | 4594 (12.07) | 17264 (12.86) | 376 (8.18) | 636 (8.37) | 2291 (12.73) | 8946 (13.35) | 228 (8.37) | 431 (8.52) |
|  | White | 30140 (79.17) | 111120 (82.80) | 3304 (71.90) | 5299 (69.70) | 14207 (78.95) | 55274 (82.47) | 1911 (70.18) | 3446 (68.13) |
|  | Other | 255 (0.67) | 1299 (0.97) | 411 (8.94) | 751 (9.88) | 129 (0.72) | 645 (0.96) | 227 (8.34) | 488 (9.65) |
|  | Unknown | 164 (0.43) | 369 (0.27) | 428 (9.31) | 793 (10.43) | 58 (0.32) | 118 (0.18) | 309 (11.35) | 607 (12.00) |
| AJCC stage at diagnosis | 0 | 1 (0.00) | NA | NA | NA |  |  |  |  |
|  | I | 4146 (10.89) | NA | 172 (3.74) | 252 (3.31) |  |  |  |  |
|  | II | 8692 (22.83) | NA | 977 (21.26) | 1276 (16.78) |  |  |  |  |
|  | III | 3067 (8.06) | NA | 318 (6.92) | 464 (6.10) |  |  |  |  |
|  | IV | 17995 (47.27) | NA | 2723 (59.26) | 5058 (66.53) |  |  |  |  |
|  | Unknown | 4170 (10.95) | NA | 405 (8.81) | 553 (7.27) |  |  |  |  |
| Metastatic at diagnosis | Yes | 18862 (49.54) | 67027 (49.95) | 2723 (59.26) | 5058 (66.53) |  |  |  |  |
|  | No | 15790 (41.48) | 58611 (43.67) | 1467 (31.93) | 1992 (26.20) |  |  |  |  |
|  | Unknown | 3419 (8.98) | 8563 (6.38) | 405 (8.81) | 553 (7.27) |  |  |  |  |
| Year of initial diagnosis | Pre-2011 | NA | NA | 0 (0) | 53 (0.7) |  |  |  |  |
|  | 2011 onward | 38071 (100.00) | 134201 (100.00) | 4595 (100.00) | 7548 (99.28) | 17995 (100.00) | 67027 (100.00) | 2723 (100.00) | 5058 (100.00) |
|  | Unknown | 0 (0) | 0 (0) | 0 (0) | 2 (0.03) |  |  |  |  |

**Table 18.**
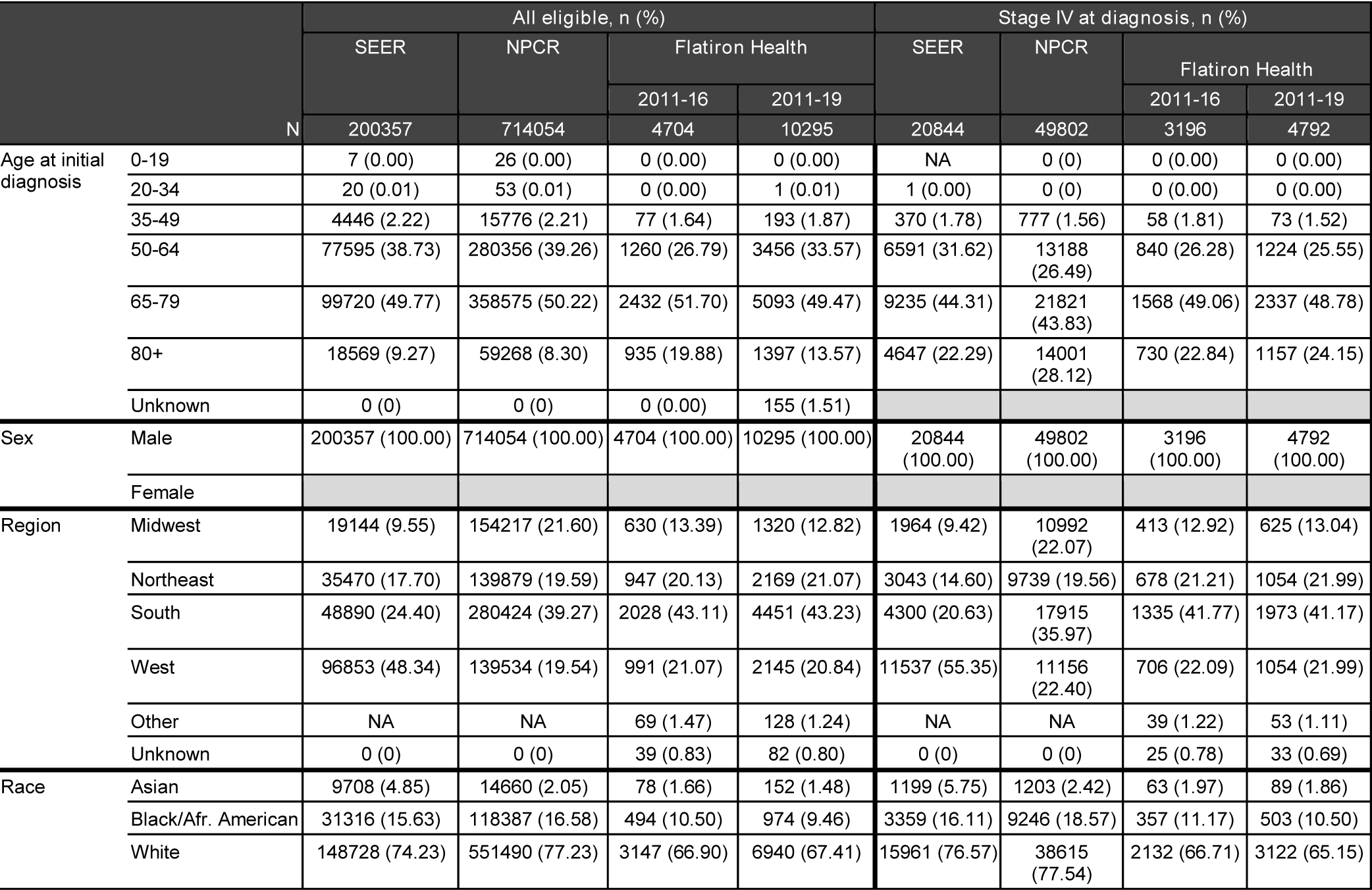
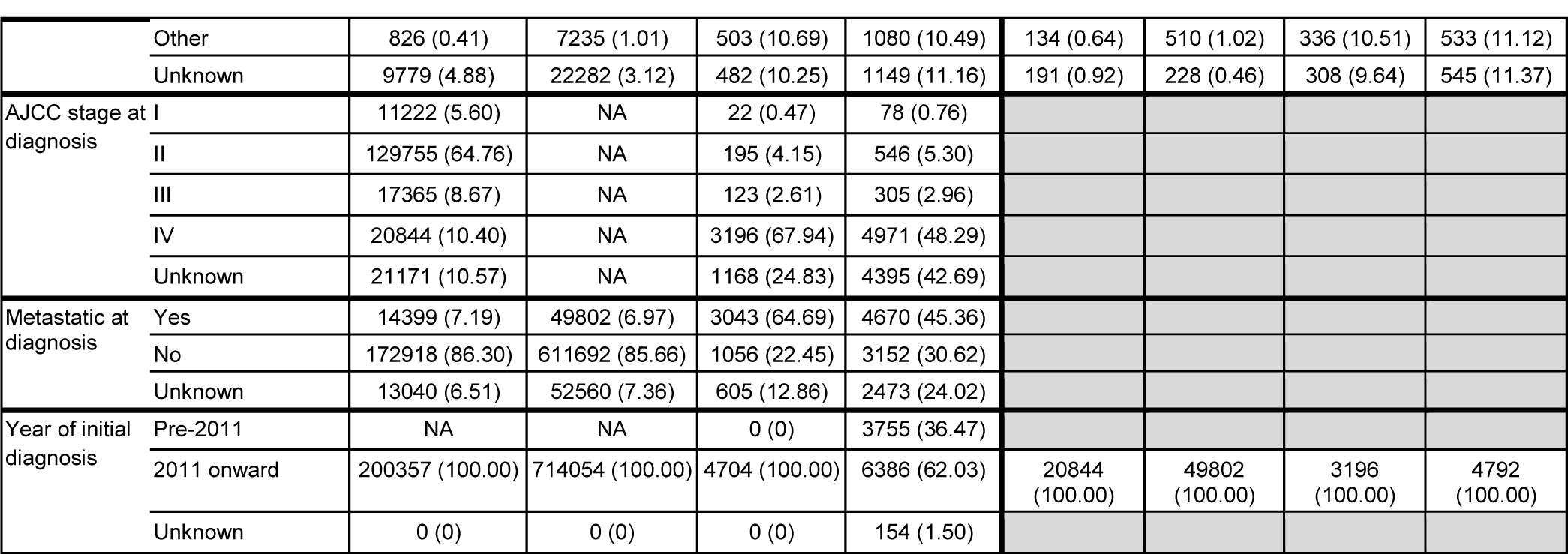
Characteristics for patients with prostate cancer

**Table 19.**
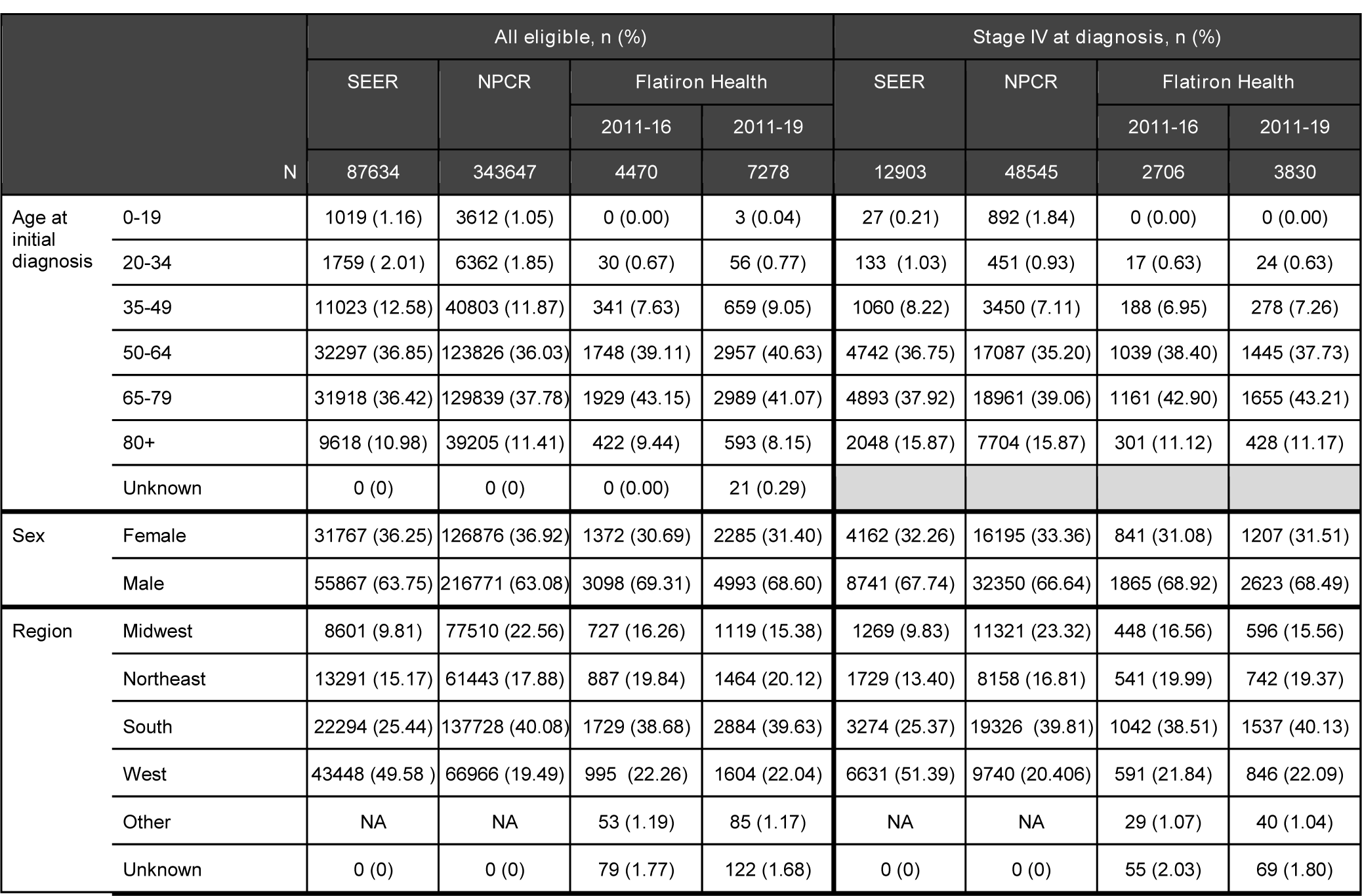
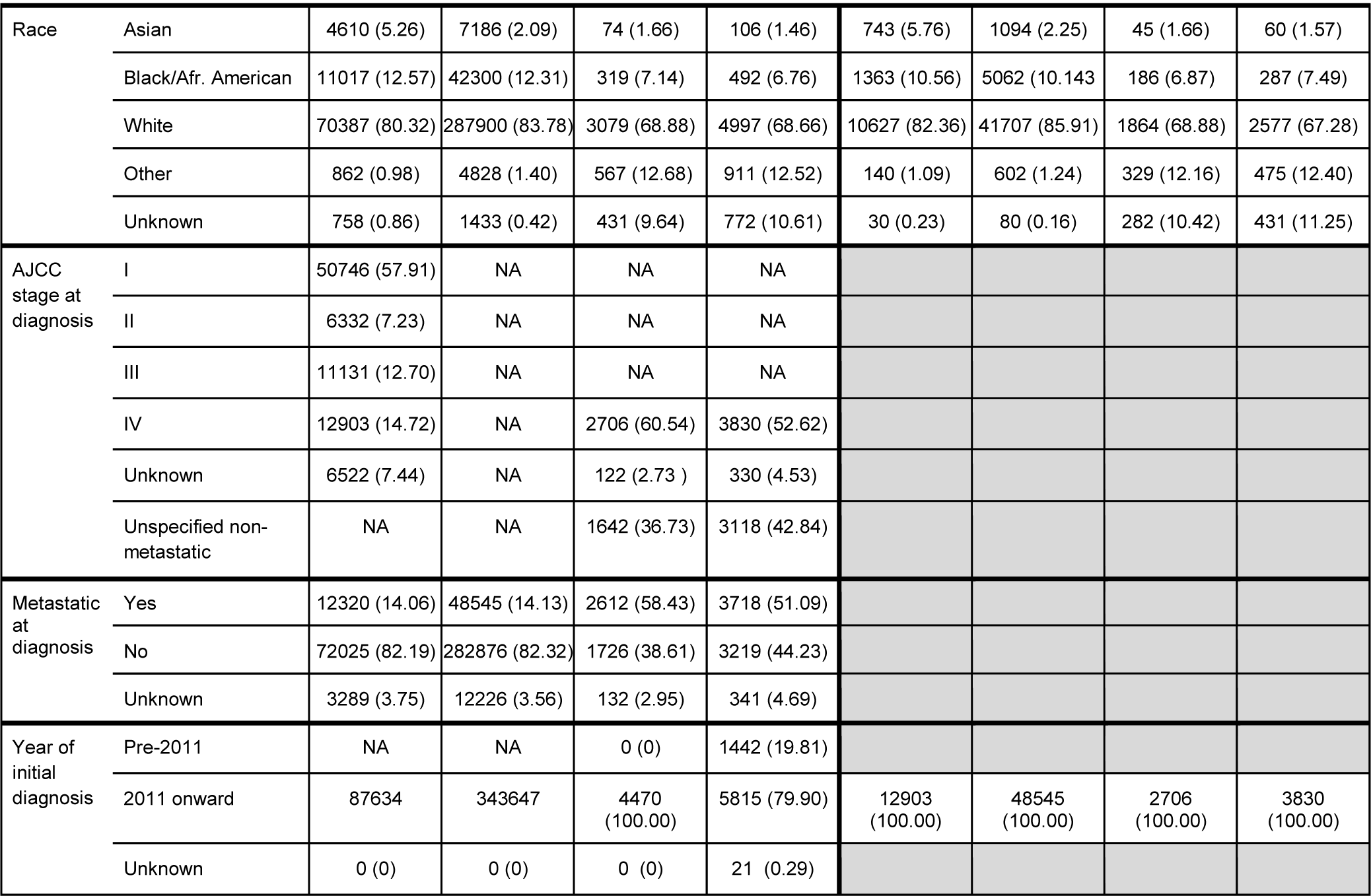
Characteristics for patients with renal-cell carcinoma

**Table 20.**
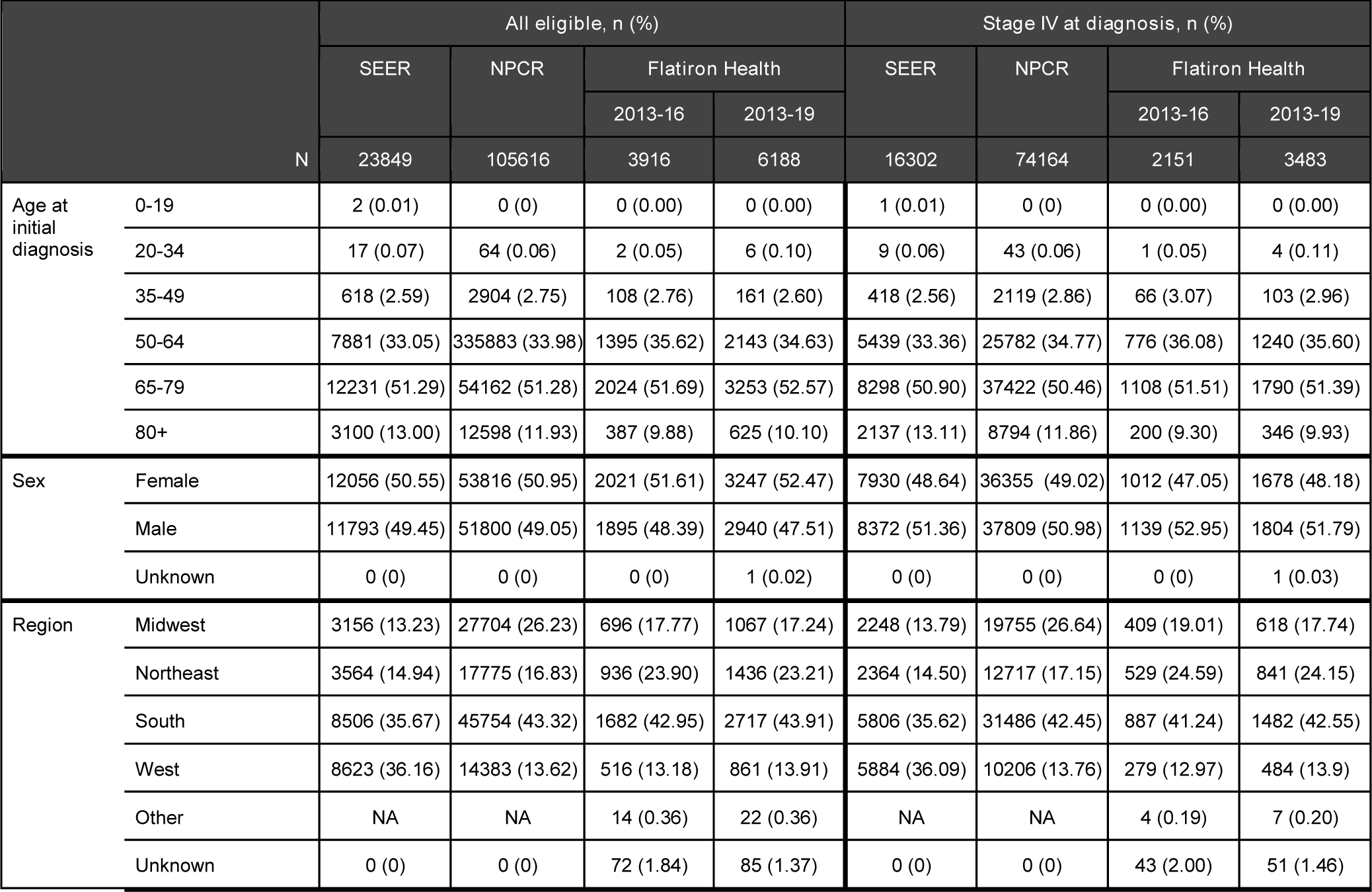
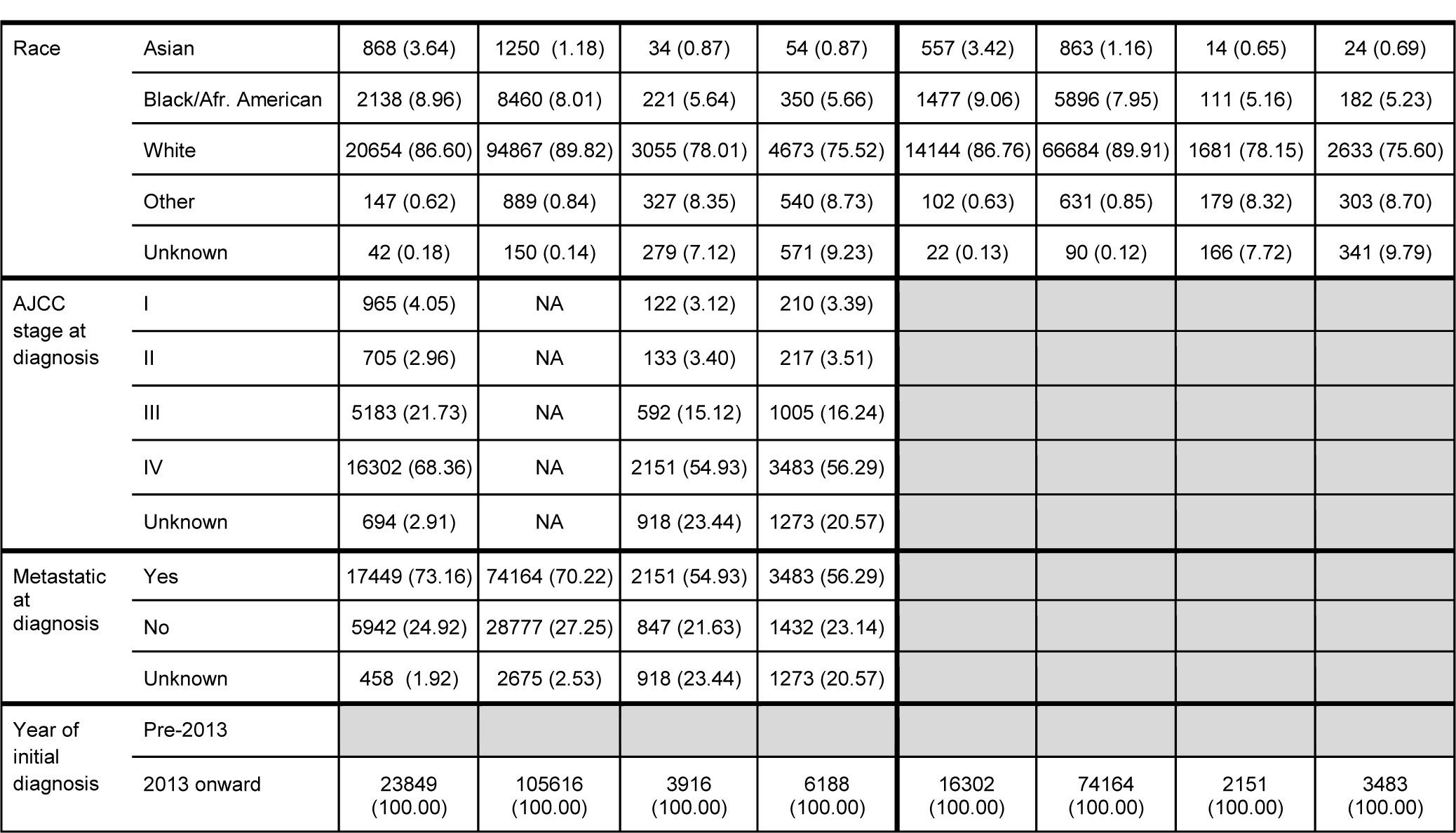
Characteristics for patients with small-cell lung cancer

## RESULTS

Among the 2.2 million patients with cancer in the Flatiron Health database as of May 2019, 201,570 were included in this analysis, as well as 1,719,277 and 6,308,342 cases from the SEER and NPCR, respectively. The disease-specific databases vary in size, depending on the incidence and prevalence of the disease, and in the case of Flatiron Health, on the length of time the database has been active. The largest comparisons corresponded to 55,554 vs 273,742 and 903,355 patients with NSCLC, and the smallest to 1,116 vs 18,148 and 72,575 for patients with FL.

Tables 2–20 present the comparisons for the following tumor types: advanced urothelial (or bladder), metastatic breast cancer, early breast cancer, CLL, metastatic colorectal cancer, DLBCL, FL, advanced gastric/esophageal carcinoma, HCC, advanced head and neck cancer, advanced melanoma, malignant pleural mesothelioma, MM, advanced NSCLC, ovarian carcinoma, metastatic pancreatic carcinoma, metastatic prostate cancer, metastatic RCC, and SCLC.

## DISCUSSION

Results reported in this descriptive study provide an overview of the originating sources, data collection methods, and comparative population characteristics for three oncology-specific RWD sources in the US: the Flatiron Health, SEER, and NPCR data. We focused our comparisons on baseline demographic and clinical variables at the time of initial cancer diagnosis that describe the populations included in these data sources, based on the data elements commonly found in all three of them. Each of these three data sources relies on different collection approaches (Table 1), and our findings reveal population differences likely stemming from those distinct collection strategies. These differences in data collection methods and resulting populations should be considered when determining whether a dataset is fit-for-use for a particular research question and can help to contextualize research results obtained when using each data source. To further assist in that contextualization, this discussion highlights some of the potential underlying explanations for the differences observed.

The distribution of patients according to sex/gender in the three data sources was comparable, but there were noticeable differences for other variables. Regarding regional distribution, Flatiron Health and the NPCR data were most closely aligned to the regional population distribution in the most recent US census (24). Due to its design, SEER data diverges the most from the census, particularly overrepresenting the West, while the Flatiron Health database provides a convenience sample that is slightly weighted towards the South and underrepresents the Western region.

The three data sources had overall similar age distributions, with a trend toward a modestly lower proportion of patients over 80 years at diagnosis in the Flatiron Health database (generally ≤5% lower across most diseases). These slight differences could be secondary to the algorithmic transformation of birth year for select elderly patients, which is required to reduce the risk of re-identification (see Appendix II). Another potential reason for modest discrepancies in the oldest age category is the difference in information sources that feed each one of them. State registries collect information regardless of patients’ site of care and from death certificates and autopsy reports (25), while Flatiron Health databases accrue information only via oncology clinics. By focusing on specialized care, Flatiron Health has limited reach into general hospice or other geriatric care settings, where some elderly patients may be referred before they complete two visits to an oncology clinic (therefore excluding them from eligibility into Flatiron Health databases). This mechanism may account for the larger age discrepancies seen in some diseases such as pancreatic cancer, as this is an aggressive cancer in which elderly patients may be more likely to be referred to hospice early in the disease course. Of note, the larger age discrepancy seen in CLL may be related to further differences in clinical inclusion criteria between the three databases, as patients are only included in the Flatiron Health database if they have a record of CLL therapy, and elderly patients have been observed to be less likely than younger patients to receive CLL treatment (26).

For information on race, the different data collection approaches result in expected differences in completeness and in population distribution across the three databases. The proportion of incomplete records for race in Flatiron Health data is greater than in the other databases. During routine oncology care in the US (i.e., in the source clinics for Flatiron Health), collection of race data is not mandatory or incentivized; on the other hand, the registries feeding both SEER and NPCR have a mandate to reach certain levels of completeness for this variable and thus, this information is collected both directly from self-reports and indirectly using algorithms (27). In addition, Flatiron Health relies on self-reported information by patients, which adds complexity and variability to an information category that is in constant evolution within a broader social context. For example, while “Hispanic or Latino” is standardly recorded as an ethnicity value, in some patients this may be coded as a race value instead; in such cases, these values were mapped to “Unknown”. These challenges probably contribute to the apparent lower completeness of this variable in the Flatiron Health databases. Furthermore, due to the purposeful design of the SEER program, there is an overrepresentation of certain groups compared to the US census (24, 28), a finding consistent with prior representativeness studies (29–31).

Lastly, a combination of differences in sources and in data collection approaches across the three data sources leads to noticeable differences in their information about AJCC disease stage at diagnosis. In some diseases, particularly in HCC, malignant pleural mesothelioma, RCC, SCLC, prostate cancer, and DLBCL, detailed AJCC disease stage information is missing from Flatiron Health databases to a substantially larger extent than in SEER, although that incompleteness is mitigated in the simplified category metastatic/non-metastatic disease. To understand that finding, it is important to note that Flatiron Health data are generated from a pipeline of medical oncology EHR-derived data, where stage information is mostly as documented in unstructured notes by the treating oncology team. In contrast, registries rely on multiple sites of care as sources, and disease stage is intentionally entered into their databases via mandated calculation and coding by trained tumor registrars (32, 33). Ultimately, these fundamental differences lead to idiosyncratic fluctuations in information completeness in EHR-derived vs systematically-collected data. For instance, clinical scenarios where medical oncologists tend to be involved in initial diagnosis (when staging takes place) are more likely to have a complete capture of initial staging in the medical oncology EHR. To wit, compare cancers commonly diagnosed at an advanced stage (e.g., SCLC) or that are eligible for systemic adjuvant therapy from early stages (e.g., breast cancer) with diseases where the medical oncologist tends to be less involved in initial diagnosis (e.g., RCC, which is often managed surgically upon initial diagnosis).

Completeness/incompleteness of EHR-derived data is also affected by practical patterns of clinical documentation; as staging algorithms become increasingly complex, practitioners may tend to be more attentive to staging specifics in settings where the link between staging and treatment is more crucial (i.e., local or locally-advanced settings, where patients are candidates for multi-modality therapy), and less stringent in other settings (i.e., advanced disease) where staging information is less critical to clinical decision making. For example, AJCC staging is not a key consideration for initial HCC treatment decisions; clinical and laboratory data to assess underlying liver functional status and inform potential transplant eligibility are far more clinically relevant. Treating clinicians may prefer to document and rely on clinically actionable, non-AJCC staging systems for the routine management of some diseases, like HCC and SCLC, resulting in less AJCC-based information available in those databases.

In conclusion, the disease-specific Flatiron Health databases provide deep demographic, clinical, and treatment data models derived from EHR information. Several of the data elements in the Flatiron Health databases cannot be found in SEER or NPCR, such as date of metastatic diagnosis and sites of metastatic disease, comprehensive standard biomarker status, longitudinal treatment sequences, and disease progression dates. Within the portfolio of data elements commonly found across the three databases, comparing Flatiron Health to SEER and NPCR shows that Flatiron Health has a regional distribution closer to the general US census than SEER, less complete racial information, and disease-dependent variability in the capture of staging data. These differences stem from the originating EHR-source and from the rules for data capture and processing. Investigators should consider these inter-database demographic differences when designing studies and interpreting results obtained with Flatiron Health data, and when contextualizing their findings relative to SEER-or NPCR-based research.

## Data Availability

The data that support the findings of this study have been originated by Flatiron Health, Inc. Requests for data sharing by license or by permission for the specific purpose of replicating results in this manuscript can be submitted to.

## Acknowledgements

Authors wish to thank Julia Saiz, PhD, Cody Patton, Hannah Gilham, and Jennifer Swanson from Flatiron Health, for editorial support, and Neil McQuarrie from Flatiron Health, for analytical support. We also wish to thank Mary Elizabeth O’Neil, MPH, from the CDC DCPC; Angela Mariotto, PhD, and Donna R. Rivera, PharmD, MSc, from the DCCPS at the NIH NCI; and Olivier Humblet, ScD, Emily Castellanos, MD, MPH, and Roxanne Diaz, BS, from Flatiron Health, for their review and valuable feedback.

## APPENDIX I

**Table A1.** Overview of diagnostic and histology codes used to define eligibility, and of additional entry criteria and definitions of ‘metastatic at diagnosis’.

| Tumor type | Flatiron Health Databases (EDMs) | SEER | NPCR |
| --- | --- | --- | --- |
| Advanced urothelial cancer | ICD-9 188x, 189.1, 189.2, 189.3, or ICD-10 C65x, C66x, C67x, C68.0<br>Metastatic definition: Stage IV, M1<br>Non-Metastatic: M0 or non-stage IV (including unknown AJCC group stage if MO is known)<br>Entered into database if two clinical encounters on or after January 1 2011. | Primary Site = "659-680"<br>Metastatic definition: Summary stage 2000 = "Distant";<br>Non-Metastatic: Summary stage 2000 = "in situ", "Local" or "Regional" | Primary Site - Labeled = "C65.9 - C68.0"<br>Metastatic definition: Summary stage 2000 = "Distant";<br>Non-Metastatic: Summary stage 2000 = "In situ", "Local" or "Regional" |
| Metastatic breast cancer | ICD-9: 174.x or 175.x or ICD-10 C50.x<br>Metastatic definition: Stage IV, M1<br>Non-Metastatic: M0 or non-stage IV (including unknown AJCC group stage if MO is known).<br>Entered into database if two clinical encounters on or after January 1 2011.<br>ICD-9: 174.x or 175.x or ICD-10 C50.x | Primary Site = "500-509"<br>Metastatic: Summary stage 2000 = "Distant";<br>Non-Metastatic: Summary stage 2000 = "Local" or "Regional" | Primary Site - Labeled = "C50.0 - C50.9"<br>Metastatic: Summary stage 2000 = "Distant";<br>Non-Metastatic: Summary stage 2000 = "Local" or "Regional" |
| Early breast cancer |  |  | Primary Site - Labeled = "C50.0 - C50.9"<br>Metastatic: Summary stage 2000 = "Distant";<br>Non-Metastatic: Summary stage 2000 = "Local" or "Regional" |
| Chronic lymphocytic leukemia | ICD-9: 204.1x or ICD-10: C91.1x, C83.0x<br>Metastatic definition: all stages<br>Entered into database if two clinical encounters on or after January 1 2011. | Site recode ICD-O-3/WHO 2008 = "Chronic Lymphocytic Leukemia" AND Histologic type ICD-O-3 = "9823"<br>Metastatic definition: Summary stage | Site recode ICD-O-3/WHO 2008 = "Chronic Lymphocytic Leukemia" AND Histologic type ICD-O-3 = "9823"<br>Metastatic definition: Summary stage |
|  |  | = "Distant" | = "Distant" |
| Metastatic colorectal cancer | ICD-9 153.x or 154.x or ICD-10 C18x, or C19x, or C20x, or C21x<br>Metastatic definition: Stage IV, M1<br>Non-Metastatic: M0 or non-stage IV (including unknown AJCC group stage if MO is known)<br>Entered into database if two clinical encounters on or after January 1 2013. | Primary Site = "180-218"<br>Metastatic: Summary stage 2000 = "Distant";<br>Non-Metastatic: Summary stage 2000 = "Local" or "Regional" | Primary Site - Labeled = "C18.0 - C21.8"<br>Metastatic: Summary stage 2000 = "Distant";<br>Non-Metastatic: Summary stage 2000 = "Local" or "Regional" |
| Diffuse large B-cell lymphoma | ICD 9: 200x, 202x; ICD 10: C82x, C83x, C84x, C85x, C86x, C88x, C96x<br>Metastatic definition: Stage IV or Stage III<br>Non-Metastatic: Stage I or II<br>Entered into database if two clinical encounters on or after January 1 2011. | Histologic type ICD-O-3 = "9680"<br>Metastatic definition: Summary stage 2000 = "Distant";<br>Non-Metastatic: Summary stage 2000 = "Local" or "Regional" | Histologic type ICD-O-3 = "9680"<br>Metastatic definition: Summary stage 2000 = "Distant";<br>Non-Metastatic: Summary stage 2000 = "Local" or "Regional" |
| Follicular lymphoma | ICD 9: 200x, 202x; ICD 10: C82x, C83x, C84x, C85x, C86x, C88x, C96x<br>Metastatic: Stage IV or Stage III<br>Non-Metastatic: Stage I or II<br>Entered into database if two clinical encounters on or after January 1 2011. | Histologic type ICD-O-3 = "9690, 9691, 9695, 9698"<br>Metastatic: Summary stage 2000 = "Distant";<br>Non-Metastatic: Summary stage 2000 = "Local" or "Regional" | Histologic type ICD-O-3 = "9690, 9691, 9695, 9698"<br>Metastatic: Summary stage 2000 = "Distant";<br>Non-Metastatic: Summary stage 2000 = "Local" or "Regional" |
| Advanced gastric/esophageal carcinoma | ICD-9: 151.x, 150.x,<br>ICD-9 C16.x, C15.x<br>Metastatic definition: Stage IV, M1<br>Non-Metastatic: M0 or non-stage IV (including unknown AJCC group stage if MO is known)<br>Entered into database if two clinical encounters on or after January 1 2011. | Primary Site = "150-169"<br>Metastatic: Summary stage 2000 = "Distant";<br>Non-Metastatic: Summary stage 2000 = "Local" or "Regional" | Primary Site - Labeled = "C15.0 - C16.9"<br>Metastatic: Summary stage 2000 = "Distant";<br>Non-Metastatic: Summary stage 2000 = "Local" or "Regional" |
| Hepatocellular carcinoma | ICD-9: 155.x OR ICD-10 C22.x<br>Metastatic: Stage IV (not otherwise specified) or Stage IVB, M1<br>Non-Metastatic: M0 or non-stage IV/IVB (including unknown AJCC group stage if MO is known)<br>Entered into database if two clinical encounters on or after January 1 2011. | Primary Site = "220, 221" AND Histologic type ICD-O-3 = "8170, 8171, 8172, 8173, 8174, 8175"<br>Metastatic: Summary stage 2000 = "Distant";<br>Non-Metastatic: Summary stage 2000 = "Local" or "Regional" | Primary Site - Labeled = "C22.0, C22.1" AND Histologic type ICD-O-3 = "8170-8175"<br>Metastatic: Summary stage 2000 = "Distant";<br>Non-Metastatic: Summary stage 2000 = "Local" or "Regional" |
| Advanced head and neck cancer | 140x, 141x, 143x, 144x, 145x, 146x, 147x, 148x, 149x, 161x; ICD 10: C00x, C01x, C02x, C03x, C04x, C05x, C06x, C09x, C10x, C11x, C12x, C13x, C14x, C32x<br>Metastatic definition: Stage IVC, M1<br>Non-Metastatic: M0 or non-stage IVC (including unknown AJCC group stage if MO is known and IVA, IVB)<br>Entered into database if two clinical encounters on or after January 1 2011. | Primary Site = "0-69, 90-148, 320-329"<br>Metastatic: Summary stage 2000 = "Distant";<br>Non-Metastatic: Summary stage 2000 = "Local" or "Regional" | Primary Site - Labeled = "C00.0 - C06.9, C09.0 - C14.8, C32.0 - C32.9"<br>Metastatic: Summary stage 2000 = "Distant";<br>Non-Metastatic: Summary stage 2000 = "Local" or "Regional" |
| Advanced melanoma | ICD-9 172.x or ICD-10 C43x or D03x<br>Metastatic: Stage IV, M1<br>Non-Metastatic: M0 or non-stage IV (including unknown AJCC group stage if MO is known)<br>Entered into database if two clinical encounters on or after January 1 2011. | Site recode ICD-O-3/WHO 2008 = "Melanoma of the Skin"<br>Met: Summary stage 2000 = "Distant";<br>Non-Met: Summary stage 2000 = "Local" or "Regional" | Site recode ICD-O-3/WHO 2008 = "Melanoma of the skin"<br>Met: Summary stage 2000 = "Distant";<br>Non-Met: Summary stage 2000 = "Local" or "Regional" |
| Malignant pleural mesothelioma | ICD-9:163* or ICD-10 C45.0<br>Metastatic: Stage IV, M1<br>Non-Metastatic: M0 or non-stage IV (including unknown AJCC group stage if MO is known)<br>Entered into database if two clinical<br>encounters on or after January 1 2011. | Site recode ICD-O-3/WHO 2008 = "Mesothelioma" AND Histologic Type 9050-9053<br>Metastatic: Summary stage 2000 = "Distant";<br>Non-Metastatic: Summary stage 2000 = "Local" or "Regional" | Site recode ICD-O-3/WHO 2008 = "Mesothelioma" AND Histologic Type 9050-9053<br>Metastatic: Summary stage 2000 = "Distant";<br>Non-Metastatic: Summary stage 2000 = "Local" or "Regional" |
| Multiple myeloma | ICD-9 203.0x or ICD-10 C90.0x, C90<br>Metastatic: all known stages.<br>Entered into database if two clinical encounters on or after January 1 2011. | Site recode ICD-O-3/WHO 2008 = "Myeloma"<br>Met: Summary stage = "Distant" | Site recode ICD-O-3/WHO 2008 = "Myeloma"<br>Met: Summary stage = "Distant" |
| Advanced non-small cell lung cancer (NSCLC) | ICD-9 162.x or ICD-10 C34x, or C39.9<br>Metastatic: Stage IV, M1<br>Non-Metastatic: M0 or non-stage IV (including unknown AJCC group stage if MO is known)<br>Entered into database if two clinical encounters on or after January 1 2011. | Primary Site = "340-349, 390-399"<br>AND Histologic type ICD-O-3 = "8046, 8033, 8022, 8012, 8980, 8140, 8560, 8070, 8550, 8250, 8251, 8252, 8253, 8254, 8255, 8310, 8470, 8083, 8052, 8084, 8071, 8072, 8073, 8480, 8481, 8260, 8490, 8230, 8012, 8013, 8014, 8082, 8123, 8310"<br>Met: Summary stage 2000 = "Distant";<br>Non-Met: Summary stage 2000 = "Local" or "Regional" | Primary Site - Labeled = "C34.0 - C34.9, C39.9" AND Histologic type ICD-O-3 = "8046, 8033, 8022, 8012, 8980, 8140, 8560, 8070, 8550, 8250, 8251, 8252, 8253, 8254, 8255, 8310, 8470, 8083, 8052, 8084, 8071, 8072, 8073, 8480, 8481, 8260, 8490, 8230, 8012, 8013, 8014, 8082, 8123, 8310"<br>Met: Summary stage 2000 = "Distant";<br>Non-Met: Summary stage 2000 = "Local" or "Regional" |
| Ovarian carcinoma | ICD 9: 183x, 158x; ICD 10: C56x, C57.0x, C48x<br>Metastatic: Stage IV, M1<br>Non-Metastatic: M0 or non-stage IV (including unknown AJCC group stage if MO is known)<br>Entered into database if two clinical encounters on or after January 1 2011. | Primary Site = "569, 570, 480-488"<br>Metastatic: Summary stage 2000 = "Distant";<br>Non-Metastatic: Summary stage 2000 = "Local" or "Regional" | Primary Site - Labeled = "C48.0 - C48.8, C56.9, C57.0"<br>Metastatic: Summary stage 2000 = "Distant";<br>Non-Metastatic: Summary stage 2000 = "Local" or "Regional" |
| Metastatic pancreatic carcinoma | ICD-9 157.x; ICD-10 C25.x<br>Metastatic: Stage IV, M1<br>Non-Metastatic: M0 or non-stage IV (including unknown AJCC group stage if MO is known)<br>Entered into database if two clinical<br>encounters on or after January 1 2014. | Primary Site = "250-259"<br>Metastatic: Summary stage 2000 = "Distant";<br>Non-Metastatic: Summary stage 2000 = "Local" or "Regional" | Primary Site - Labeled = "C25.0 - C25.9"<br>Metastatic: Summary stage 2000 = "Distant";<br>Non-Metastatic: Summary stage 2000 = "Local" or "Regional" |
| Metastatic prostate cancer | ICD 9 code: 185x<br>ICD 10 code: C61x<br>Metastatic: Stage IV, M1<br>Non-Metastatic: M0 or non-stage IV<br>(including unknown AJCC group stage if MO is known)<br>Entered into database if two clinical encounters on or after January 1 2013. | Primary Site = "619"<br>Metastatic: Summary stage 2000 = "Distant";<br>Non-Metastatic: Summary stage 2000 = "Local" or "Regional" | Primary Site - Labeled = "C61.9"<br>Metastatic: Summary stage 2000 = "Distant";<br>Non-Metastatic: Summary stage 2000 = "Local" or "Regional" |
| Metastatic renal-cell carcinoma | ICD-9 189.x or ICD-10 C64x or C65x<br>Metastatic: Stage IV (excluding patients who are T4NxM0), M1<br>Non-Metastatic: M0, T4 only Stage IV or non-stage IV (including unknown AJCC group stage if MO is known)<br>Entered into database if two clinical encounters on or after January 1 2011. | Primary Site = "649, 659"<br>Metastatic: Summary stage 2000 = "Distant";<br>Non-Metastatic: Summary stage 2000 = "Local" or "Regional" | Primary Site - Labeled = "C64.9, C65.9"<br>Metastatic: Summary stage 2000 = "Distant";<br>Non-Metastatic: Summary stage 2000 = "Local" or "Regional" |
| Small-cell lung cancer (SCLC) | ICD-9 162.x or ICD-10 C34x, or C39.9<br>Metastatic: Extensive stage and stage IV or M1<br>Non-Metastatic: limited stage or M0 or non-Stage IV (including AJCC group stage if MO is unknown)<br>Entered into database if two clinical encounters on or after January 1 2013. | Primary Site = "340-349, 390-399" and Histologic type ICD-O-3 = "8041-8045"<br>Metastatic: Summary stage 2000 = "Distant";<br>Non-Metastatic: Summary stage 2000 = "Local" or "Regional" | Primary Site - Labeled = "C34.0 - C34.9, C39.9" AND Histologic type ICD-O-3 = "8041- 8045"<br>Metastatic: Summary stage 2000 = "Distant";<br>Non-Metastatic: Summary stage 2000 = "Local" or "Regional" |

**Table A2.** Overview of variable definitions and steps taken to align variables across the different databases.

|  | Definitions |  |  |
| --- | --- | --- | --- |
|  | SEER | NPCR | Flatiron Health |
| Age at initial diagnosis |  |  |  |
| Region | Assigned as: West, Northeast, South, Midwest, other according to US census definitions. |  |  |
| Race | Defined as:<br>• Asian = "Asian" or "Pacific Islander"<br>• Black or African American = "Black"<br>• Other Race = "American Indian" or "Alaska Native"<br>• Unknown = "Unknown"<br>• White = "White" |  | As self-reported by patients, and captured into the EHR. Defined as:<br>• White = "White"<br>• Black or African American = "Black or African American"<br>• Asian = "Asian"<br>• Other Race = "Other"<br>• Unknown = "Unknown", "Hispanic or Latino"<br>Categories had to be combined to harmonize comparisons across databases |
| Stage at diagnosis | Defined as:<br>• Derived AJCC Stage Group, 7th ed for diagnosis before 2016.<br>• Derived SEER Cmb Stg Grp for diagnosis in 2016. |  | As reported by the treating clinician in the EHR, following AJCC criteria, 7th or 8th edition. Or as noted by a Flatiron abstractor via assessment of T, N, M components, using AJCC 7th ed guidelines until July, 2018 and AJCC 8th ed guidelines thereafter. |

**Table A3.**
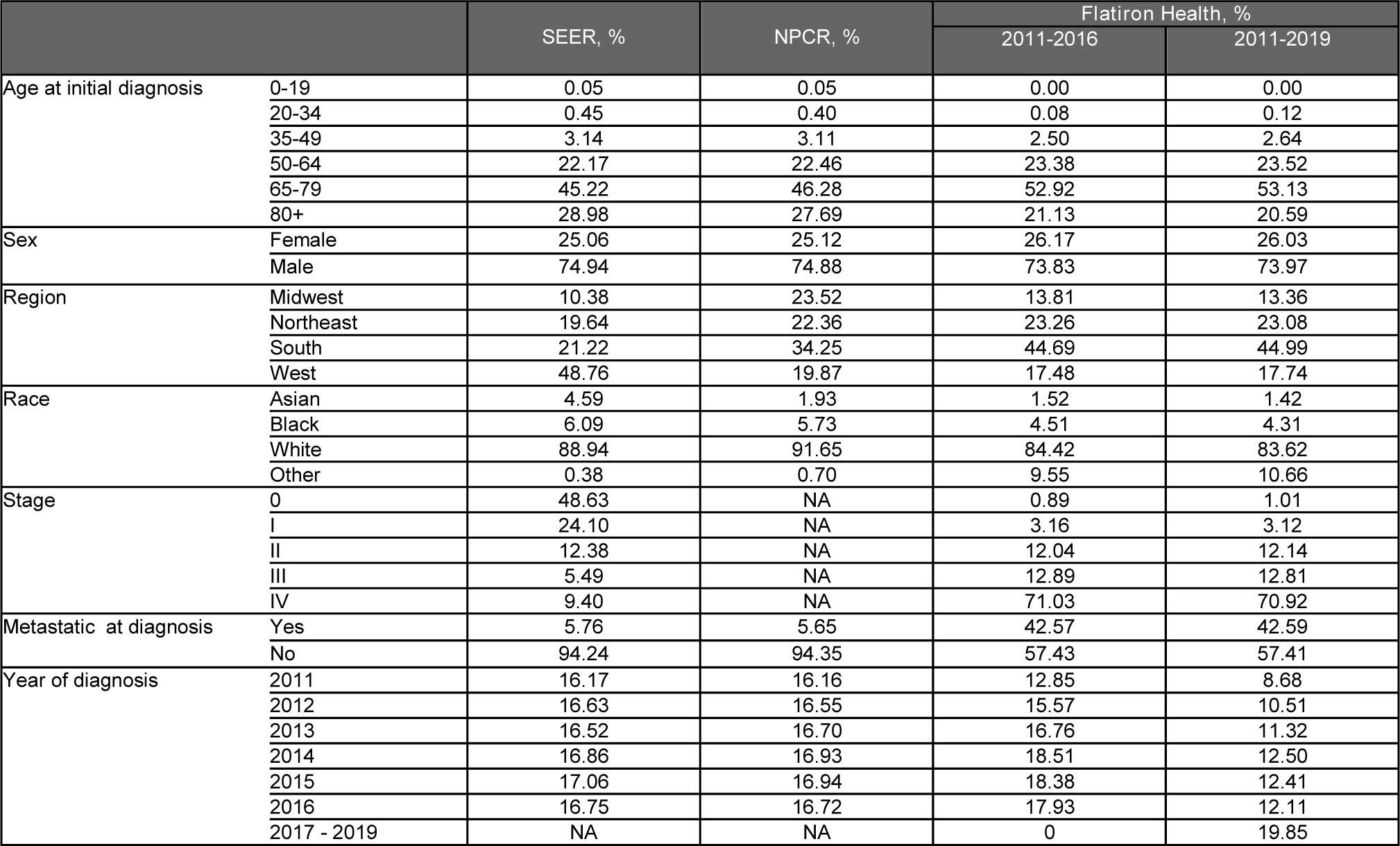
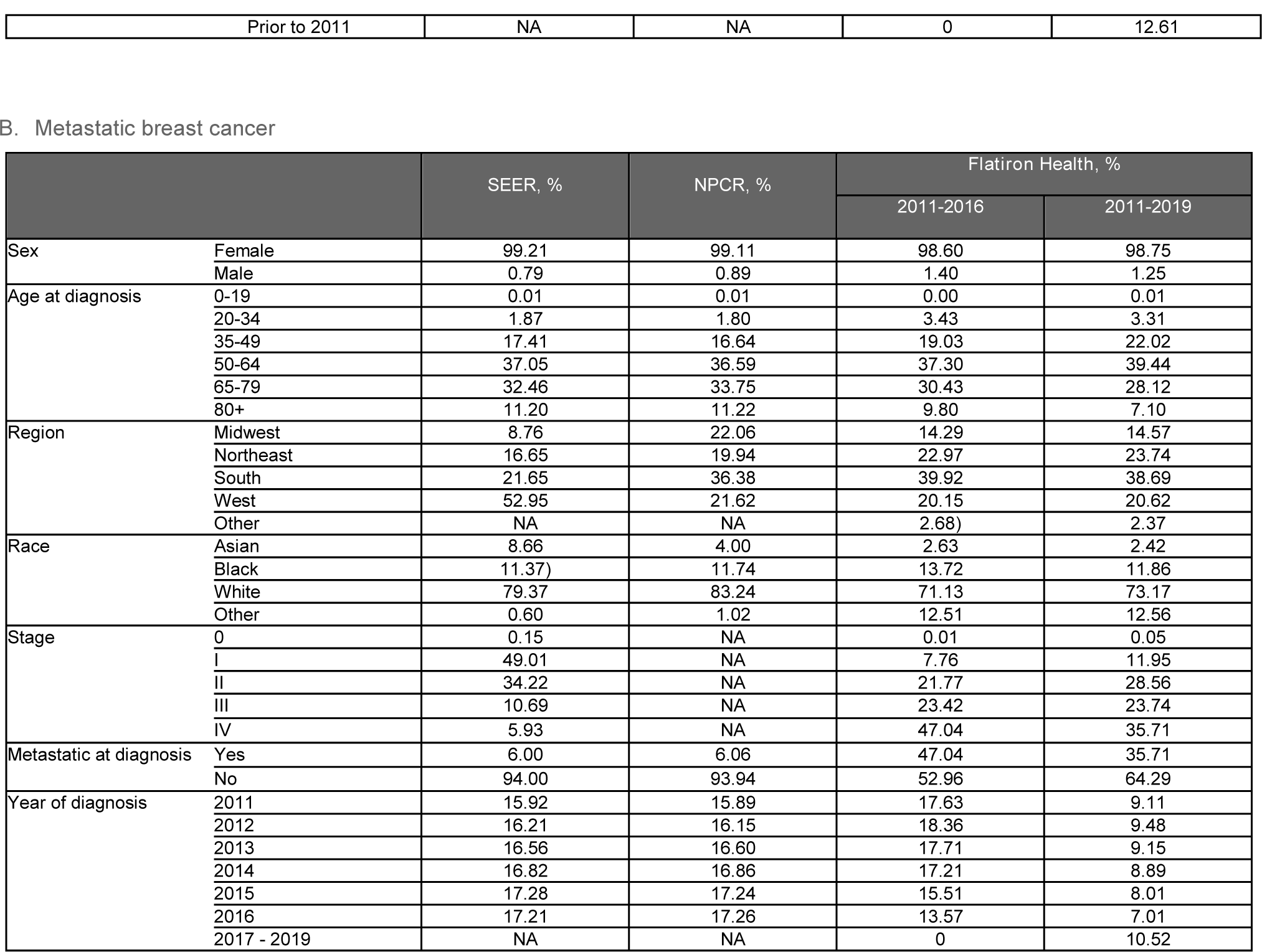
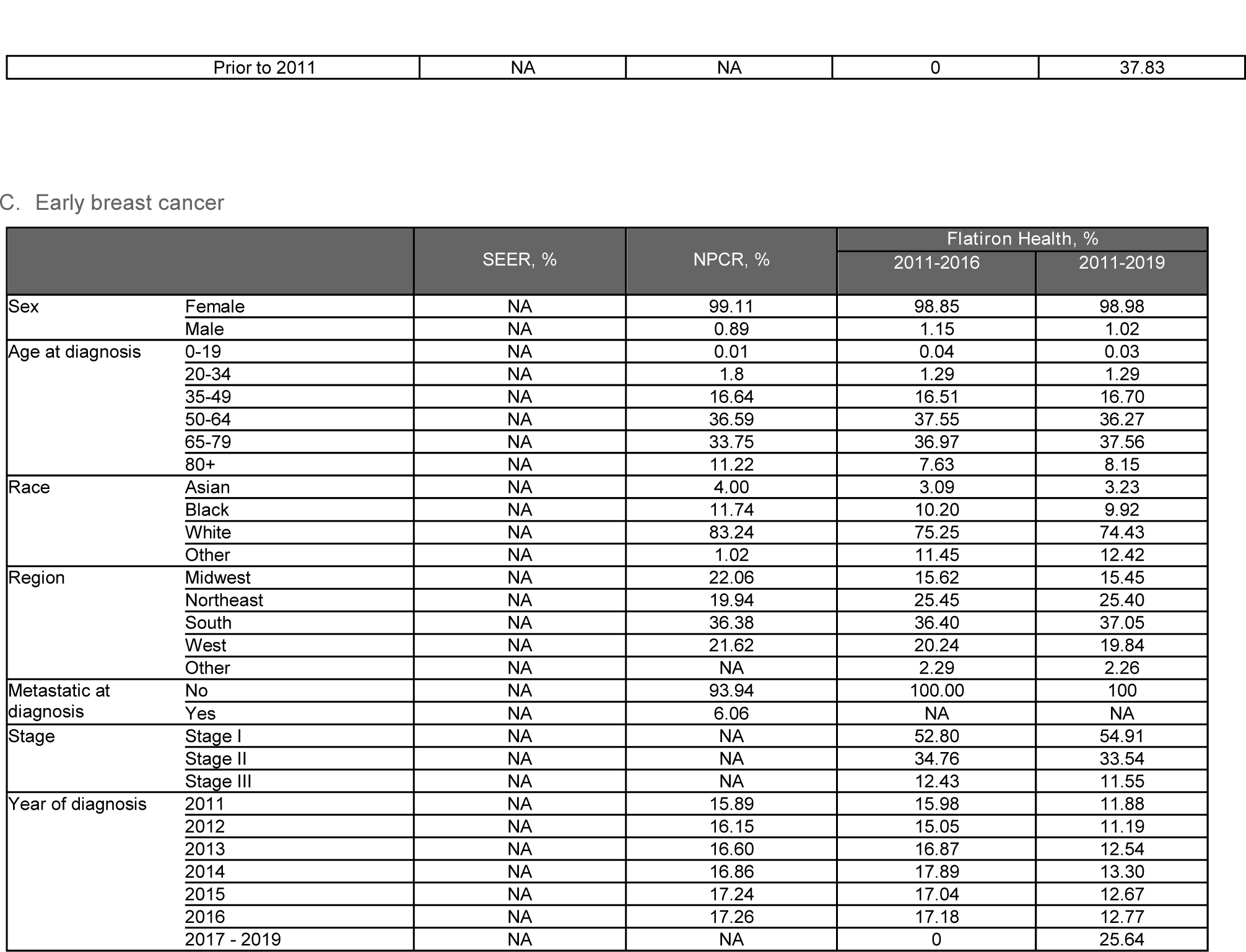
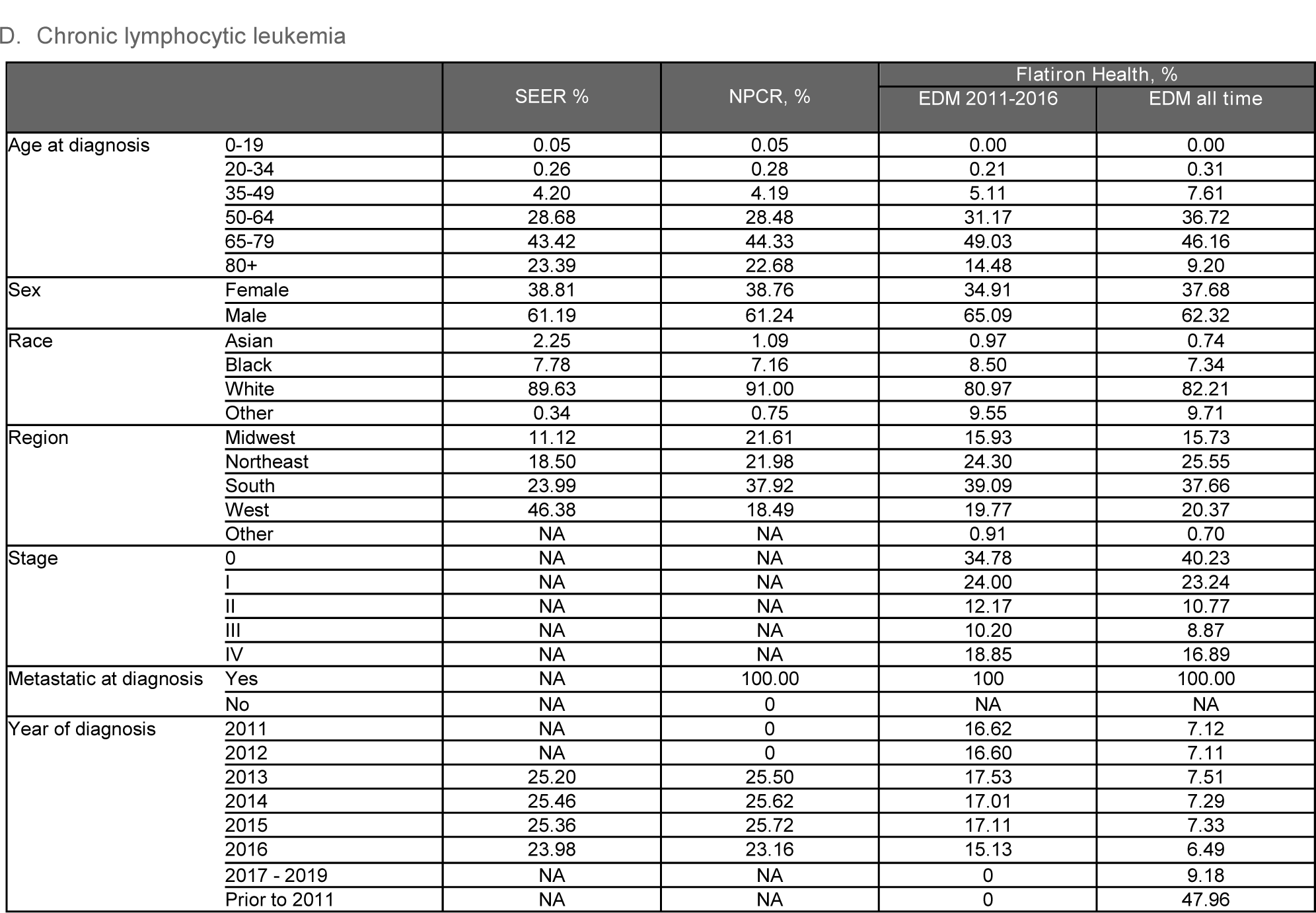
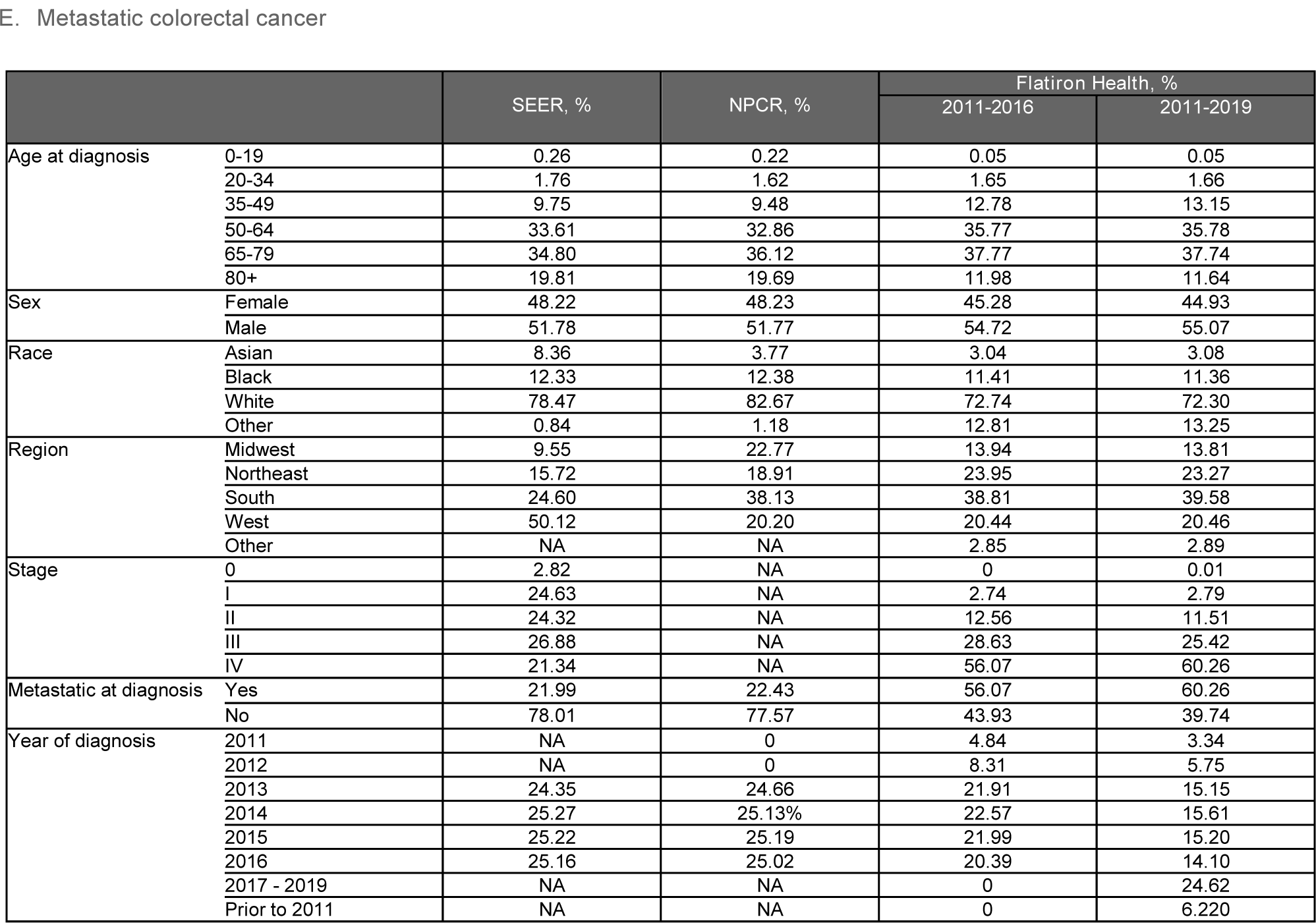
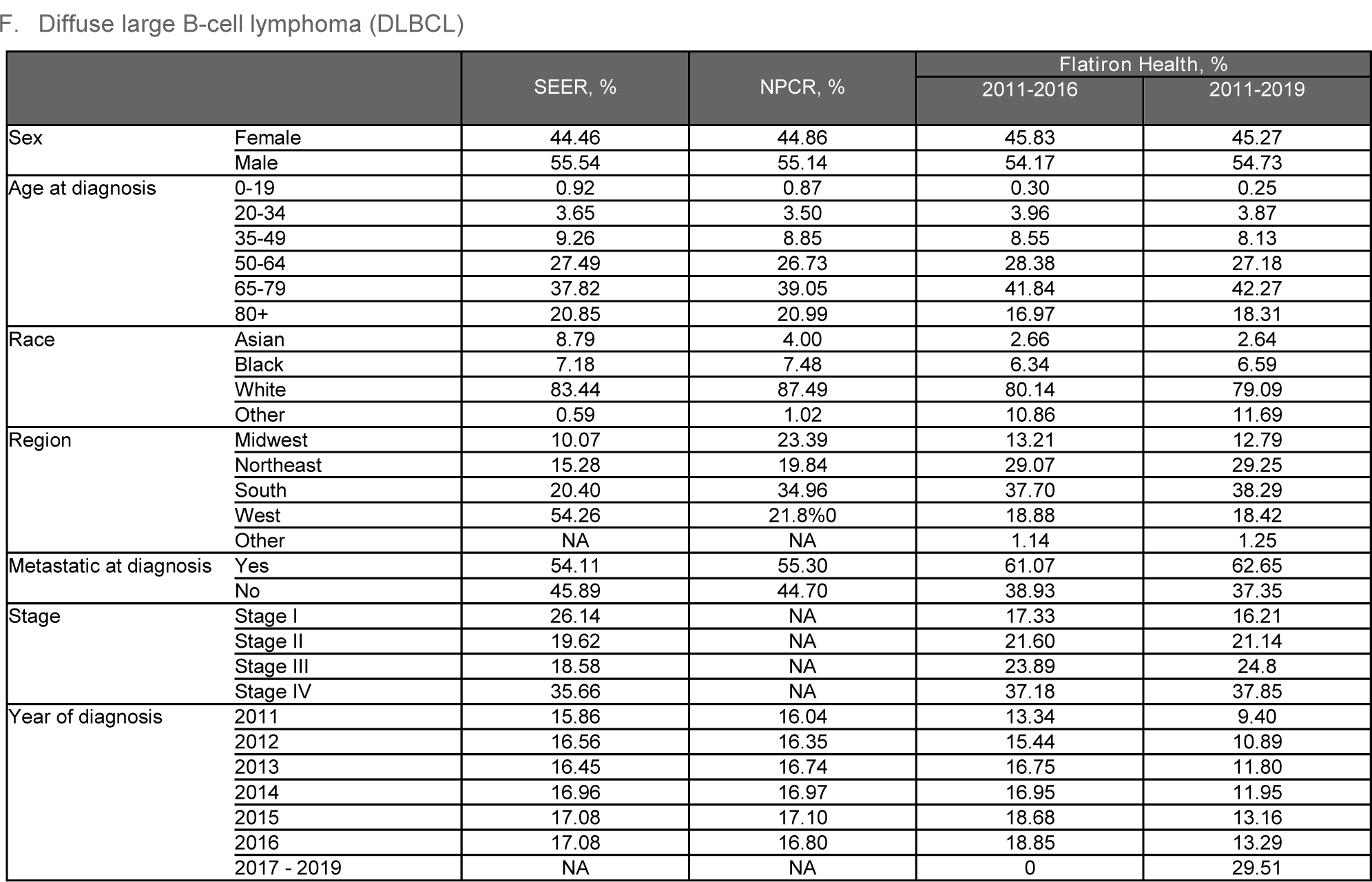
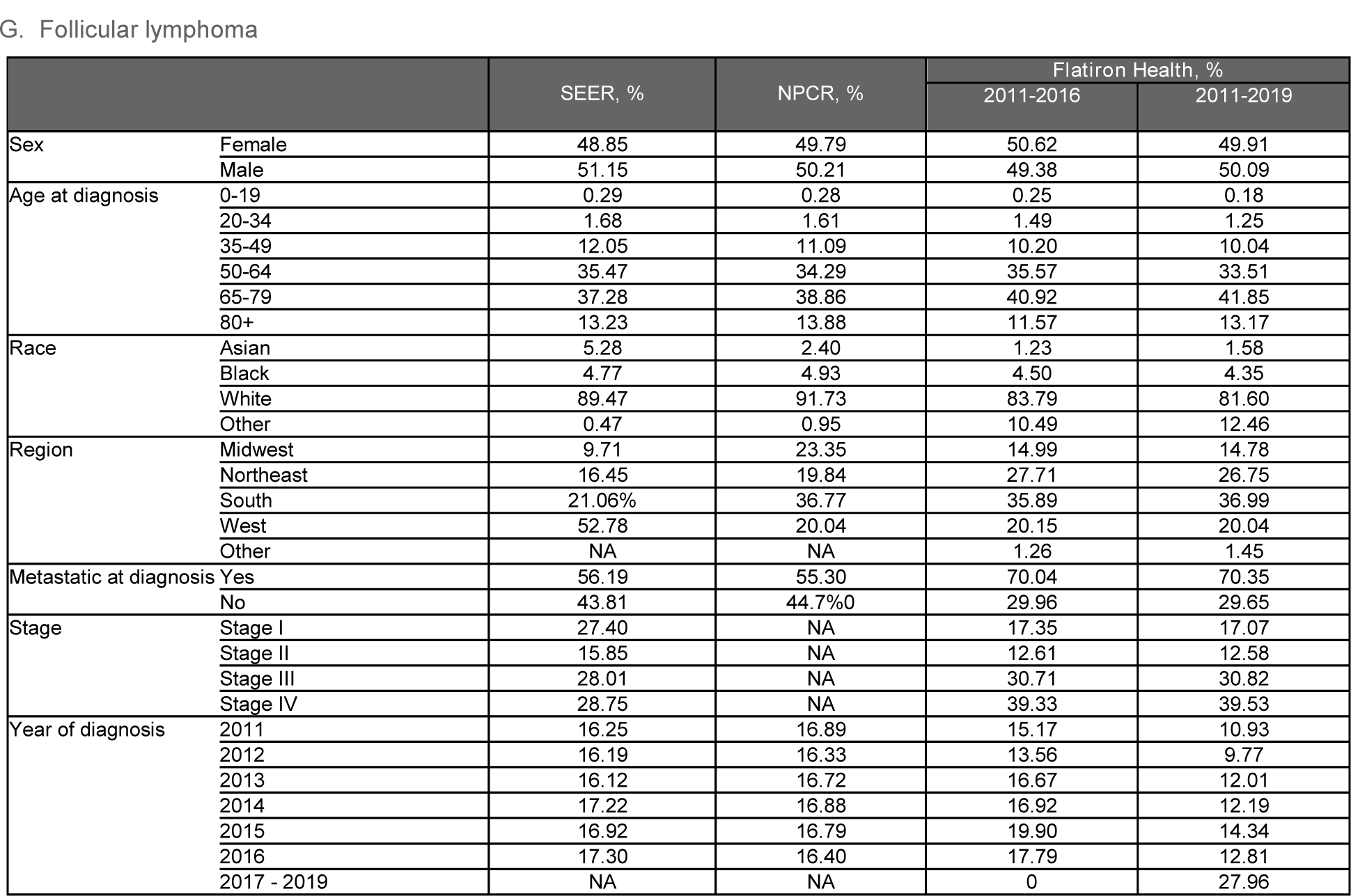
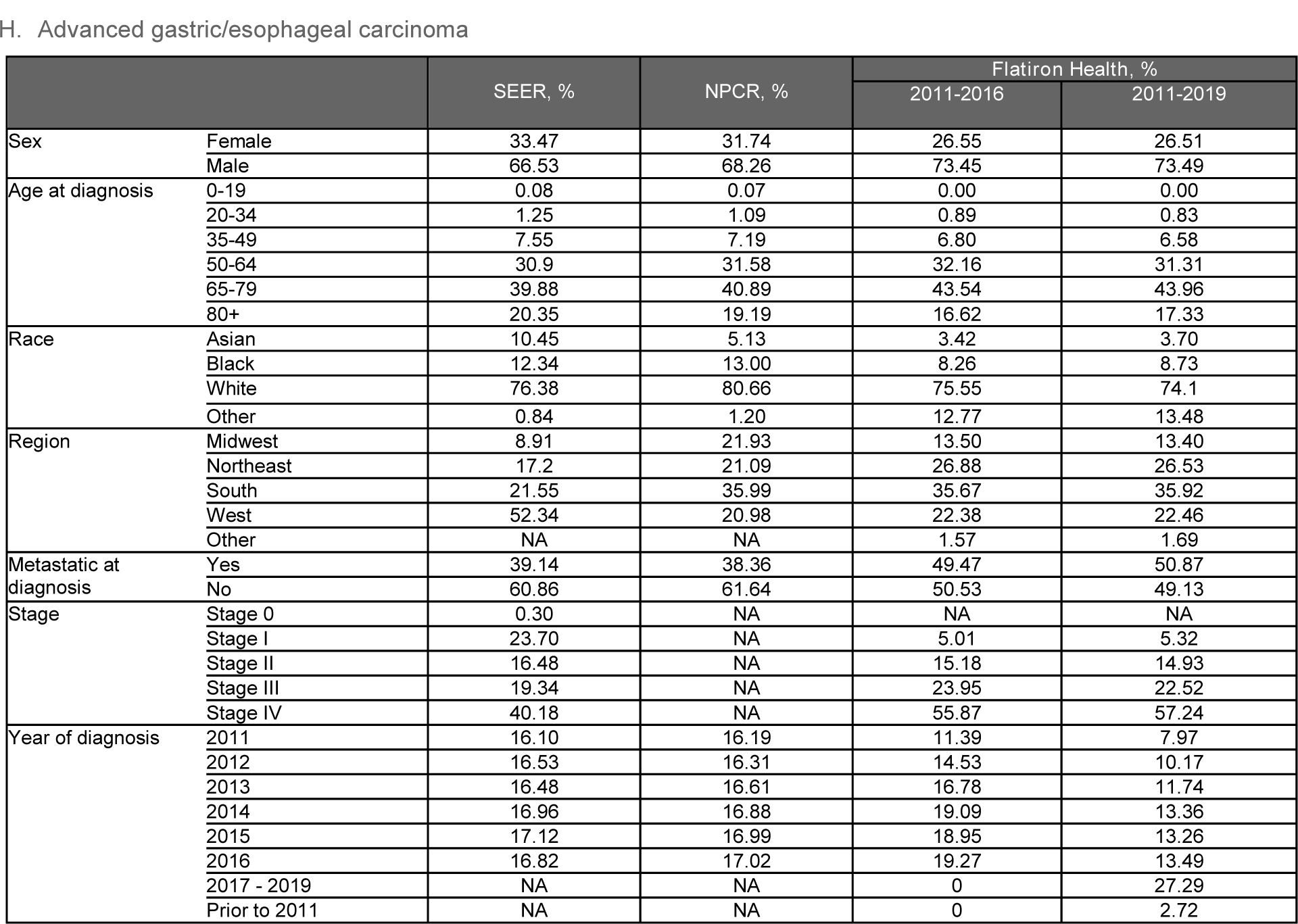
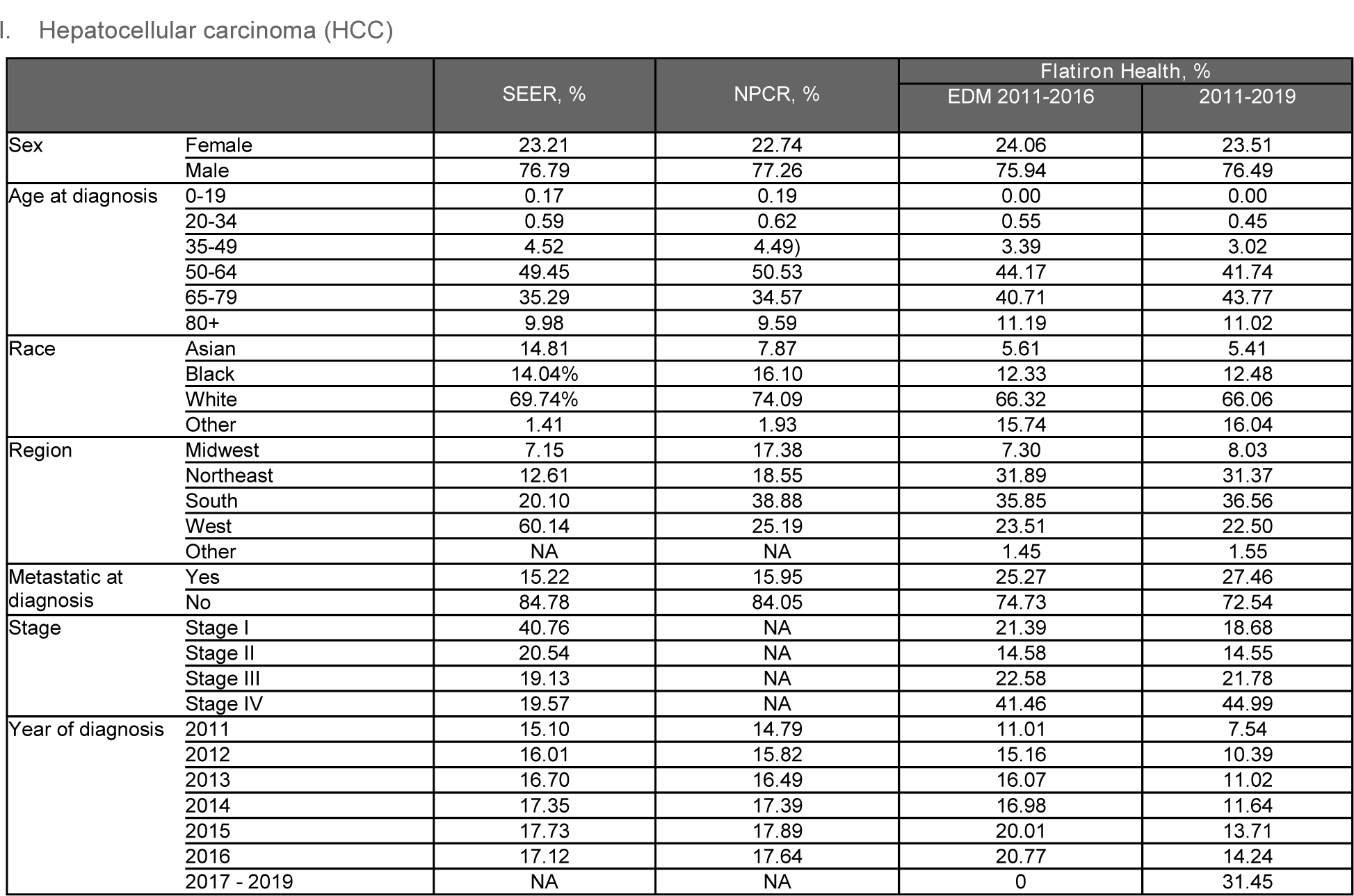
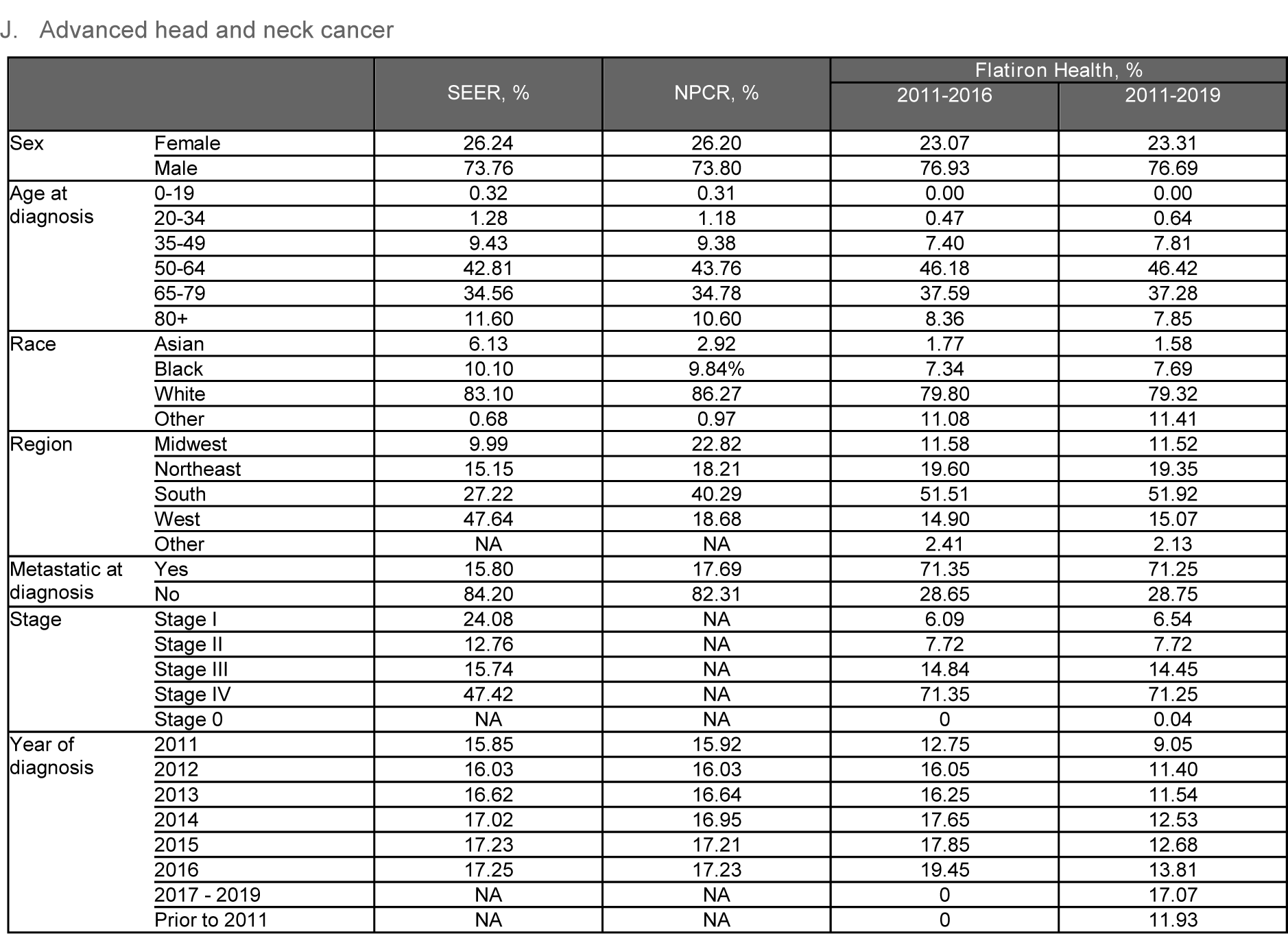
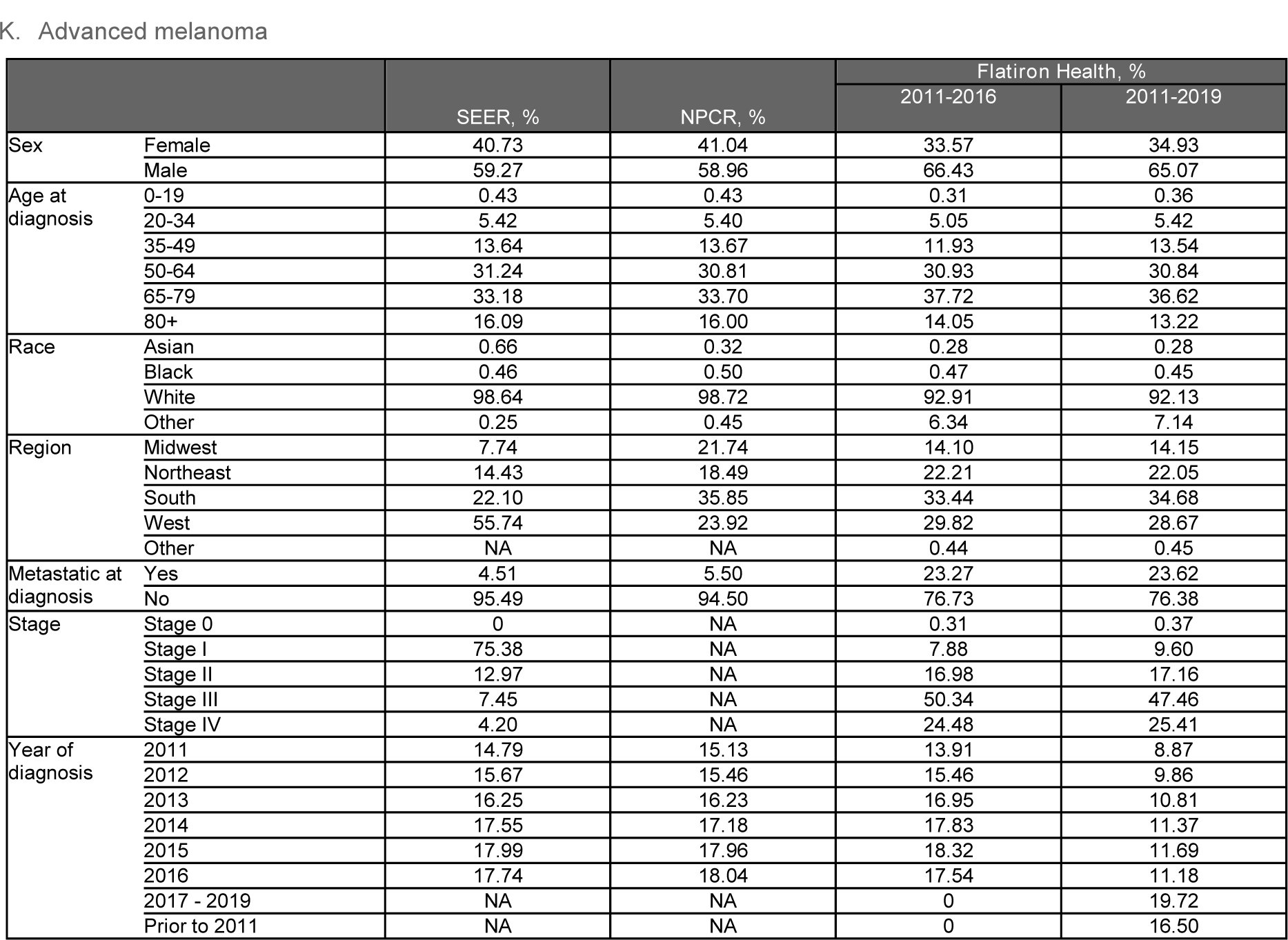
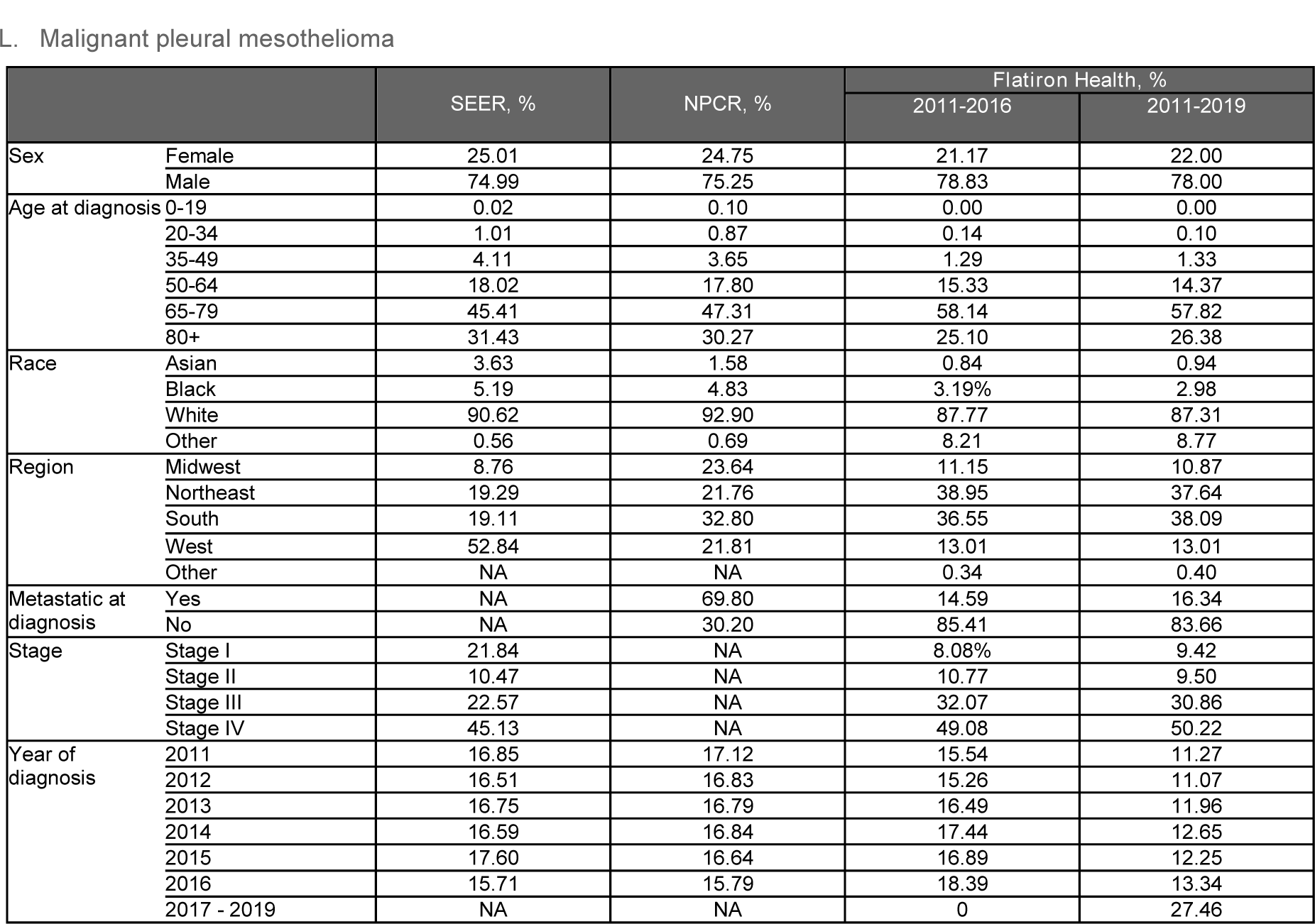
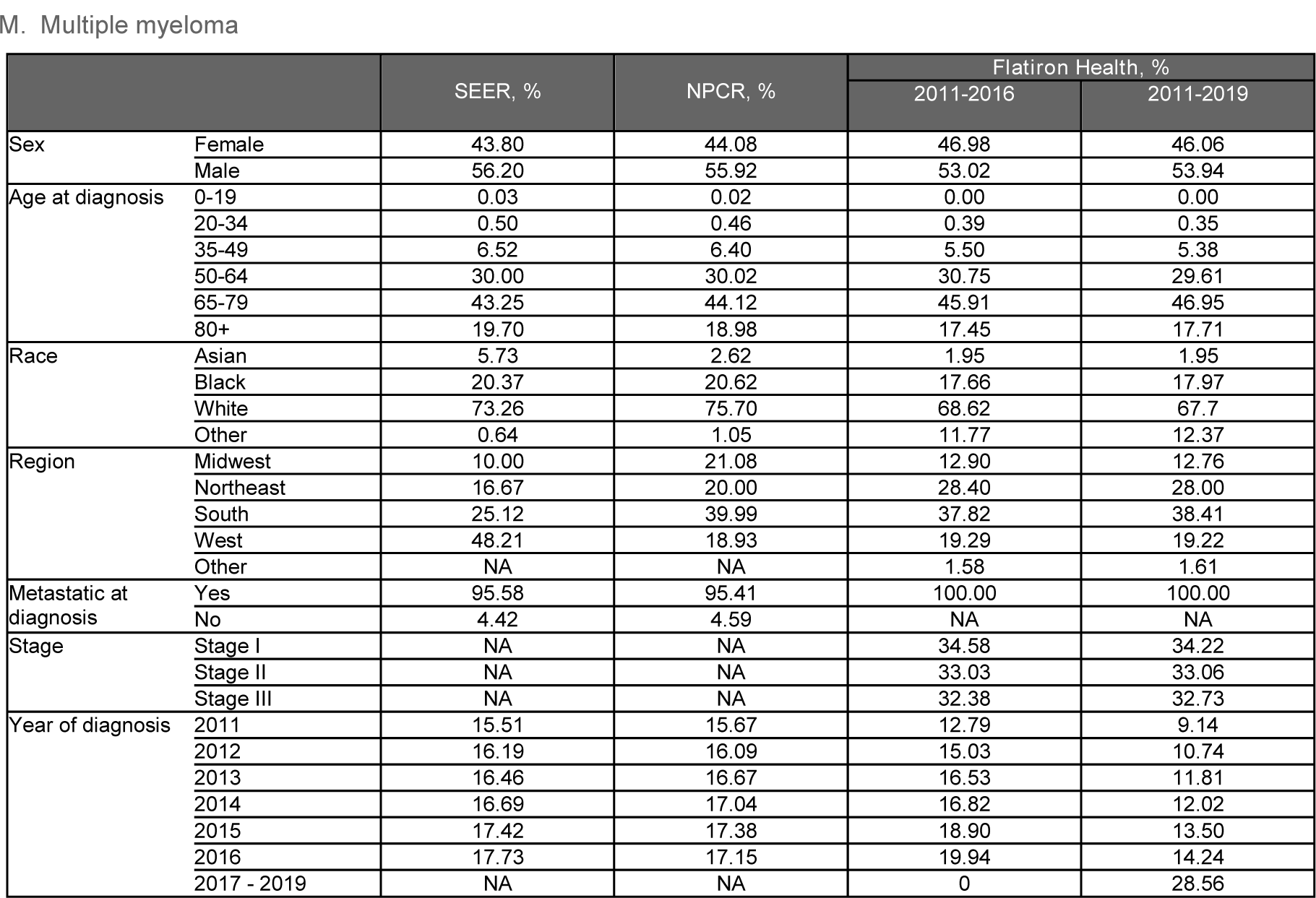
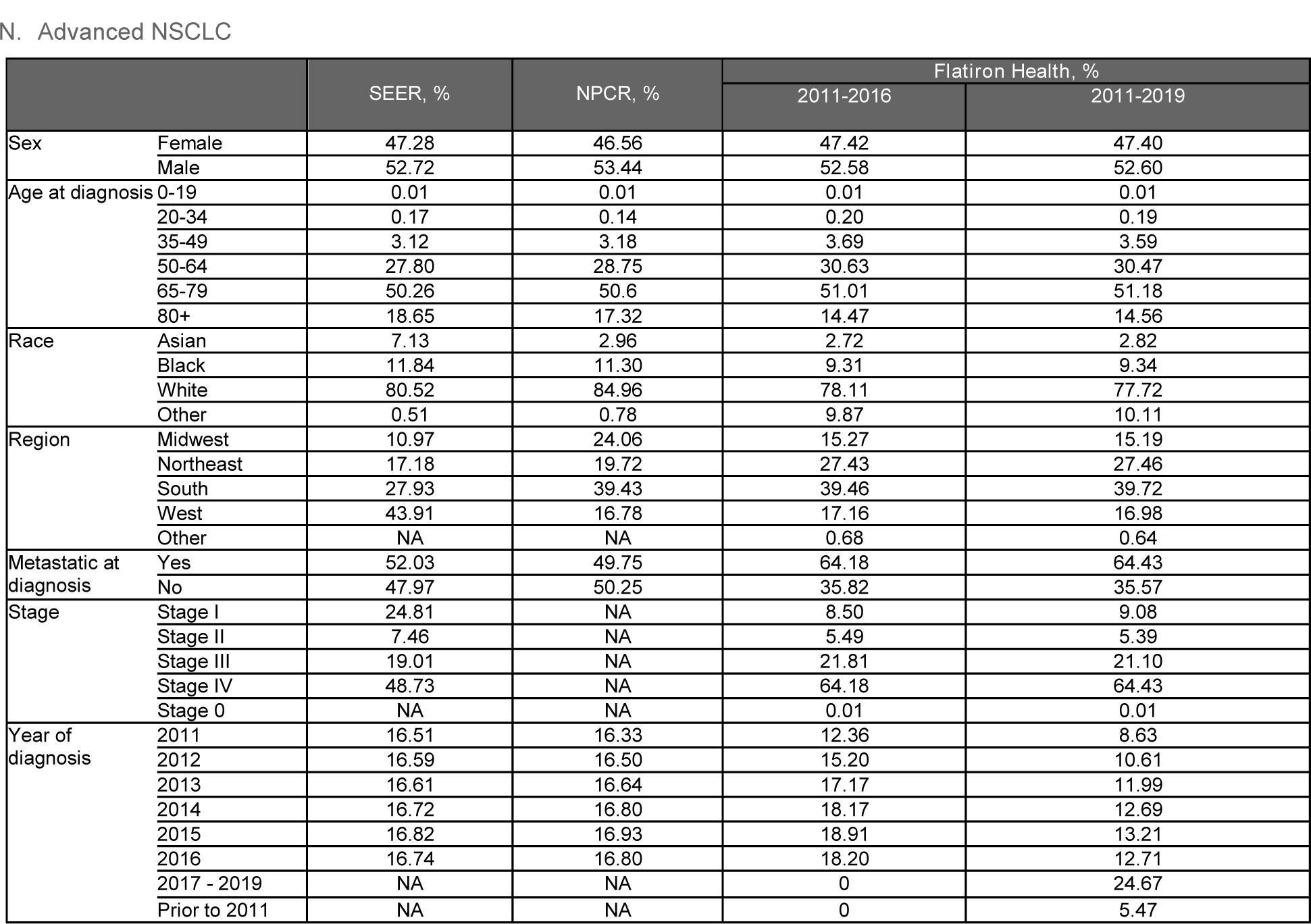
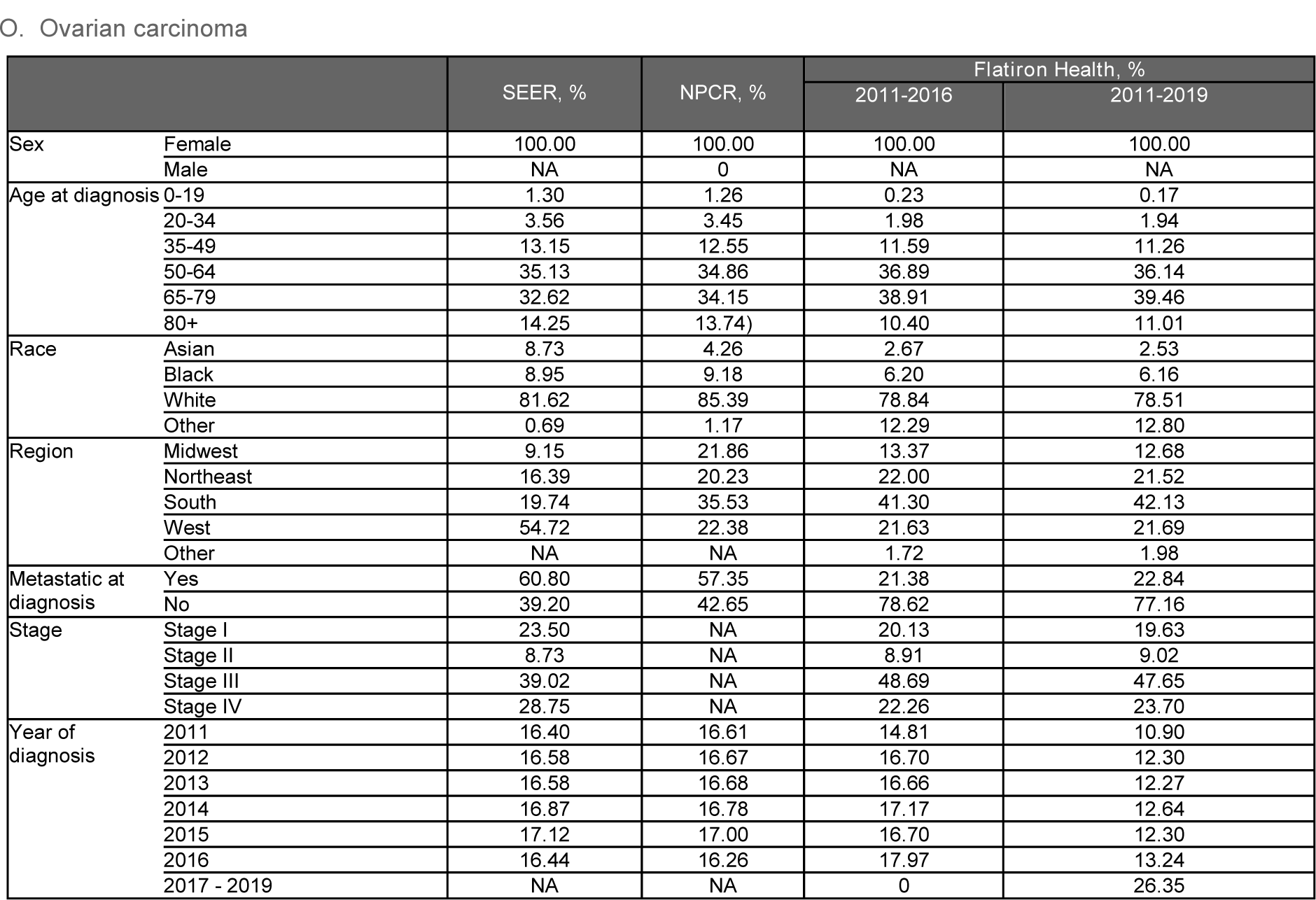
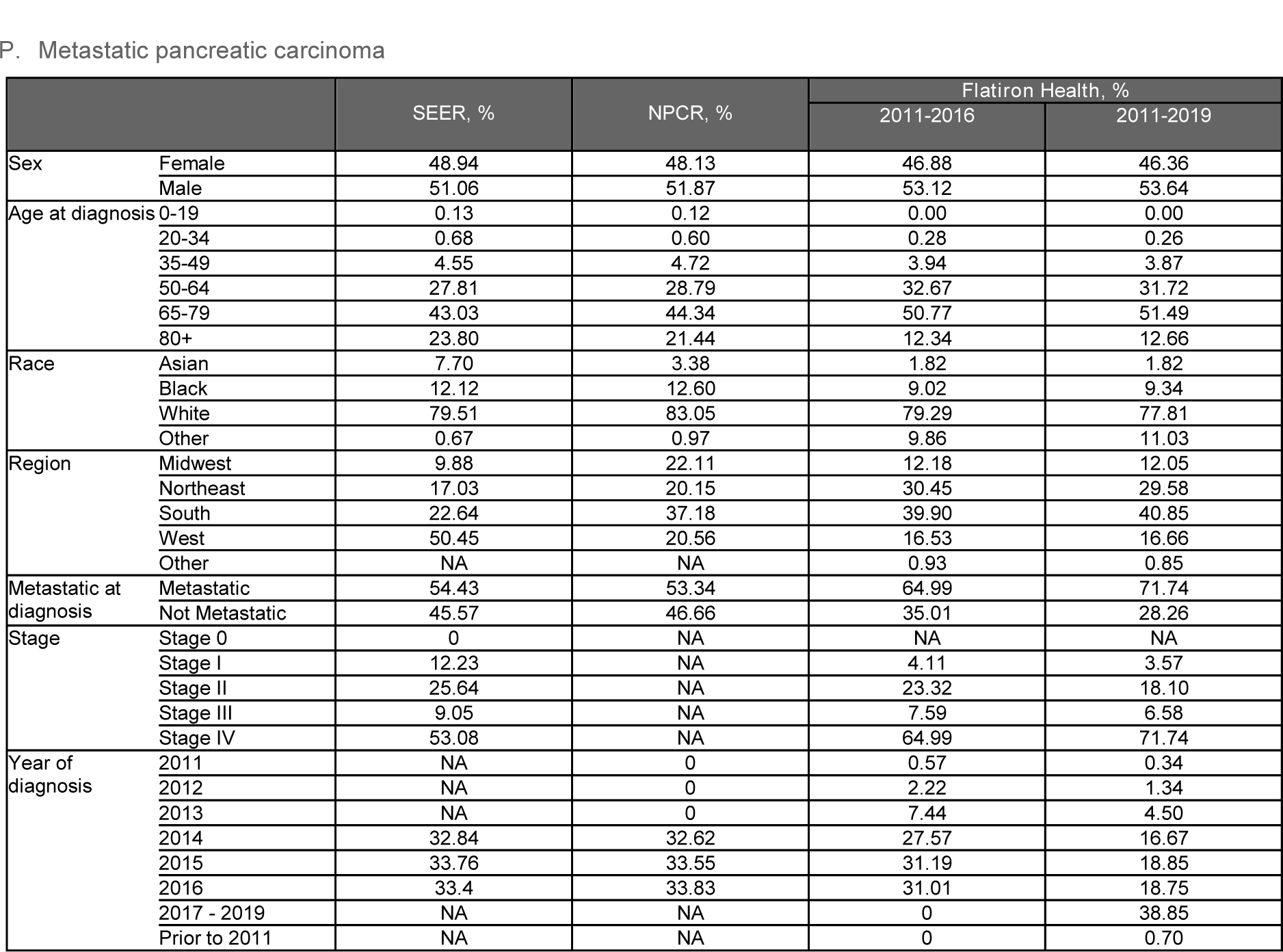
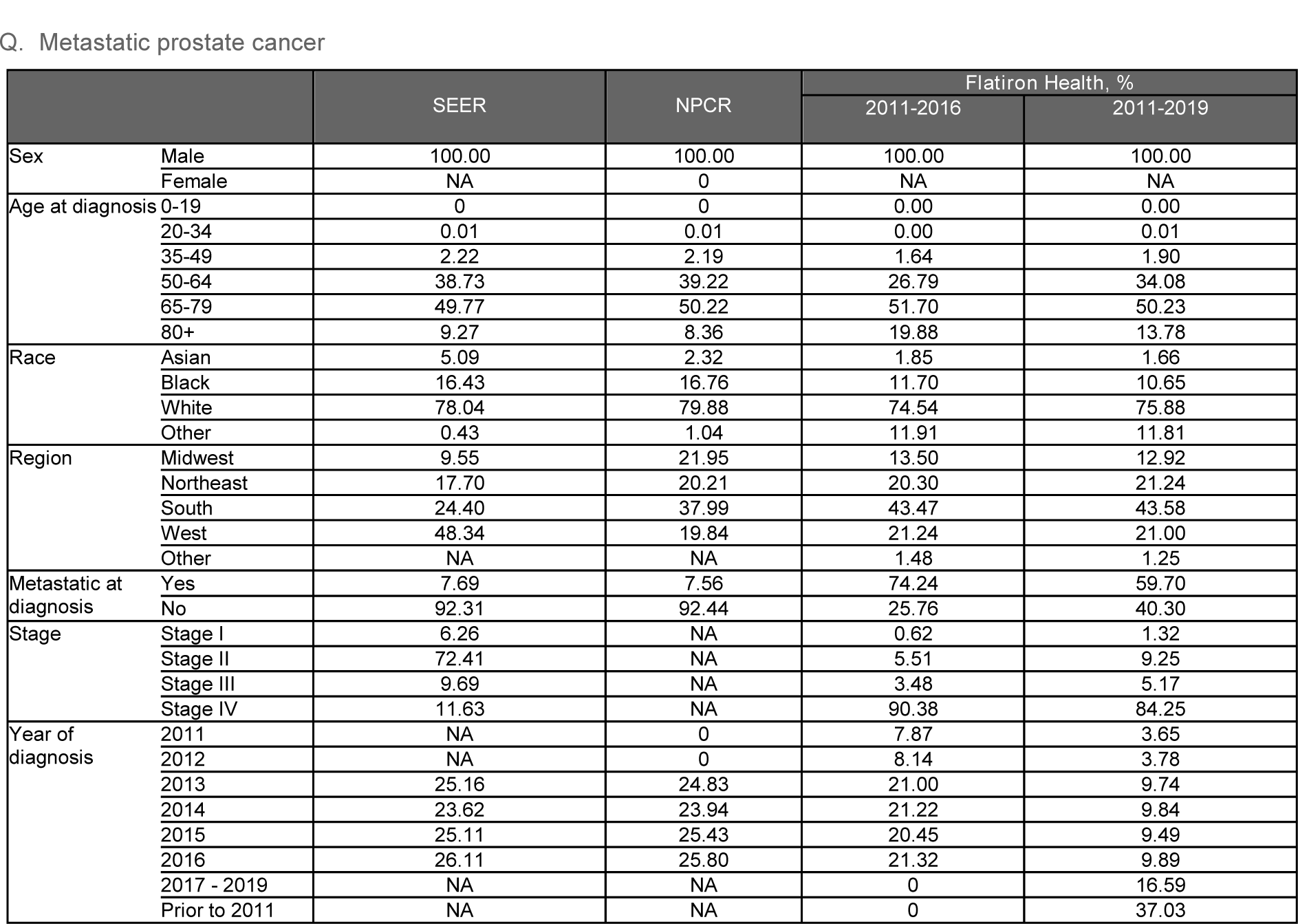
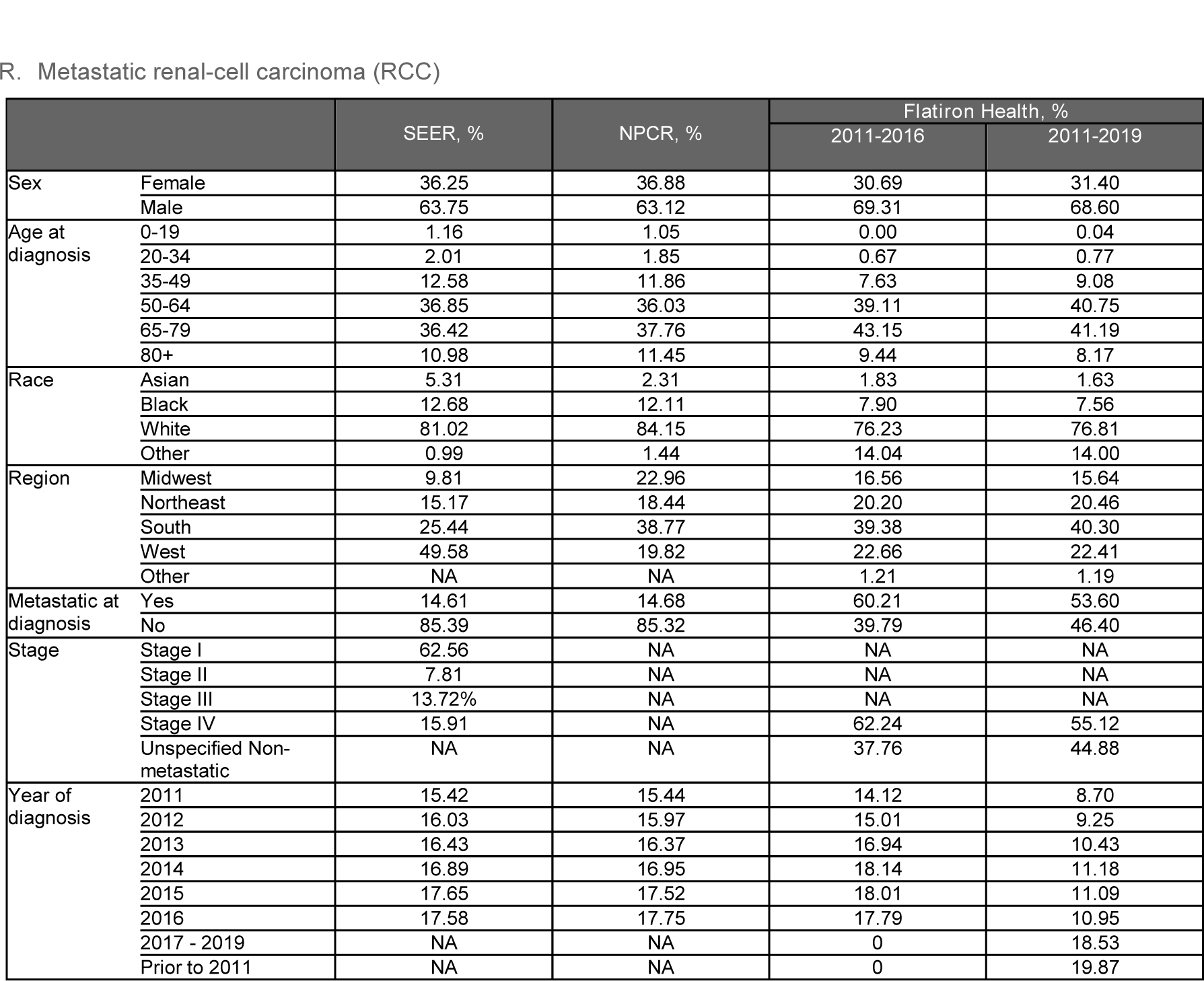
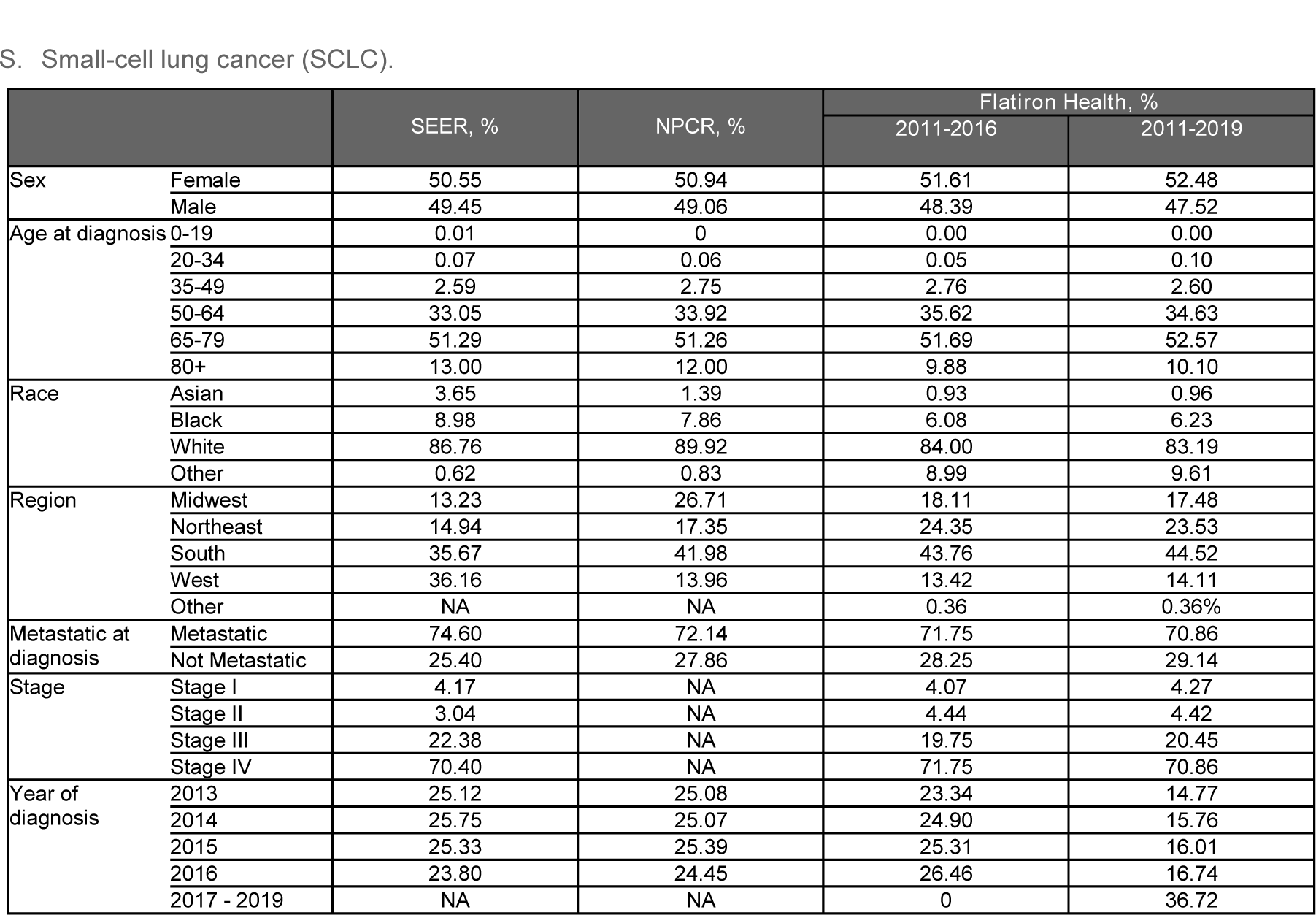
Distribution of patient characteristics excluding patients with unknown/missing status across the categories evaluated. **Tables A3.A-S**: NA = not available from the source data, or numbers under the reporting suppression value to preserve patient confidentiality.

|  |  | SEER, % | NPCR, % | Flatiron Health, % |  |
| --- | --- | --- | --- | --- | --- |
|  |  |  |  | 2011-2016 | 2011-2019 |
| Age at initial diagnosis | 0-19 | 0.05 | 0.05 | 0.00 | 0.00 |
|  | 20-34 | 0.45 | 0.40 | 0.08 | 0.12 |
|  | 35-49 | 3.14 | 3.11 | 2.50 | 2.64 |
|  | 50-64 | 22.17 | 22.46 | 23.38 | 23.52 |
|  | 65-79 | 45.22 | 46.28 | 52.92 | 53.13 |
|  | 80+ | 28.98 | 27.69 | 21.13 | 20.59 |
| Sex | Female | 25.06 | 25.12 | 26.17 | 26.03 |
|  | Male | 74.94 | 74.88 | 73.83 | 73.97 |
| Region | Midwest | 10.38 | 23.52 | 13.81 | 13.36 |
|  | Northeast | 19.64 | 22.36 | 23.26 | 23.08 |
|  | South | 21.22 | 34.25 | 44.69 | 44.99 |
|  | West | 48.76 | 19.87 | 17.48 | 17.74 |
| Race | Asian | 4.59 | 1.93 | 1.52 | 1.42 |
|  | Black | 6.09 | 5.73 | 4.51 | 4.31 |
|  | White | 88.94 | 91.65 | 84.42 | 83.62 |
|  | Other | 0.38 | 0.70 | 9.55 | 10.66 |
| Stage | 0 | 48.63 | NA | 0.89 | 1.01 |
|  | I | 24.10 | NA | 3.16 | 3.12 |
|  | II | 12.38 | NA | 12.04 | 12.14 |
|  | III | 5.49 | NA | 12.89 | 12.81 |
|  | IV | 9.40 | NA | 71.03 | 70.92 |
| Metastatic at diagnosis | Yes | 5.76 | 5.65 | 42.57 | 42.59 |
|  | No | 94.24 | 94.35 | 57.43 | 57.41 |
| Year of diagnosis | 2011 | 16.17 | 16.16 | 12.85 | 8.68 |
|  | 2012 | 16.63 | 16.55 | 15.57 | 10.51 |
|  | 2013 | 16.52 | 16.70 | 16.76 | 11.32 |
|  | 2014 | 16.86 | 16.93 | 18.51 | 12.50 |
|  | 2015 | 17.06 | 16.94 | 18.38 | 12.41 |
|  | 2016 | 16.75 | 16.72 | 17.93 | 12.11 |
|  | 2017 - 2019 | NA | NA | 0 | 19.85 |

|  |  |  |  |  |
| --- | --- | --- | --- | --- |
| Prior to 2011 | NA | NA | 0 | 12.61 |

B. Metastatic breast cancer
|  |  | SEER, % | NPCR, % | Flatiron Health, % |  |
| --- | --- | --- | --- | --- | --- |
|  |  |  |  | 2011-2016 | 2011-2019 |
| Sex | Female | 99.21 | 99.11 | 98.60 | 98.75 |
|  | Male | 0.79 | 0.89 | 1.40 | 1.25 |
| Age at diagnosis | 0-19 | 0.01 | 0.01 | 0.00 | 0.01 |
|  | 20-34 | 1.87 | 1.80 | 3.43 | 3.31 |
|  | 35-49 | 17.41 | 16.64 | 19.03 | 22.02 |
|  | 50-64 | 37.05 | 36.59 | 37.30 | 39.44 |
|  | 65-79 | 32.46 | 33.75 | 30.43 | 28.12 |
|  | 80+ | 11.20 | 11.22 | 9.80 | 7.10 |
| Region | Midwest | 8.76 | 22.06 | 14.29 | 14.57 |
|  | Northeast | 16.65 | 19.94 | 22.97 | 23.74 |
|  | South | 21.65 | 36.38 | 39.92 | 38.69 |
|  | West | 52.95 | 21.62 | 20.15 | 20.62 |
|  | Other | NA | NA | 2.68) | 2.37 |
| Race | Asian | 8.66 | 4.00 | 2.63 | 2.42 |
|  | Black | 11.37) | 11.74 | 13.72 | 11.86 |
|  | White | 79.37 | 83.24 | 71.13 | 73.17 |
|  | Other | 0.60 | 1.02 | 12.51 | 12.56 |
| Stage | 0 | 0.15 | NA | 0.01 | 0.05 |
|  | I | 49.01 | NA | 7.76 | 11.95 |
|  | II | 34.22 | NA | 21.77 | 28.56 |
|  | III | 10.69 | NA | 23.42 | 23.74 |
|  | IV | 5.93 | NA | 47.04 | 35.71 |
| Metastatic at diagnosis | Yes | 6.00 | 6.06 | 47.04 | 35.71 |
|  | No | 94.00 | 93.94 | 52.96 | 64.29 |
| Year of diagnosis | 2011 | 15.92 | 15.89 | 17.63 | 9.11 |
|  | 2012 | 16.21 | 16.15 | 18.36 | 9.48 |
|  | 2013 | 16.56 | 16.60 | 17.71 | 9.15 |
|  | 2014 | 16.82 | 16.86 | 17.21 | 8.89 |
|  | 2015 | 17.28 | 17.24 | 15.51 | 8.01 |
|  | 2016 | 17.21 | 17.26 | 13.57 | 7.01 |
|  | 2017 - 2019 | NA | NA | 0 | 10.52 |

|  |  |  |  |  |
| --- | --- | --- | --- | --- |
| Prior to 2011 | NA | NA | 0 | 37.83 |

C. Early breast cancer
|  |  | SEER, % | NPCR, % | Flatiron Health, % |  |
| --- | --- | --- | --- | --- | --- |
|  |  |  |  | 2011-2016 | 2011-2019 |
| Sex | Female | NA | 99.11 | 98.85 | 98.98 |
|  | Male | NA | 0.89 | 1.15 | 1.02 |
| Age at diagnosis | 0-19 | NA | 0.01 | 0.04 | 0.03 |
|  | 20-34 | NA | 1.8 | 1.29 | 1.29 |
|  | 35-49 | NA | 16.64 | 16.51 | 16.70 |
|  | 50-64 | NA | 36.59 | 37.55 | 36.27 |
|  | 65-79 | NA | 33.75 | 36.97 | 37.56 |
|  | 80+ | NA | 11.22 | 7.63 | 8.15 |
| Race | Asian | NA | 4.00 | 3.09 | 3.23 |
|  | Black | NA | 11.74 | 10.20 | 9.92 |
|  | White | NA | 83.24 | 75.25 | 74.43 |
|  | Other | NA | 1.02 | 11.45 | 12.42 |
| Region | Midwest | NA | 22.06 | 15.62 | 15.45 |
|  | Northeast | NA | 19.94 | 25.45 | 25.40 |
|  | South | NA | 36.38 | 36.40 | 37.05 |
|  | West | NA | 21.62 | 20.24 | 19.84 |
|  | Other | NA | NA | 2.29 | 2.26 |
| Metastatic at diagnosis | No | NA | 93.94 | 100.00 | 100 |
|  | Yes | NA | 6.06 | NA | NA |
| Stage | Stage I | NA | NA | 52.80 | 54.91 |
|  | Stage II | NA | NA | 34.76 | 33.54 |
|  | Stage III | NA | NA | 12.43 | 11.55 |
| Year of diagnosis | 2011 | NA | 15.89 | 15.98 | 11.88 |
|  | 2012 | NA | 16.15 | 15.05 | 11.19 |
|  | 2013 | NA | 16.60 | 16.87 | 12.54 |
|  | 2014 | NA | 16.86 | 17.89 | 13.30 |
|  | 2015 | NA | 17.24 | 17.04 | 12.67 |
|  | 2016 | NA | 17.26 | 17.18 | 12.77 |
|  | 2017 - 2019 | NA | NA | 0 | 25.64 |

D. Chronic lymphocytic leukemia
|  |  | SEER % | NPCR, % | Flatiron Health, % |  |
| --- | --- | --- | --- | --- | --- |
|  |  |  |  | EDM 2011-2016 | EDM all time |
| Age at diagnosis | 0-19 | 0.05 | 0.05 | 0.00 | 0.00 |
|  | 20-34 | 0.26 | 0.28 | 0.21 | 0.31 |
|  | 35-49 | 4.20 | 4.19 | 5.11 | 7.61 |
|  | 50-64 | 28.68 | 28.48 | 31.17 | 36.72 |
|  | 65-79 | 43.42 | 44.33 | 49.03 | 46.16 |
|  | 80+ | 23.39 | 22.68 | 14.48 | 9.20 |
| Sex | Female | 38.81 | 38.76 | 34.91 | 37.68 |
|  | Male | 61.19 | 61.24 | 65.09 | 62.32 |
| Race | Asian | 2.25 | 1.09 | 0.97 | 0.74 |
|  | Black | 7.78 | 7.16 | 8.50 | 7.34 |
|  | White | 89.63 | 91.00 | 80.97 | 82.21 |
|  | Other | 0.34 | 0.75 | 9.55 | 9.71 |
| Region | Midwest | 11.12 | 21.61 | 15.93 | 15.73 |
|  | Northeast | 18.50 | 21.98 | 24.30 | 25.55 |
|  | South | 23.99 | 37.92 | 39.09 | 37.66 |
|  | West | 46.38 | 18.49 | 19.77 | 20.37 |
|  | Other | NA | NA | 0.91 | 0.70 |
| Stage | 0 | NA | NA | 34.78 | 40.23 |
|  | I | NA | NA | 24.00 | 23.24 |
|  | II | NA | NA | 12.17 | 10.77 |
|  | III | NA | NA | 10.20 | 8.87 |
|  | IV | NA | NA | 18.85 | 16.89 |
| Metastatic at diagnosis | Yes | NA | 100.00 | 100 | 100.00 |
|  | No | NA | 0 | NA | NA |
| Year of diagnosis | 2011 | NA | 0 | 16.62 | 7.12 |
|  | 2012 | NA | 0 | 16.60 | 7.11 |
|  | 2013 | 25.20 | 25.50 | 17.53 | 7.51 |
|  | 2014 | 25.46 | 25.62 | 17.01 | 7.29 |
|  | 2015 | 25.36 | 25.72 | 17.11 | 7.33 |
|  | 2016 | 23.98 | 23.16 | 15.13 | 6.49 |
|  | 2017 - 2019 | NA | NA | 0 | 9.18 |
|  | Prior to 2011 | NA | NA | 0 | 47.96 |

E. Metastatic colorectal cancer
|  |  | SEER, % | NPCR, % | Flatiron Health, % |  |
| --- | --- | --- | --- | --- | --- |
|  |  |  |  | 2011-2016 | 2011-2019 |
| Age at diagnosis | 0-19 | 0.26 | 0.22 | 0.05 | 0.05 |
|  | 20-34 | 1.76 | 1.62 | 1.65 | 1.66 |
|  | 35-49 | 9.75 | 9.48 | 12.78 | 13.15 |
|  | 50-64 | 33.61 | 32.86 | 35.77 | 35.78 |
|  | 65-79 | 34.80 | 36.12 | 37.77 | 37.74 |
|  | 80+ | 19.81 | 19.69 | 11.98 | 11.64 |
| Sex | Female | 48.22 | 48.23 | 45.28 | 44.93 |
|  | Male | 51.78 | 51.77 | 54.72 | 55.07 |
| Race | Asian | 8.36 | 3.77 | 3.04 | 3.08 |
|  | Black | 12.33 | 12.38 | 11.41 | 11.36 |
|  | White | 78.47 | 82.67 | 72.74 | 72.30 |
|  | Other | 0.84 | 1.18 | 12.81 | 13.25 |
| Region | Midwest | 9.55 | 22.77 | 13.94 | 13.81 |
|  | Northeast | 15.72 | 18.91 | 23.95 | 23.27 |
|  | South | 24.60 | 38.13 | 38.81 | 39.58 |
|  | West | 50.12 | 20.20 | 20.44 | 20.46 |
|  | Other | NA | NA | 2.85 | 2.89 |
| Stage | 0 | 2.82 | NA | 0 | 0.01 |
|  | I | 24.63 | NA | 2.74 | 2.79 |
|  | II | 24.32 | NA | 12.56 | 11.51 |
|  | III | 26.88 | NA | 28.63 | 25.42 |
|  | IV | 21.34 | NA | 56.07 | 60.26 |
| Metastatic at diagnosis | Yes | 21.99 | 22.43 | 56.07 | 60.26 |
|  | No | 78.01 | 77.57 | 43.93 | 39.74 |
| Year of diagnosis | 2011 | NA | 0 | 4.84 | 3.34 |
|  | 2012 | NA | 0 | 8.31 | 5.75 |
|  | 2013 | 24.35 | 24.66 | 21.91 | 15.15 |
|  | 2014 | 25.27 | 25.13% | 22.57 | 15.61 |
|  | 2015 | 25.22 | 25.19 | 21.99 | 15.20 |
|  | 2016 | 25.16 | 25.02 | 20.39 | 14.10 |
|  | 2017 - 2019 | NA | NA | 0 | 24.62 |
|  | Prior to 2011 | NA | NA | 0 | 6.220 |

F. Diffuse large B-cell lymphoma (DLBCL)
|  |  | SEER, % | NPCR, % | Flatiron Health, % |  |
| --- | --- | --- | --- | --- | --- |
|  |  |  |  | 2011-2016 | 2011-2019 |
| Sex | Female | 44.46 | 44.86 | 45.83 | 45.27 |
|  | Male | 55.54 | 55.14 | 54.17 | 54.73 |
| Age at diagnosis | 0-19 | 0.92 | 0.87 | 0.30 | 0.25 |
|  | 20-34 | 3.65 | 3.50 | 3.96 | 3.87 |
|  | 35-49 | 9.26 | 8.85 | 8.55 | 8.13 |
|  | 50-64 | 27.49 | 26.73 | 28.38 | 27.18 |
|  | 65-79 | 37.82 | 39.05 | 41.84 | 42.27 |
|  | 80+ | 20.85 | 20.99 | 16.97 | 18.31 |
| Race | Asian | 8.79 | 4.00 | 2.66 | 2.64 |
|  | Black | 7.18 | 7.48 | 6.34 | 6.59 |
|  | White | 83.44 | 87.49 | 80.14 | 79.09 |
|  | Other | 0.59 | 1.02 | 10.86 | 11.69 |
| Region | Midwest | 10.07 | 23.39 | 13.21 | 12.79 |
|  | Northeast | 15.28 | 19.84 | 29.07 | 29.25 |
|  | South | 20.40 | 34.96 | 37.70 | 38.29 |
|  | West | 54.26 | 21.8%0 | 18.88 | 18.42 |
|  | Other | NA | NA | 1.14 | 1.25 |
| Metastatic at diagnosis | Yes | 54.11 | 55.30 | 61.07 | 62.65 |
|  | No | 45.89 | 44.70 | 38.93 | 37.35 |
| Stage | Stage I | 26.14 | NA | 17.33 | 16.21 |
|  | Stage II | 19.62 | NA | 21.60 | 21.14 |
|  | Stage III | 18.58 | NA | 23.89 | 24.8 |
|  | Stage IV | 35.66 | NA | 37.18 | 37.85 |
| Year of diagnosis | 2011 | 15.86 | 16.04 | 13.34 | 9.40 |
|  | 2012 | 16.56 | 16.35 | 15.44 | 10.89 |
|  | 2013 | 16.45 | 16.74 | 16.75 | 11.80 |
|  | 2014 | 16.96 | 16.97 | 16.95 | 11.95 |
|  | 2015 | 17.08 | 17.10 | 18.68 | 13.16 |
|  | 2016 | 17.08 | 16.80 | 18.85 | 13.29 |
|  | 2017 - 2019 | NA | NA | 0 | 29.51 |

G. Follicular lymphoma
|  |  | SEER, % | NPCR, % | Flatiron Health, % |  |
| --- | --- | --- | --- | --- | --- |
|  |  |  |  | 2011-2016 | 2011-2019 |
| Sex | Female | 48.85 | 49.79 | 50.62 | 49.91 |
|  | Male | 51.15 | 50.21 | 49.38 | 50.09 |
| Age at diagnosis | 0-19 | 0.29 | 0.28 | 0.25 | 0.18 |
|  | 20-34 | 1.68 | 1.61 | 1.49 | 1.25 |
|  | 35-49 | 12.05 | 11.09 | 10.20 | 10.04 |
|  | 50-64 | 35.47 | 34.29 | 35.57 | 33.51 |
|  | 65-79 | 37.28 | 38.86 | 40.92 | 41.85 |
|  | 80+ | 13.23 | 13.88 | 11.57 | 13.17 |
| Race | Asian | 5.28 | 2.40 | 1.23 | 1.58 |
|  | Black | 4.77 | 4.93 | 4.50 | 4.35 |
|  | White | 89.47 | 91.73 | 83.79 | 81.60 |
|  | Other | 0.47 | 0.95 | 10.49 | 12.46 |
| Region | Midwest | 9.71 | 23.35 | 14.99 | 14.78 |
|  | Northeast | 16.45 | 19.84 | 27.71 | 26.75 |
|  | South | 21.06% | 36.77 | 35.89 | 36.99 |
|  | West | 52.78 | 20.04 | 20.15 | 20.04 |
|  | Other | NA | NA | 1.26 | 1.45 |
| Metastatic at diagnosis | Yes | 56.19 | 55.30 | 70.04 | 70.35 |
|  | No | 43.81 | 44.7%0 | 29.96 | 29.65 |
| Stage | Stage I | 27.40 | NA | 17.35 | 17.07 |
|  | Stage II | 15.85 | NA | 12.61 | 12.58 |
|  | Stage III | 28.01 | NA | 30.71 | 30.82 |
|  | Stage IV | 28.75 | NA | 39.33 | 39.53 |
| Year of diagnosis | 2011 | 16.25 | 16.89 | 15.17 | 10.93 |
|  | 2012 | 16.19 | 16.33 | 13.56 | 9.77 |
|  | 2013 | 16.12 | 16.72 | 16.67 | 12.01 |
|  | 2014 | 17.22 | 16.88 | 16.92 | 12.19 |
|  | 2015 | 16.92 | 16.79 | 19.90 | 14.34 |
|  | 2016 | 17.30 | 16.40 | 17.79 | 12.81 |
|  | 2017 - 2019 | NA | NA | 0 | 27.96 |

H. Advanced gastric/esophageal carcinoma
|  |  | SEER, % | NPCR, % | Flatiron Health, % |  |
| --- | --- | --- | --- | --- | --- |
|  |  |  |  | 2011-2016 | 2011-2019 |
| Sex | Female | 33.47 | 31.74 | 26.55 | 26.51 |
|  | Male | 66.53 | 68.26 | 73.45 | 73.49 |
| Age at diagnosis | 0-19 | 0.08 | 0.07 | 0.00 | 0.00 |
|  | 20-34 | 1.25 | 1.09 | 0.89 | 0.83 |
|  | 35-49 | 7.55 | 7.19 | 6.80 | 6.58 |
|  | 50-64 | 30.9 | 31.58 | 32.16 | 31.31 |
|  | 65-79 | 39.88 | 40.89 | 43.54 | 43.96 |
|  | 80+ | 20.35 | 19.19 | 16.62 | 17.33 |
| Race | Asian | 10.45 | 5.13 | 3.42 | 3.70 |
|  | Black | 12.34 | 13.00 | 8.26 | 8.73 |
|  | White | 76.38 | 80.66 | 75.55 | 74.1 |
|  | Other | 0.84 | 1.20 | 12.77 | 13.48 |
| Region | Midwest | 8.91 | 21.93 | 13.50 | 13.40 |
|  | Northeast | 17.2 | 21.09 | 26.88 | 26.53 |
|  | South | 21.55 | 35.99 | 35.67 | 35.92 |
|  | West | 52.34 | 20.98 | 22.38 | 22.46 |
|  | Other | NA | NA | 1.57 | 1.69 |
| Metastatic at diagnosis | Yes | 39.14 | 38.36 | 49.47 | 50.87 |
|  | No | 60.86 | 61.64 | 50.53 | 49.13 |
| Stage | Stage 0 | 0.30 | NA | NA | NA |
|  | Stage I | 23.70 | NA | 5.01 | 5.32 |
|  | Stage II | 16.48 | NA | 15.18 | 14.93 |
|  | Stage III | 19.34 | NA | 23.95 | 22.52 |
|  | Stage IV | 40.18 | NA | 55.87 | 57.24 |
| Year of diagnosis | 2011 | 16.10 | 16.19 | 11.39 | 7.97 |
|  | 2012 | 16.53 | 16.31 | 14.53 | 10.17 |
|  | 2013 | 16.48 | 16.61 | 16.78 | 11.74 |
|  | 2014 | 16.96 | 16.88 | 19.09 | 13.36 |
|  | 2015 | 17.12 | 16.99 | 18.95 | 13.26 |
|  | 2016 | 16.82 | 17.02 | 19.27 | 13.49 |
|  | 2017 - 2019 | NA | NA | 0 | 27.29 |
|  | Prior to 2011 | NA | NA | 0 | 2.72 |

I. Hepatocellular carcinoma (HCC)
|  |  | SEER, % | NPCR, % | Flatiron Health, % |  |
| --- | --- | --- | --- | --- | --- |
|  |  |  |  | EDM 2011-2016 | 2011-2019 |
| Sex | Female | 23.21 | 22.74 | 24.06 | 23.51 |
|  | Male | 76.79 | 77.26 | 75.94 | 76.49 |
| Age at diagnosis | 0-19 | 0.17 | 0.19 | 0.00 | 0.00 |
|  | 20-34 | 0.59 | 0.62 | 0.55 | 0.45 |
|  | 35-49 | 4.52 | 4.49 | 3.39 | 3.02 |
|  | 50-64 | 49.45 | 50.53 | 44.17 | 41.74 |
|  | 65-79 | 35.29 | 34.57 | 40.71 | 43.77 |
|  | 80+ | 9.98 | 9.59 | 11.19 | 11.02 |
| Race | Asian | 14.81 | 7.87 | 5.61 | 5.41 |
|  | Black | 14.04% | 16.10 | 12.33 | 12.48 |
|  | White | 69.74% | 74.09 | 66.32 | 66.06 |
|  | Other | 1.41 | 1.93 | 15.74 | 16.04 |
| Region | Midwest | 7.15 | 17.38 | 7.30 | 8.03 |
|  | Northeast | 12.61 | 18.55 | 31.89 | 31.37 |
|  | South | 20.10 | 38.88 | 35.85 | 36.56 |
|  | West | 60.14 | 25.19 | 23.51 | 22.50 |
|  | Other | NA | NA | 1.45 | 1.55 |
| Metastatic at diagnosis | Yes | 15.22 | 15.95 | 25.27 | 27.46 |
|  | No | 84.78 | 84.05 | 74.73 | 72.54 |
| Stage | Stage I | 40.76 | NA | 21.39 | 18.68 |
|  | Stage II | 20.54 | NA | 14.58 | 14.55 |
|  | Stage III | 19.13 | NA | 22.58 | 21.78 |
|  | Stage IV | 19.57 | NA | 41.46 | 44.99 |
| Year of diagnosis | 2011 | 15.10 | 14.79 | 11.01 | 7.54 |
|  | 2012 | 16.01 | 15.82 | 15.16 | 10.39 |
|  | 2013 | 16.70 | 16.49 | 16.07 | 11.02 |
|  | 2014 | 17.35 | 17.39 | 16.98 | 11.64 |
|  | 2015 | 17.73 | 17.89 | 20.01 | 13.71 |
|  | 2016 | 17.12 | 17.64 | 20.77 | 14.24 |
|  | 2017 - 2019 | NA | NA | 0 | 31.45 |

J. Advanced head and neck cancer
|  |  | SEER, % | NPCR, % | Flatiron Health, % |  |
| --- | --- | --- | --- | --- | --- |
|  |  |  |  | 2011-2016 | 2011-2019 |
| Sex | Female | 26.24 | 26.20 | 23.07 | 23.31 |
|  | Male | 73.76 | 73.80 | 76.93 | 76.69 |
| Age at diagnosis | 0-19 | 0.32 | 0.31 | 0.00 | 0.00 |
|  | 20-34 | 1.28 | 1.18 | 0.47 | 0.64 |
|  | 35-49 | 9.43 | 9.38 | 7.40 | 7.81 |
|  | 50-64 | 42.81 | 43.76 | 46.18 | 46.42 |
|  | 65-79 | 34.56 | 34.78 | 37.59 | 37.28 |
|  | 80+ | 11.60 | 10.60 | 8.36 | 7.85 |
| Race | Asian | 6.13 | 2.92 | 1.77 | 1.58 |
|  | Black | 10.10 | 9.84% | 7.34 | 7.69 |
|  | White | 83.10 | 86.27 | 79.80 | 79.32 |
|  | Other | 0.68 | 0.97 | 11.08 | 11.41 |
| Region | Midwest | 9.99 | 22.82 | 11.58 | 11.52 |
|  | Northeast | 15.15 | 18.21 | 19.60 | 19.35 |
|  | South | 27.22 | 40.29 | 51.51 | 51.92 |
|  | West | 47.64 | 18.68 | 14.90 | 15.07 |
|  | Other | NA | NA | 2.41 | 2.13 |
| Metastatic at diagnosis | Yes | 15.80 | 17.69 | 71.35 | 71.25 |
|  | No | 84.20 | 82.31 | 28.65 | 28.75 |
| Stage | Stage I | 24.08 | NA | 6.09 | 6.54 |
|  | Stage II | 12.76 | NA | 7.72 | 7.72 |
|  | Stage III | 15.74 | NA | 14.84 | 14.45 |
|  | Stage IV | 47.42 | NA | 71.35 | 71.25 |
|  | Stage 0 | NA | NA | 0 | 0.04 |
| Year of diagnosis | 2011 | 15.85 | 15.92 | 12.75 | 9.05 |
|  | 2012 | 16.03 | 16.03 | 16.05 | 11.40 |
|  | 2013 | 16.62 | 16.64 | 16.25 | 11.54 |
|  | 2014 | 17.02 | 16.95 | 17.65 | 12.53 |
|  | 2015 | 17.23 | 17.21 | 17.85 | 12.68 |
|  | 2016 | 17.25 | 17.23 | 19.45 | 13.81 |
|  | 2017 - 2019 | NA | NA | 0 | 17.07 |
|  | Prior to 2011 | NA | NA | 0 | 11.93 |

K. Advanced melanoma
|  |  | SEER, % | NPCR, % | Flatiron Health, % |  |
| --- | --- | --- | --- | --- | --- |
|  |  |  |  | 2011-2016 | 2011-2019 |
| Sex | Female | 40.73 | 41.04 | 33.57 | 34.93 |
|  | Male | 59.27 | 58.96 | 66.43 | 65.07 |
| Age at diagnosis | 0-19 | 0.43 | 0.43 | 0.31 | 0.36 |
|  | 20-34 | 5.42 | 5.40 | 5.05 | 5.42 |
|  | 35-49 | 13.64 | 13.67 | 11.93 | 13.54 |
|  | 50-64 | 31.24 | 30.81 | 30.93 | 30.84 |
|  | 65-79 | 33.18 | 33.70 | 37.72 | 36.62 |
|  | 80+ | 16.09 | 16.00 | 14.05 | 13.22 |
| Race | Asian | 0.66 | 0.32 | 0.28 | 0.28 |
|  | Black | 0.46 | 0.50 | 0.47 | 0.45 |
|  | White | 98.64 | 98.72 | 92.91 | 92.13 |
|  | Other | 0.25 | 0.45 | 6.34 | 7.14 |
| Region | Midwest | 7.74 | 21.74 | 14.10 | 14.15 |
|  | Northeast | 14.43 | 18.49 | 22.21 | 22.05 |
|  | South | 22.10 | 35.85 | 33.44 | 34.68 |
|  | West | 55.74 | 23.92 | 29.82 | 28.67 |
|  | Other | NA | NA | 0.44 | 0.45 |
| Metastatic at diagnosis | Yes | 4.51 | 5.50 | 23.27 | 23.62 |
|  | No | 95.49 | 94.50 | 76.73 | 76.38 |
| Stage | Stage 0 | 0 | NA | 0.31 | 0.37 |
|  | Stage I | 75.38 | NA | 7.88 | 9.60 |
|  | Stage II | 12.97 | NA | 16.98 | 17.16 |
|  | Stage III | 7.45 | NA | 50.34 | 47.46 |
|  | Stage IV | 4.20 | NA | 24.48 | 25.41 |
| Year of diagnosis | 2011 | 14.79 | 15.13 | 13.91 | 8.87 |
|  | 2012 | 15.67 | 15.46 | 15.46 | 9.86 |
|  | 2013 | 16.25 | 16.23 | 16.95 | 10.81 |
|  | 2014 | 17.55 | 17.18 | 17.83 | 11.37 |
|  | 2015 | 17.99 | 17.96 | 18.32 | 11.69 |
|  | 2016 | 17.74 | 18.04 | 17.54 | 11.18 |
|  | 2017 - 2019 | NA | NA | 0 | 19.72 |
|  | Prior to 2011 | NA | NA | 0 | 16.50 |

L. Malignant pleural mesothelioma
|  |  | SEER, % | NPCR, % | Flatiron Health, % |  |
| --- | --- | --- | --- | --- | --- |
|  |  |  |  | 2011-2016 | 2011-2019 |
| Sex | Female | 25.01 | 24.75 | 21.17 | 22.00 |
|  | Male | 74.99 | 75.25 | 78.83 | 78.00 |
| Age at diagnosis | 0-19 | 0.02 | 0.10 | 0.00 | 0.00 |
|  | 20-34 | 1.01 | 0.87 | 0.14 | 0.10 |
|  | 35-49 | 4.11 | 3.65 | 1.29 | 1.33 |
|  | 50-64 | 18.02 | 17.80 | 15.33 | 14.37 |
|  | 65-79 | 45.41 | 47.31 | 58.14 | 57.82 |
|  | 80+ | 31.43 | 30.27 | 25.10 | 26.38 |
| Race | Asian | 3.63 | 1.58 | 0.84 | 0.94 |
|  | Black | 5.19 | 4.83 | 3.19% | 2.98 |
|  | White | 90.62 | 92.90 | 87.77 | 87.31 |
|  | Other | 0.56 | 0.69 | 8.21 | 8.77 |
| Region | Midwest | 8.76 | 23.64 | 11.15 | 10.87 |
|  | Northeast | 19.29 | 21.76 | 38.95 | 37.64 |
|  | South | 19.11 | 32.80 | 36.55 | 38.09 |
|  | West | 52.84 | 21.81 | 13.01 | 13.01 |
|  | Other | NA | NA | 0.34 | 0.40 |
| Metastatic at diagnosis | Yes | NA | 69.80 | 14.59 | 16.34 |
|  | No | NA | 30.20 | 85.41 | 83.66 |
| Stage | Stage I | 21.84 | NA | 8.08% | 9.42 |
|  | Stage II | 10.47 | NA | 10.77 | 9.50 |
|  | Stage III | 22.57 | NA | 32.07 | 30.86 |
|  | Stage IV | 45.13 | NA | 49.08 | 50.22 |
| Year of diagnosis | 2011 | 16.85 | 17.12 | 15.54 | 11.27 |
|  | 2012 | 16.51 | 16.83 | 15.26 | 11.07 |
|  | 2013 | 16.75 | 16.79 | 16.49 | 11.96 |
|  | 2014 | 16.59 | 16.84 | 17.44 | 12.65 |
|  | 2015 | 17.60 | 16.64 | 16.89 | 12.25 |
|  | 2016 | 15.71 | 15.79 | 18.39 | 13.34 |
|  | 2017 - 2019 | NA | NA | 0 | 27.46 |

M. Multiple myeloma
|  |  | SEER, % | NPCR, % | Flatiron Health, % |  |
| --- | --- | --- | --- | --- | --- |
|  |  |  |  | 2011-2016 | 2011-2019 |
| Sex | Female | 43.80 | 44.08 | 46.98 | 46.06 |
|  | Male | 56.20 | 55.92 | 53.02 | 53.94 |
| Age at diagnosis | 0-19 | 0.03 | 0.02 | 0.00 | 0.00 |
|  | 20-34 | 0.50 | 0.46 | 0.39 | 0.35 |
|  | 35-49 | 6.52 | 6.40 | 5.50 | 5.38 |
|  | 50-64 | 30.00 | 30.02 | 30.75 | 29.61 |
|  | 65-79 | 43.25 | 44.12 | 45.91 | 46.95 |
|  | 80+ | 19.70 | 18.98 | 17.45 | 17.71 |
| Race | Asian | 5.73 | 2.62 | 1.95 | 1.95 |
|  | Black | 20.37 | 20.62 | 17.66 | 17.97 |
|  | White | 73.26 | 75.70 | 68.62 | 67.7 |
|  | Other | 0.64 | 1.05 | 11.77 | 12.37 |
| Region | Midwest | 10.00 | 21.08 | 12.90 | 12.76 |
|  | Northeast | 16.67 | 20.00 | 28.40 | 28.00 |
|  | South | 25.12 | 39.99 | 37.82 | 38.41 |
|  | West | 48.21 | 18.93 | 19.29 | 19.22 |
|  | Other | NA | NA | 1.58 | 1.61 |
| Metastatic at diagnosis | Yes | 95.58 | 95.41 | 100.00 | 100.00 |
|  | No | 4.42 | 4.59 | NA | NA |
| Stage | Stage I | NA | NA | 34.58 | 34.22 |
|  | Stage II | NA | NA | 33.03 | 33.06 |
|  | Stage III | NA | NA | 32.38 | 32.73 |
| Year of diagnosis | 2011 | 15.51 | 15.67 | 12.79 | 9.14 |
|  | 2012 | 16.19 | 16.09 | 15.03 | 10.74 |
|  | 2013 | 16.46 | 16.67 | 16.53 | 11.81 |
|  | 2014 | 16.69 | 17.04 | 16.82 | 12.02 |
|  | 2015 | 17.42 | 17.38 | 18.90 | 13.50 |
|  | 2016 | 17.73 | 17.15 | 19.94 | 14.24 |
|  | 2017 - 2019 | NA | NA | 0 | 28.56 |

N. Advanced NSCLC
|  |  | SEER, % | NPCR, % | Flatiron Health, % |  |
| --- | --- | --- | --- | --- | --- |
|  |  |  |  | 2011-2016 | 2011-2019 |
| Sex | Female | 47.28 | 46.56 | 47.42 | 47.40 |
|  | Male | 52.72 | 53.44 | 52.58 | 52.60 |
| Age at diagnosis | 0-19 | 0.01 | 0.01 | 0.01 | 0.01 |
|  | 20-34 | 0.17 | 0.14 | 0.20 | 0.19 |
|  | 35-49 | 3.12 | 3.18 | 3.69 | 3.59 |
|  | 50-64 | 27.80 | 28.75 | 30.63 | 30.47 |
|  | 65-79 | 50.26 | 50.6 | 51.01 | 51.18 |
|  | 80+ | 18.65 | 17.32 | 14.47 | 14.56 |
| Race | Asian | 7.13 | 2.96 | 2.72 | 2.82 |
|  | Black | 11.84 | 11.30 | 9.31 | 9.34 |
|  | White | 80.52 | 84.96 | 78.11 | 77.72 |
|  | Other | 0.51 | 0.78 | 9.87 | 10.11 |
| Region | Midwest | 10.97 | 24.06 | 15.27 | 15.19 |
|  | Northeast | 17.18 | 19.72 | 27.43 | 27.46 |
|  | South | 27.93 | 39.43 | 39.46 | 39.72 |
|  | West | 43.91 | 16.78 | 17.16 | 16.98 |
|  | Other | NA | NA | 0.68 | 0.64 |
| Metastatic at diagnosis | Yes | 52.03 | 49.75 | 64.18 | 64.43 |
|  | No | 47.97 | 50.25 | 35.82 | 35.57 |
| Stage | Stage I | 24.81 | NA | 8.50 | 9.08 |
|  | Stage II | 7.46 | NA | 5.49 | 5.39 |
|  | Stage III | 19.01 | NA | 21.81 | 21.10 |
|  | Stage IV | 48.73 | NA | 64.18 | 64.43 |
|  | Stage 0 | NA | NA | 0.01 | 0.01 |
| Year of diagnosis | 2011 | 16.51 | 16.33 | 12.36 | 8.63 |
|  | 2012 | 16.59 | 16.50 | 15.20 | 10.61 |
|  | 2013 | 16.61 | 16.64 | 17.17 | 11.99 |
|  | 2014 | 16.72 | 16.80 | 18.17 | 12.69 |
|  | 2015 | 16.82 | 16.93 | 18.91 | 13.21 |
|  | 2016 | 16.74 | 16.80 | 18.20 | 12.71 |
|  | 2017 - 2019 | NA | NA | 0 | 24.67 |
|  | Prior to 2011 | NA | NA | 0 | 5.47 |

O. Ovarian carcinoma
|  |  | SEER, % | NPCR, % | Flatiron Health, % |  |
| --- | --- | --- | --- | --- | --- |
|  |  |  |  | 2011-2016 | 2011-2019 |
| Sex | Female | 100.00 | 100.00 | 100.00 | 100.00 |
|  | Male | NA | 0 | NA | NA |
| Age at diagnosis | 0-19 | 1.30 | 1.26 | 0.23 | 0.17 |
|  | 20-34 | 3.56 | 3.45 | 1.98 | 1.94 |
|  | 35-49 | 13.15 | 12.55 | 11.59 | 11.26 |
|  | 50-64 | 35.13 | 34.86 | 36.89 | 36.14 |
|  | 65-79 | 32.62 | 34.15 | 38.91 | 39.46 |
|  | 80+ | 14.25 | 13.74) | 10.40 | 11.01 |
| Race | Asian | 8.73 | 4.26 | 2.67 | 2.53 |
|  | Black | 8.95 | 9.18 | 6.20 | 6.16 |
|  | White | 81.62 | 85.39 | 78.84 | 78.51 |
|  | Other | 0.69 | 1.17 | 12.29 | 12.80 |
| Region | Midwest | 9.15 | 21.86 | 13.37 | 12.68 |
|  | Northeast | 16.39 | 20.23 | 22.00 | 21.52 |
|  | South | 19.74 | 35.53 | 41.30 | 42.13 |
|  | West | 54.72 | 22.38 | 21.63 | 21.69 |
|  | Other | NA | NA | 1.72 | 1.98 |
| Metastatic at diagnosis | Yes | 60.80 | 57.35 | 21.38 | 22.84 |
|  | No | 39.20 | 42.65 | 78.62 | 77.16 |
| Stage | Stage I | 23.50 | NA | 20.13 | 19.63 |
|  | Stage II | 8.73 | NA | 8.91 | 9.02 |
|  | Stage III | 39.02 | NA | 48.69 | 47.65 |
|  | Stage IV | 28.75 | NA | 22.26 | 23.70 |
| Year of diagnosis | 2011 | 16.40 | 16.61 | 14.81 | 10.90 |
|  | 2012 | 16.58 | 16.67 | 16.70 | 12.30 |
|  | 2013 | 16.58 | 16.68 | 16.66 | 12.27 |
|  | 2014 | 16.87 | 16.78 | 17.17 | 12.64 |
|  | 2015 | 17.12 | 17.00 | 16.70 | 12.30 |
|  | 2016 | 16.44 | 16.26 | 17.97 | 13.24 |
|  | 2017 - 2019 | NA | NA | 0 | 26.35 |

P. Metastatic pancreatic carcinoma
|  |  | SEER, % | NPCR, % | Flatiron Health, % |  |
| --- | --- | --- | --- | --- | --- |
|  |  |  |  | 2011-2016 | 2011-2019 |
| Sex | Female | 48.94 | 48.13 | 46.88 | 46.36 |
|  | Male | 51.06 | 51.87 | 53.12 | 53.64 |
| Age at diagnosis | 0-19 | 0.13 | 0.12 | 0.00 | 0.00 |
|  | 20-34 | 0.68 | 0.60 | 0.28 | 0.26 |
|  | 35-49 | 4.55 | 4.72 | 3.94 | 3.87 |
|  | 50-64 | 27.81 | 28.79 | 32.67 | 31.72 |
|  | 65-79 | 43.03 | 44.34 | 50.77 | 51.49 |
|  | 80+ | 23.80 | 21.44 | 12.34 | 12.66 |
| Race | Asian | 7.70 | 3.38 | 1.82 | 1.82 |
|  | Black | 12.12 | 12.60 | 9.02 | 9.34 |
|  | White | 79.51 | 83.05 | 79.29 | 77.81 |
|  | Other | 0.67 | 0.97 | 9.86 | 11.03 |
| Region | Midwest | 9.88 | 22.11 | 12.18 | 12.05 |
|  | Northeast | 17.03 | 20.15 | 30.45 | 29.58 |
|  | South | 22.64 | 37.18 | 39.90 | 40.85 |
|  | West | 50.45 | 20.56 | 16.53 | 16.66 |
|  | Other | NA | NA | 0.93 | 0.85 |
| Metastatic at diagnosis | Metastatic | 54.43 | 53.34 | 64.99 | 71.74 |
|  | Not Metastatic | 45.57 | 46.66 | 35.01 | 28.26 |
| Stage | Stage 0 | 0 | NA | NA | NA |
|  | Stage I | 12.23 | NA | 4.11 | 3.57 |
|  | Stage II | 25.64 | NA | 23.32 | 18.10 |
|  | Stage III | 9.05 | NA | 7.59 | 6.58 |
|  | Stage IV | 53.08 | NA | 64.99 | 71.74 |
| Year of diagnosis | 2011 | NA | 0 | 0.57 | 0.34 |
|  | 2012 | NA | 0 | 2.22 | 1.34 |
|  | 2013 | NA | 0 | 7.44 | 4.50 |
|  | 2014 | 32.84 | 32.62 | 27.57 | 16.67 |
|  | 2015 | 33.76 | 33.55 | 31.19 | 18.85 |
|  | 2016 | 33.4 | 33.83 | 31.01 | 18.75 |
|  | 2017 - 2019 | NA | NA | 0 | 38.85 |
|  | Prior to 2011 | NA | NA | 0 | 0.70 |

Q. Metastatic prostate cancer
|  |  | SEER | NPCR | Flatiron Health, % |  |
| --- | --- | --- | --- | --- | --- |
|  |  |  |  | 2011-2016 | 2011-2019 |
| Sex | Male | 100.00 | 100.00 | 100.00 | 100.00 |
|  | Female | NA | 0 | NA | NA |
| Age at diagnosis | 0-19 | 0 | 0 | 0.00 | 0.00 |
|  | 20-34 | 0.01 | 0.01 | 0.00 | 0.01 |
|  | 35-49 | 2.22 | 2.19 | 1.64 | 1.90 |
|  | 50-64 | 38.73 | 39.22 | 26.79 | 34.08 |
|  | 65-79 | 49.77 | 50.22 | 51.70 | 50.23 |
|  | 80+ | 9.27 | 8.36 | 19.88 | 13.78 |
| Race | Asian | 5.09 | 2.32 | 1.85 | 1.66 |
|  | Black | 16.43 | 16.76 | 11.70 | 10.65 |
|  | White | 78.04 | 79.88 | 74.54 | 75.88 |
|  | Other | 0.43 | 1.04 | 11.91 | 11.81 |
| Region | Midwest | 9.55 | 21.95 | 13.50 | 12.92 |
|  | Northeast | 17.70 | 20.21 | 20.30 | 21.24 |
|  | South | 24.40 | 37.99 | 43.47 | 43.58 |
|  | West | 48.34 | 19.84 | 21.24 | 21.00 |
|  | Other | NA | NA | 1.48 | 1.25 |
| Metastatic at diagnosis | Yes | 7.69 | 7.56 | 74.24 | 59.70 |
|  | No | 92.31 | 92.44 | 25.76 | 40.30 |
| Stage | Stage I | 6.26 | NA | 0.62 | 1.32 |
|  | Stage II | 72.41 | NA | 5.51 | 9.25 |
|  | Stage III | 9.69 | NA | 3.48 | 5.17 |
|  | Stage IV | 11.63 | NA | 90.38 | 84.25 |
| Year of diagnosis | 2011 | NA | 0 | 7.87 | 3.65 |
|  | 2012 | NA | 0 | 8.14 | 3.78 |
|  | 2013 | 25.16 | 24.83 | 21.00 | 9.74 |
|  | 2014 | 23.62 | 23.94 | 21.22 | 9.84 |
|  | 2015 | 25.11 | 25.43 | 20.45 | 9.49 |
|  | 2016 | 26.11 | 25.80 | 21.32 | 9.89 |
|  | 2017 - 2019 | NA | NA | 0 | 16.59 |
|  | Prior to 2011 | NA | NA | 0 | 37.03 |

R. Metastatic renal-cell carcinoma (RCC)
|  |  | SEER, % | NPCR, % | Flatiron Health, % |  |
| --- | --- | --- | --- | --- | --- |
|  |  |  |  | 2011-2016 | 2011-2019 |
| Sex | Female | 36.25 | 36.88 | 30.69 | 31.40 |
|  | Male | 63.75 | 63.12 | 69.31 | 68.60 |
| Age at diagnosis | 0-19 | 1.16 | 1.05 | 0.00 | 0.04 |
|  | 20-34 | 2.01 | 1.85 | 0.67 | 0.77 |
|  | 35-49 | 12.58 | 11.86 | 7.63 | 9.08 |
|  | 50-64 | 36.85 | 36.03 | 39.11 | 40.75 |
|  | 65-79 | 36.42 | 37.76 | 43.15 | 41.19 |
|  | 80+ | 10.98 | 11.45 | 9.44 | 8.17 |
| Race | Asian | 5.31 | 2.31 | 1.83 | 1.63 |
|  | Black | 12.68 | 12.11 | 7.90 | 7.56 |
|  | White | 81.02 | 84.15 | 76.23 | 76.81 |
|  | Other | 0.99 | 1.44 | 14.04 | 14.00 |
| Region | Midwest | 9.81 | 22.96 | 16.56 | 15.64 |
|  | Northeast | 15.17 | 18.44 | 20.20 | 20.46 |
|  | South | 25.44 | 38.77 | 39.38 | 40.30 |
|  | West | 49.58 | 19.82 | 22.66 | 22.41 |
|  | Other | NA | NA | 1.21 | 1.19 |
| Metastatic at diagnosis | Yes | 14.61 | 14.68 | 60.21 | 53.60 |
|  | No | 85.39 | 85.32 | 39.79 | 46.40 |
| Stage | Stage I | 62.56 | NA | NA | NA |
|  | Stage II | 7.81 | NA | NA | NA |
|  | Stage III | 13.72% | NA | NA | NA |
|  | Stage IV | 15.91 | NA | 62.24 | 55.12 |
|  | Unspecified Non-metastatic | NA | NA | 37.76 | 44.88 |
| Year of diagnosis | 2011 | 15.42 | 15.44 | 14.12 | 8.70 |
|  | 2012 | 16.03 | 15.97 | 15.01 | 9.25 |
|  | 2013 | 16.43 | 16.37 | 16.94 | 10.43 |
|  | 2014 | 16.89 | 16.95 | 18.14 | 11.18 |
|  | 2015 | 17.65 | 17.52 | 18.01 | 11.09 |
|  | 2016 | 17.58 | 17.75 | 17.79 | 10.95 |
|  | 2017 - 2019 | NA | NA | 0 | 18.53 |
|  | Prior to 2011 | NA | NA | 0 | 19.87 |

S. Small-cell lung cancer (SCLC).
|  |  | SEER, % | NPCR, % | Flatiron Health, % |  |
| --- | --- | --- | --- | --- | --- |
|  |  |  |  | 2011-2016 | 2011-2019 |
| Sex | Female | 50.55 | 50.94 | 51.61 | 52.48 |
|  | Male | 49.45 | 49.06 | 48.39 | 47.52 |
| Age at diagnosis | 0-19 | 0.01 | 0 | 0.00 | 0.00 |
|  | 20-34 | 0.07 | 0.06 | 0.05 | 0.10 |
|  | 35-49 | 2.59 | 2.75 | 2.76 | 2.60 |
|  | 50-64 | 33.05 | 33.92 | 35.62 | 34.63 |
|  | 65-79 | 51.29 | 51.26 | 51.69 | 52.57 |
|  | 80+ | 13.00 | 12.00 | 9.88 | 10.10 |
| Race | Asian | 3.65 | 1.39 | 0.93 | 0.96 |
|  | Black | 8.98 | 7.86 | 6.08 | 6.23 |
|  | White | 86.76 | 89.92 | 84.00 | 83.19 |
|  | Other | 0.62 | 0.83 | 8.99 | 9.61 |
| Region | Midwest | 13.23 | 26.71 | 18.11 | 17.48 |
|  | Northeast | 14.94 | 17.35 | 24.35 | 23.53 |
|  | South | 35.67 | 41.98 | 43.76 | 44.52 |
|  | West | 36.16 | 13.96 | 13.42 | 14.11 |
|  | Other | NA | NA | 0.36 | 0.36% |
| Metastatic at diagnosis | Metastatic | 74.60 | 72.14 | 71.75 | 70.86 |
|  | Not Metastatic | 25.40 | 27.86 | 28.25 | 29.14 |
| Stage | Stage I | 4.17 | NA | 4.07 | 4.27 |
|  | Stage II | 3.04 | NA | 4.44 | 4.42 |
|  | Stage III | 22.38 | NA | 19.75 | 20.45 |
|  | Stage IV | 70.40 | NA | 71.75 | 70.86 |
| Year of diagnosis | 2013 | 25.12 | 25.08 | 23.34 | 14.77 |
|  | 2014 | 25.75 | 25.07 | 24.90 | 15.76 |
|  | 2015 | 25.33 | 25.39 | 25.31 | 16.01 |
|  | 2016 | 23.80 | 24.45 | 26.46 | 16.74 |
|  | 2017 - 2019 | NA | NA | 0 | 36.72 |

## APPENDIX II: APRIL 2023 UPDATE

Differences in age distribution between the three databases were previously noted in this study, such that a lower proportion of patients over 80 years of age at diagnosis was seen in the Flatiron Health database as compared with SEER and NPCR sources. Of note, birth year data for elderly patients in this analysis was subject to a standard algorithmic transformation in order to mitigate their higher risk of patient re-identification. In December 2022, Flatiron Health instituted an improvement to this algorithm that reduced the extent of birth year transformation and, hence, calculated age at diagnosis. The improved algorithm enables the reporting of the true birth year for a greater share (>90%) of patients than in the original algorithm. Using the improved algorithm with patient records available in the Flatiron Health databases for analysis as of May 31, 2019 and December 31, 2016 results in changes to the distributions previously reported in patients’ age at initial diagnosis. These changes are incorporated into Tables 2 through 20 and A3.A through A3.S herein (previously reported distributions are available in older versions of this paper).

For most disease-specific databases, the updated birth years resulted in an increase of patient records showing a diagnosis at 80 years or greater. Most updated distributions match their counterpart SEER and/or NPCR distributions to within 5% for each age category. The greatest changes appear among patients with malignant pleural mesothelioma (MPM), with, for example, 25.1% of the MPM patients available for analysis with an initial diagnosis from January 2011 to December 2016 showing an age of 80 years or greater at diagnosis, versus 14.65% prior to the update. These findings suggest that similarity in age distribution between Flatiron Health, SEER, and NPCR is greater than initially reported, as differences seen in the original dataset were largely related to the prior approach to algorithmic masking of birth year for elderly patients.

## Notes

### Competing Interest Statement

At the time of the study, all authors report employment at Flatiron Health, Inc., which is an independent member of the Roche Group, and stock ownership in Roche. SSB, LL own equity in Flatiron Health.

### Clinical Trial

Not applicable

### Funding Statement

This study was sponsored by Flatiron Health, Inc. (Flatiron Health), which is an independent member of the Roche group.

### Summary of Updates

The analysis of patients in the Flatiron Health databases was refreshed in April 2023 to incorporate an update to the birth year variable, reflecting best practices in patient de-identification. This refresh resulted in updates to the distributions in calculated age at initial diagnosis. These updates have been incorporated in the methods section, Tables 2 through 20 and A3.A through A3.S, and a new appendix (Appendix II). Additionally, race/ethnicity and sex/gender variables have been clarified in Table 1.

